# A multi-ancestry polygenic risk score for Alzheimer disease is associated with cognitive decline, hippocampal atrophy and neuropathological hallmarks in diverse populations

**DOI:** 10.1101/2025.09.24.25336555

**Authors:** Nuzulul Kurniansyah, Shinya Tasaki, Habbibur Rehman, Congcong Zhu, John Farrell, Richard Sherva, Richard Hauger, Victoria C. Merritt, Matthew Panizzon, Rui Zhang, J. Michael Gaziano, Jungsoo Gim, Kunho Lee, Dong Yong Lee, Kwansik Nho, Ricardo A. Vialle, Shubhabrata Mukherjee, Emily H. Trittschuh, Annie J. Lee, Adam M. Brickman, Carlos Cruchaga, Shannon Risacher, Douglas N. Greve, Paul Crane, VA Million Veteran Program, Alzheimer’s Disease Genetics Consortium, Alzheimer Disease Sequencing Project, Eden Martin, William Bush, Richard Mayeux, Jonathan L. Haines, Margaret A. Pericak-Vance, Mark Logue, David A. Bennett, Lisa L. Barnes, Andrew Saykin, Timothy Hohman, Li-San Wang, Gerard C. Schellenberg, Ting Fang Alvin Ang, Rhoda Au, Jesse Mez, Kathryn L. Lunetta, Xiaoling Zhang, Lindsay A. Farrer

## Abstract

Alzheimer disease (AD) has a strong genetic basis, yet previously derived polygenic risk scores (PRS) are heavily weighted by the *APOE* locus and perform inconsistently across diverse ancestries. We developed an *APOE*-independent multi-ancestry AD PRS using genome-wide association study summary statistics from cohorts in the United States, Europe and East Asia that were applied to European ancestry (EA), African American (AA), Caribbean Hispanic (CH), and East Asian cohorts from the Alzheimer’s Disease Genetics Consortium. PRS performance was evaluated in the multi-ancestry Alzheimer’s Disease Sequencing Project (ADSP) dataset and validated in several additional multi-ancestry cohorts. The PRS was significantly associated with AD in the ADSP EA, AA, CH, and Native American Hispanic groups with adjusted odds ratios (ORs) between 1.14 and 1.52 per standard deviation of the PRS. PRS performance was validated in the replication cohorts (ORs 1.21-1.65). The PRS was also associated with poorer memory, executive function, and language performance; greater AD-related neuropathological burden (including CERAD, Braak stage, and Thal phase scores); reduced hippocampal volume; lower CSF Aβ42; and elevated total tau and phosphorylated tau (p-tau), with stronger p-tau associations observed in women. Longitudinal analyses revealed that individuals in the highest PRS decile exhibited the steepest cognitive decline, particularly among those who progressed to AD. Our findings demonstrate the utility of an ancestry-aware and APOE-independent PRS for advancing understanding of the genetic basis of AD across diverse populations. Associations observed with early biological and cognitive changes and potential sex-specific differences support the incorporation of a PRS in clinical trials and personalized intervention and prevention strategies.

## Main

Late-onset Alzheimer disease (AD) is a progressive disorder characterized by cognitive decline and memory impairment, affecting an estimated 6.9 million people in the United States ^1^. The clinical course of AD unfolds over many years, often beginning with subtle cognitive changes before progressing to overt dementia. Although the rate of decline varies across individuals, growing evidence suggests that genetic factors play a central role in shaping both the onset and trajectory of cognitive deterioration ^2^. While the *APOE ε4* allele remains the strongest known genetic risk factor ^3,4^, genome-wide association studies (GWAS) have identified numerous additional common and rare variants contributing to polygenic susceptibility primarily in individuals of European ancestry (EA) ^5,6^ and to a lesser extent in other populations^7,8^.

Polygenic risk scores (PRS) aggregate the effects of multiple genetic variants into a single quantitative measure of inherited susceptibility, each of which is weighted based on GWAS summary statistics ^9^. A critical challenge in the application of PRS lies in the underrepresentation of diverse genetic ancestries in AD GWAS datasets, which include predominantly white individuals of European ancestry (EA). Although AD PRS and genetic risk scores derived from European ancestry (EA) cohorts show predictive value in several non-European populations, their performance varies by ancestry, with notably reduced predictive value in some underrepresented groups ^10–12^. The availability of GWAS data from diverse populations, including African Americans (AA) ^7,13^, Caribbean Hispanics (CH) ^14^, and East Asian (EAS) ^1516^ provides an opportunity to enhance the transferability and accuracy of PRS across multiple ancestries. This expanded genetic representation is crucial for developing more robust and universally applicable PRS models, which can improve risk prediction.

AD PRSs have been associated with cerebrospinal fluid (CSF) biomarkers, neuropathological features, hippocampal atrophy, and cognitive decline^17–21^ However, these findings have largely been derived from studies in EA populations, limiting their relevance to other populations. A broader evaluation of PRS in ancestrally diverse settings is essential to determine its clinical utility and potential role in early detection^11^. We developed a multi-ancestry AD PRS using GWAS summary statistics from multiple population ancestries. A weighted PRS summation was derived from the multi-ancestry datasets assembled by the Alzheimer’s Disease Genetics Consortium (ADGC) and evaluated for performance with and without variants in the *APOE* region across diverse ancestries represented in the AD Sequencing Project (ADSP). To ensure robustness, the PRS was validated in independent cohorts, including the multi-ancestry All of Us (AoU), EA participants from the Framingham Heart Study (FHS), the Korean Brain Aging Study for the Early Diagnosis and Prediction of AD (KBASE) Study, and AA participants from the Religious Orders Study and Memory Aging Project (ROSMAP), Minority Aging Research Study (MARS), and Rush Alzheimer’s Disease Center (collectively referred to as Rush). We examined its association with CSF and plasma biomarkers, a brain MRI measure of hippocampal volume, neuropathological traits, and longitudinal trajectories of cognitive decline to assess whether genetic factors influence early manifestations of AD and the degree to which these effects differ by sex and ancestry.

## Results

### Overall study design for the development and evaluation of a multi-ancestry AD PRS

**Figure 1** provides an overview of the study. We first derived several PRS using multiple AD GWAS datasets including EA, AA, and EAS individuals that were constructed using clumping and thresholding via PRSice2 and Bayesian shrinkage methods using LDPred2, PRS-CS, and PRS-CSx. We applied three strategies to develop a robust multi-ancestry AD PRS, including (1) combining GWAS results from all datasets by meta-analysis first, then constructing the PRS, (2) weighted summation with weights derived from the ADGC dataset, and (3) unweighted summation. PRS performance was evaluated in the ADSP dataset for AD risk prediction, selecting the best-performing PRS based on the largest odds ratio (OR), highest area under the curve (AUC), and lowest p-value. To further refine the most robust PRS, we recalculated PRS weights after removing SNPs within 1 Mb of the *APOE* coding region to eliminate the confounding effect of *APOE* genotype and applying the same methods. The final *APOE* region-excluded PRS was validated in independent cohorts, including AoU version 7, KBASE, FHS, and Rush. We then examined associations of the AD PRS with AD biomarkers, cognitive function, hippocampal volume, and neuropathological traits.

**Figure 1.**
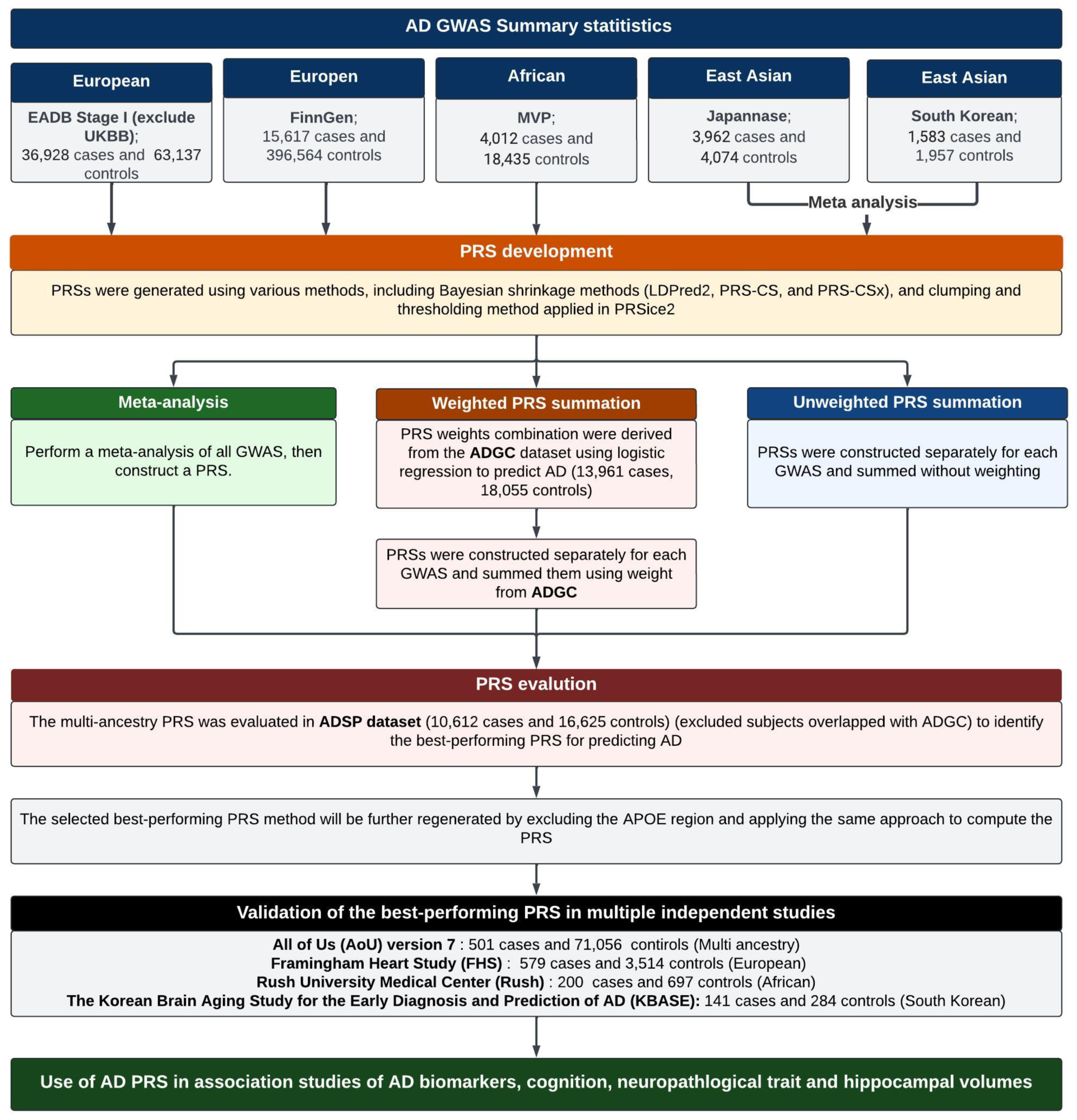
The overview of the study. Development and evaluation of AD PRS. The process involves: (a) PRS development using clumping-thresholding and Bayesian shrinkage approaches; (b) multi-ancestry PRS evaluation using meta-analysis, unweighted PRS summation, and weighted PRS summation with weights derived from ADGC; (c) evaluation of PRS performance in the ADSP dataset; (d) refinement of the best-performing PRS by excluding the *APOE* region, followed by validation of this PRS in independent datasets; (e) Evaluation of the association of the AD PRS with AD-related phenotypes.

### Multi-ancestry PRS strongly associated with AD risk in diverse ancestry groups

Evaluation of population substructure among 36,362 individuals in the ADSP Release 4 dataset by principal component (PC) analysis identified six ancestry clusters shown in **Supplementary Figure 1**: European (EA, N=16,840), African (AA, N=6,532), Caribbean Hispanic (CH, N=5,317), Native American Hispanic (NAH, N=4,722), South Asian (SA, N=2,772), and East Asian (EAS, N=178). Ancestry inference estimates highlight the genetic diversity and composition within each major ancestry group (**Supplementary Figure 2**). Individuals within the EA, AA, NAH, SA, and EAS clusters appear relatively homogeneous, whereas significant genetic admixture was observed in the CH cluster. After excluding individuals with missing AD status, age or sex, 26,874 individuals of diverse ancestral backgrounds remained for analysis (**Supplementary Table 1**).

We compared the performance of a multi-ancestry AD PRS, constructed using GWAS summary statistics from ancestrally diverse populations in the ADSP dataset. Three different PRS construction strategies were evaluated, including one based on PRS weight summation described in the **Methods** that was derived from the ADGC dataset (**Supplementary Table 2**). The PRS-CS weighted summation method consistently outperformed other strategies, both with and without adjustment for *APOE* ɛ4 carrier status (**Supplementary Figure 3**), and this PRS construction was employed in subsequent analyses. To assess the specific impact of variants in the *APOE* region on PRS performance, we recalculated the PRS after removing SNPs within 1 Mb of the *APOE* coding region from the training data using the PRS-CS weighted summation approach. The PRS distributions among *APOE* ε4 carriers and non-carriers were nearly identical in all ancestries, whereas on average, ε4 carriers in all groups had a higher PRS when *APOE* region SNPs were included in the PRS (**Figure 2A**). These findings suggest the disproportionate effect of *APOE* genotype on PRS compared to the collective contribution of SNPs in other portions of the genome.

**Figure 2.**
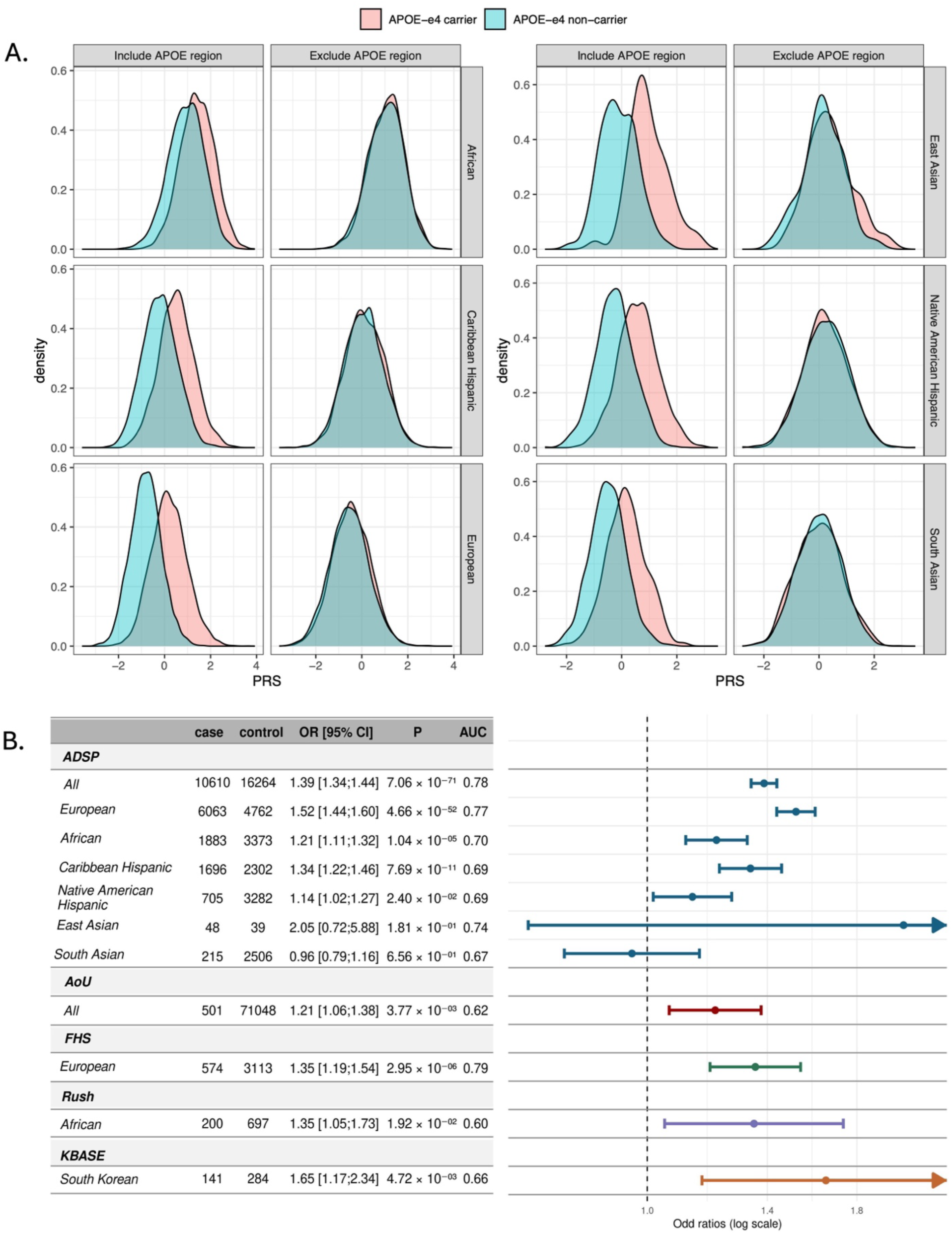
AD PRS characteristics and behavior. **A.** Distribution of AD PRS calculated with and without the APOE region by ancestry and ε4 carrier status. **B.** Association of AD PRS excluded APOE region (per 1 SD increase) with AD in the ADSP (stratified by ancestry), FHS, and RADC cohorts. Analyses were adjusted for age, APOE ε4 and ε2, sex, and the first 10 principal components of ancestry. In FHS, models were further adjusted for study cohort and family ID as random effects. Both FHS and RADC models also adjusted for education. In all participants ADSP and AoU, models included additional adjustment for ancestry group. AUC: Area under the curve, CI: confidence interval, OR: Odds ratio; SD: standard deviation

Next, we evaluated the performance of the multi-ancestry AD PRS with and without SNPs in the *APOE* region across ADSP ancestry groups. The PRS including *APOE* region SNPs was significantly associated with AD in all ancestry groups except SA and EAS populations due to small sample sizes. The performance of the PRS was attenuated after adjusting for *APOE* ε4 and ε2 carrier status (**Supplementary Figure 4**). PRS performance remained robust even after excluding the *APOE* region, with models adjusted for *APOE* ε4 and ε2 carrier status showing slightly stronger associations than unadjusted models (**Supplementary Figure 5**). When including APOE ε4 and ε2 carrier status in the model, the AD PRS excluding the *APOE* region was significantly associated with AD (OR per 1 standard deviation [SD] increase = 1.39, 95% CI: 1.34– 1.44, P = 7.06 × 10⁻⁷¹, AUC = 0.78) in the total sample (**Figure 2B**). Among ancestry groups, the strongest association was observed in EAs (OR = 1.52, 95% CI: 1.44–1.60, P = 4.66×10⁻⁵², AUC = 0.77), followed by the CH (OR = 1.34, 95% CI: 1.22–1.46, P = 7.69×10⁻¹¹, AUC = 0.69), AA (OR = 1.21, 95% CI: 1.11–1.32, P = 1.04×10⁻⁵, AUC = 0.70), and NAH (OR = 1.14, 95% CI: 1.02–1.27, P = 2.40×10⁻², AUC = 0.69) groups. No significant associations were observed for the SA or EAS groups.

The PRS excluding the *APOE* region was selected for subsequent analyses of AD and related outcomes because it provided a robust measure of genetic risk, captured ancestry-specific effects, and minimized the influence of the large effect of *APOE* genotype on the overall score. Further analysis showed that the association between PRS and AD risk was similar in men and women across all ancestry groups, except in the NAH group in which the PRS was not significantly associated with AD in men (**Supplementary Figure 6**). Examination within *APOE* genotype in the overall sample revealed that the association of the AD PRS was significant in all genotype groups, but the OR for ε2 carriers (1.16) was substantially smaller than the ORs for the ε3/ε3 (1.37), ε3/ε4 (1.45) and ε4/ε4 (1.52) groups (**Supplementary Figure 7**). These patterns were most evident in the EA group, while effect sizes were attenuated in the AA group. In the CH sample, the strength of association was similar across ε2/ε2+ε2/ε3, ε3/ε3, and ε3/ε4 genotypes (ORs = 1.33–1.39). The numbers of ε4 homozygotes in this group and NAH ε2 and ε4 carriers was too small to draw reliable conclusions.

### AD PRS validation in the independent cohorts

We validated the robust performance of the AD PRS in three independent cohorts: KBASE which includes 425 South Koreans (**Supplementary Table 1),** AoU (version 7) which includes 71,549 individuals from diverse ancestries (**Supplementary Table 3**), Rush comprising 897 AA participants (**Supplementary Table 4**), and FHS which includes 4,080 EA participants (**Supplementary Table 5**). The AD PRS was significantly associated with AD (per 1 SD increase) in the AoU (OR = 1.21, 95% CI: 1.06–1.38, P = 3.77 × 10⁻³, AUC = 0.62), Rush (OR = 1.35, 95% CI: 1.05–1.73, P = 1.92 × 10⁻², AUC = 0.60), KBASE (OR = 1.65, 95% CI: 1.17–2.34, P = 4.72 × 10⁻^3^, AUC = 0.66), and FHS (OR = 1.35, 95% CI: 1.19–1.54, P = 2.95 × 10⁻⁶, AUC = 0.79) cohorts (**Figure 2B**). Further analysis to evaluate sex differences revealed that in the FHS dataset the AD PRS was significantly associated with AD in women (OR = 1.42, 95% CI: 1.23-1.65, P =2.22 × 10⁻⁶ ), but not men (OR = 1.15, 95% CI: 0.94-1.41, P =0.17). However, no sex-specific association was observed in the larger ADSP cohort (**Supplementary Figure 6**). The AD PRS was not significant in either men or women in the Rush cohort, likely because of small sample sizes (**Supplementary Figure 8**).

To contextualize the performance of our multi-ancestry AD PRS, we compared it to a PRS that we calculated using previously published AD-specific variant weights from the polygenic score (PGS) catalog and recent literature. This allowed us to assess whether our approach improves prediction across diverse populations. We restricted comparisons to scores explicitly developed for AD. For consistency, we downloaded variant weights (excluding the *APOE* region) and applied them to the ADSP and KBASE datasets to construct PRSs and directly compare effect sizes. Because all published scores were derived from GWAS that included European samples from ADGC and ADSP, we focused our comparisons on non-European ancestry groups in ADSP (AA, CH, and NAH) to avoid sample overlap. We tested the performance of each PRS within ancestry groups in ADSP and in KBASE using logistic regression models adjusted for age, sex, *APOE* ε4/ε2 status, and the top ten PCs. In the ADSP dataset, our PRS showed the strongest association with AD in the AA and CH groups, but was less predictive in the NAH group compared to the score by Lourida et al ^22^. (**Supplementary Table 6**). Our PRS consistently outperformed all other scores in the Korean KBASE dataset (**Supplementary Table 7**).

### Association of AD PRS with cerebrospinal fluid (CSF) and plasma AD biomarkers

We assessed the association of AD PRS with amyloid-β 42 (Aβ42), total Tau (t-Tau), and phosphorylated Tau 181 (pTau181) measured in CSF obtained from 1,389 ADSP participants (49.7% men) whose characteristics are shown in **Supplementary Table 8**. All biomarker values were harmonized and standardized across cohorts prior to analysis. A 1 SD higher AD PRS was significantly associated with reduced Aβ42 level (β = -0.14, 95% CI: -0.20 to -0.08, P = 1.03 × 10⁻⁶), increased pTau181 (β = 0.14, 95% CI: 0.07 to 0.20, P = 2.03 × 10⁻⁵) and increased t-Tau (β = 0.18, 95% CI: 0.12 to 0.24, P = 1.79 × 10⁻⁸). The association with pTau181 was significant only in women (β = 0.21, 95% CI: 0.12 to 0.30, P = 2.76 × 10⁻⁶) (**Figure 3A**).

**Figure 3.**
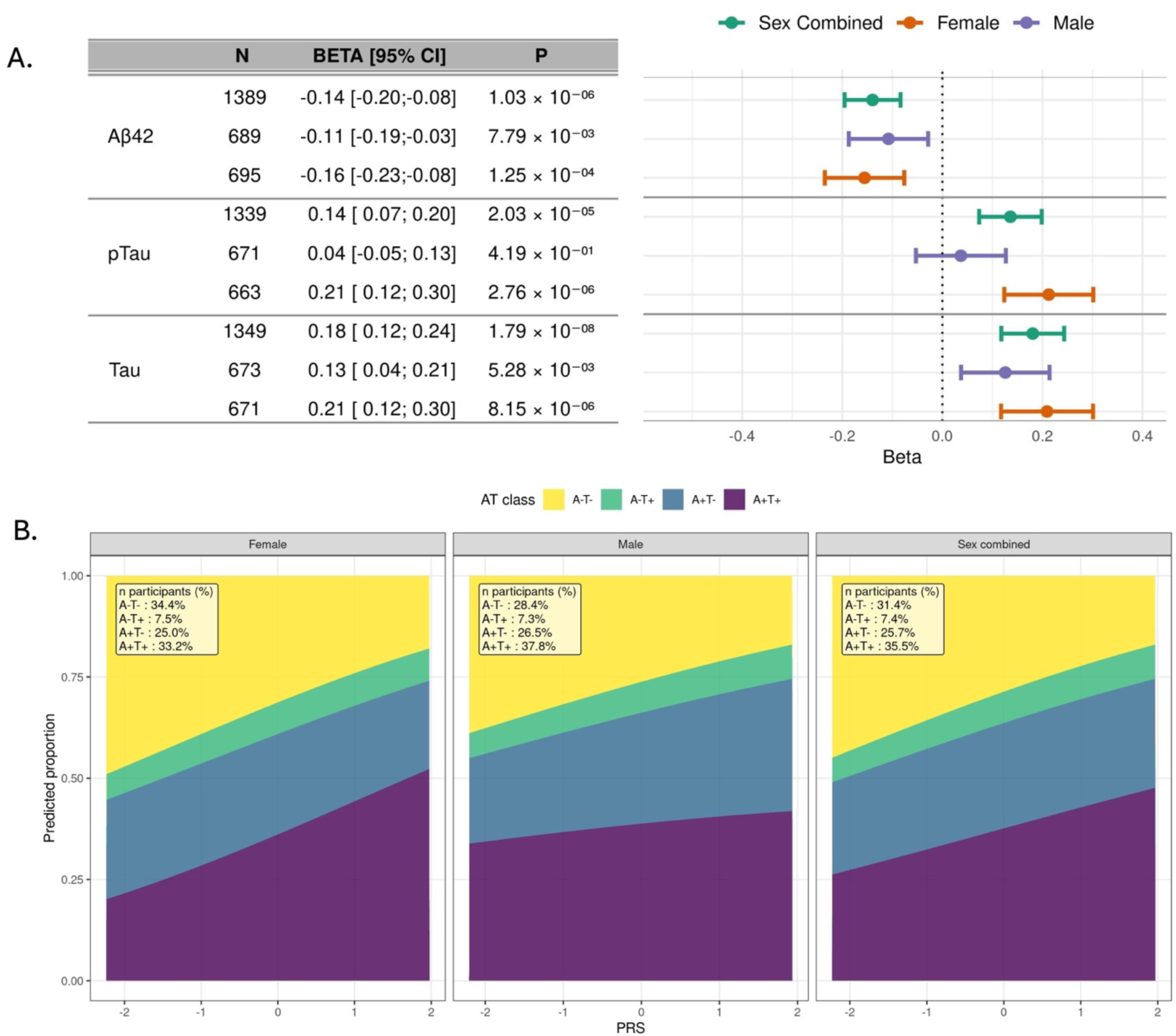
Associations of the AD PRS with AD, CSF biomarkers and AT classification in the ADSP dataset. **A.** Association of the AD PRS and CSF biomarkers (Aβ42, pTau, and Tau) stratified by sex. Beta coefficients represent the change in standardized CSF biomarker level per 1 SD increase in the AD PRS. Analysis models included covariates for age at CSF biomarker measurement, sex (unless stratified), education, APOE ε4 and ε2 status, ancestry group and the first 10 principal components of ancestry. **B.** Predicted probabilities for each AT category are shown across PRS values, with other covariates fixed at their observed values. Models were adjusted for the same covariates as in panel A. The AT classification categorizes biomarkers as follows: A+/T+ (amyloid and tau pathology present), A–/T+ (tau pathology without amyloid) ,A+/T– (amyloid pathology without tau involvement), and A–/T– (neither amyloid nor tau pathology present). CSF: cerebrospinal fluid; CI: confidence interval, SD: standard deviation.

We further investigated these associations by modeling amyloid and tau (AT) classification, where A+T+ represents amyloid and tau positivity consistent with AD pathology; A+T− reflects amyloid positivity without tau accumulation; A−T+ suggests non-AD tauopathies; and A−T− indicates no AD-related pathology. AT classification was modeled as the outcome in a multinomial logistic regression with AD PRS as a continuous predictor, adjusting for age, *APOE* ε4 and ε2 carrier status, and the top ten ancestry PCs. In the overall sample, higher PRS was associated with a higher predicted probability of A+T+ and a lower probability of A−T−, indicating a graded increase in biomarker-defined AD pathology across the PRS distribution (**Figure 3B**). In sex-stratified models, men had a higher predicted probability of A+T+ at lower PRS values, whereas women showed a steeper increase in A+T+ probability across the PRS range, suggesting that women may experience a more pronounced shift toward biomarker-positive AD pathology as genetic risk increases.

We also examined the association of AD PRS with plasma p-tau181 in the Framingham Heart Study (FHS; n = 1,680) and longitudinal plasma p-tau217 in the Alzheimer’s Disease Neuroimaging Initiative (ADNI; n = 301) cohorts (**Supplementary Tables 8 and 9**). A 1 SD higher AD PRS was associated with elevated p-tau181 levels (β = 0.10, 95% CI: 0.02–0.17, P = 0.01), an effect that was evident in women (β = 0.11, 95% CI: 0.01–0.20, P = 0.03) but not men (β = 0.05, 95% CI: –0.08 to 0.18, P = 0.45). A higher PRS was similarly associated with elevated p-tau217 (β = 0.20, 95% CI: 0.05– 0.35, P = 8.9 × 10⁻³) which was evident in women (β = 0.33, 95% CI: 0.10–0.57, P = 7.6 × 10⁻³) but not men (β = 0.02, 95% CI: –0.17 to 0.22, P = 0.82) (**Supplementary Figure 9**).

### Association of AD PRS with neuropathological traits

We assessed the association of the AD PRS (per 1 SD increase) with AD-related neuropathological traits among ADSP participants (**Supplementary Table 10**). Neuropathological traits were modeled as ordered categorical outcomes, preserving the full range of each scale. A 1 SD higher AD PRS was significantly associated with greater severity across all traits: CERAD score for neuritic plaque burden (OR = 1.37, 95% CI: 1.25–1.49, *P* = 3.95×10⁻¹²), amyloid deposition measured by Thal phase (OR = 1.38, 95% CI: 1.20–1.58, *P* = 6.90×10⁻⁶), Braak stage (OR = 1.41, 95% CI: 1.30–1.53, *P* = 7.15×10⁻¹⁵), and AD neuropathologic change (ADNC) score (OR = 1.52, 95% CI: 1.30–1.76, *P* = 6.80×10⁻⁸) (**Supplementary Figure 10**).

### Association of AD PRS with cognitive performance

We assessed the association of the AD PRS with co-calibrated, harmonized and standardized scores for memory, executive function, and language, derived from longitudinal cognitive assessments across multiple time points in ADSP, KBASE, and FHS participants (**Supplementary Table 11**). The derivation of cognitive domain scores is detailed in the **Methods**. Each 1 SD increase in AD PRS was significantly associated with poorer performance across all time points for memory (β = -0.17, 95% CI: -0.19 to - 0.57, P = 1.44 × 10⁻^68^), executive function (β = -0.11, 95% CI: -0.13 to -0.10, P = 1.01 × 10⁻^33^), and language (β = -0.12, 95% CI: -0.14 to -0.10, P = 7.01 × 10^⁻39^). Associations of AD PRS with cognitive performance scores were significant in the EA, AA, and CH groups but not in the NAH group (**Figure 4, Supplementary Figures 11 and 12**). Similar trends were observed in FHS participants for all cognitive domains but for memory only in KBASE participants. Further examination of the association of the AD PRS with episodic memory and global cognition in Rush AA participants (**Supplementary Table 4**) showed that a 1 SD higher AD PRS was significantly associated with lower episodic memory (β = -0.20, 95% CI: -0.31 to -0.10, P = 1.62 × 10⁻⁴) and global cognition (β = -0.12, 95% CI: -0.21 to -0.04, P = 5.99 × 10⁻³) scores (**Supplementary Figure 13**).

**Figure 4.**
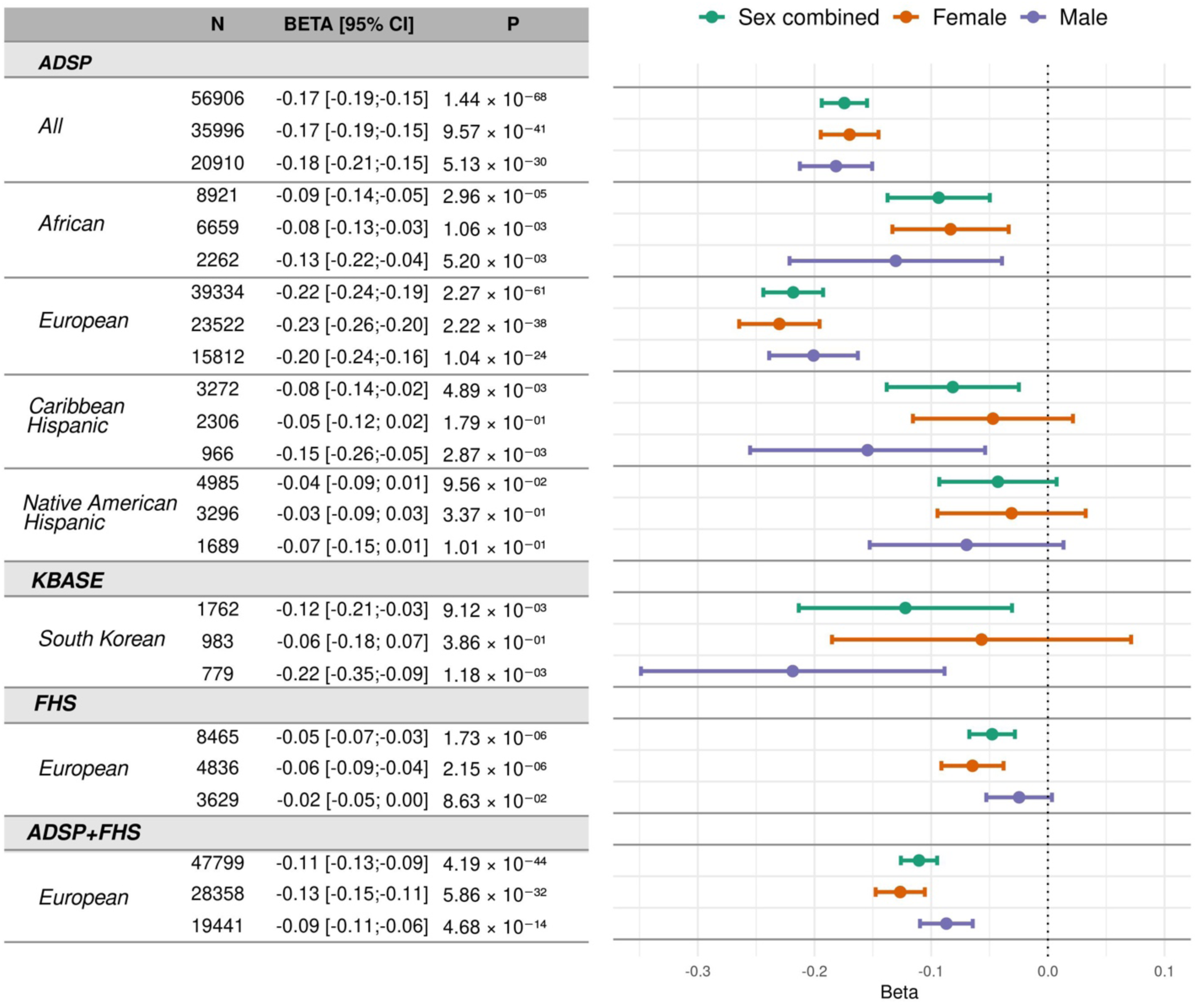
Association of AD PRS with memory score. Beta coefficients represent the decrease in standardized memory scores per 1 SD increase in the AD PRS. Analyses were adjusted for age at exam, sex (unless stratified), education, ancestry (in the combined ADSP dataset), APOE ε4 and ε2 carrier status, and the first 10 principal components of ancestry. In the FHS dataset, models also included covariates for generation cohort and family ID as random effects. CI: confidence interval, SD: standard deviation.

Sex-stratified analyses conducted in the ADSP and FHS datasets showed that the association of a 1 SD higher AD PRS was more strongly associated with lower memory performance in women than men, particularly among EA participants as indicated by a meta-analysis of results from both datasets in women (β = -0.13, 95% CI: -0.15 to -0.11, P = 5.86 × 10⁻^32^) compared to men (β = -0.09, 95% CI: -0.11 to -0.06, P = 4.68 × 10⁻^14^) (**Figure 4**). In contrast, associations of AD PRS with executive function and language scores were similar between sexes across all ancestries (**Supplementary Figures 11 and 12**). Sex-stratified analyses in the Rush and KBASE cohorts yielded comparable results (**Supplementary Figure 13**).

### AD PRS predicts age-related cognitive decline

We estimated trajectories of cognitive performance between ages 60 and 80 using generalized additive mixed models (GAMMs), modeling age as a smooth term and stratifying individuals by AD PRS percentiles (<10%, 10–50%, 50–90%, >90%). Predicted scores for performance in memory, executive function, and language domains were derived from longitudinal cognitive assessments in ADSP participants. Details of the derivation of cognitive domain scores and modeling procedures are described in the Methods section. As shown in **Figure 5**, among all ADSP participants, individuals in the highest PRS stratum (>90%) had lower memory performance from age 60 onward, with the performance gap relative to lower-risk groups widening with age. Similar but less pronounced differences were observed for executive function and language. These findings suggest that higher AD polygenic risk is associated with lower cognitive performance in later life, particularly in memory, with largely parallel rates of age-related decline. Further analysis of cognitive trajectories by sex and revealed no significant differences in the slope or level of decline across PRS strata between males and females in the overall sample (**Supplementary Figure 14**).

**Figure 5.**
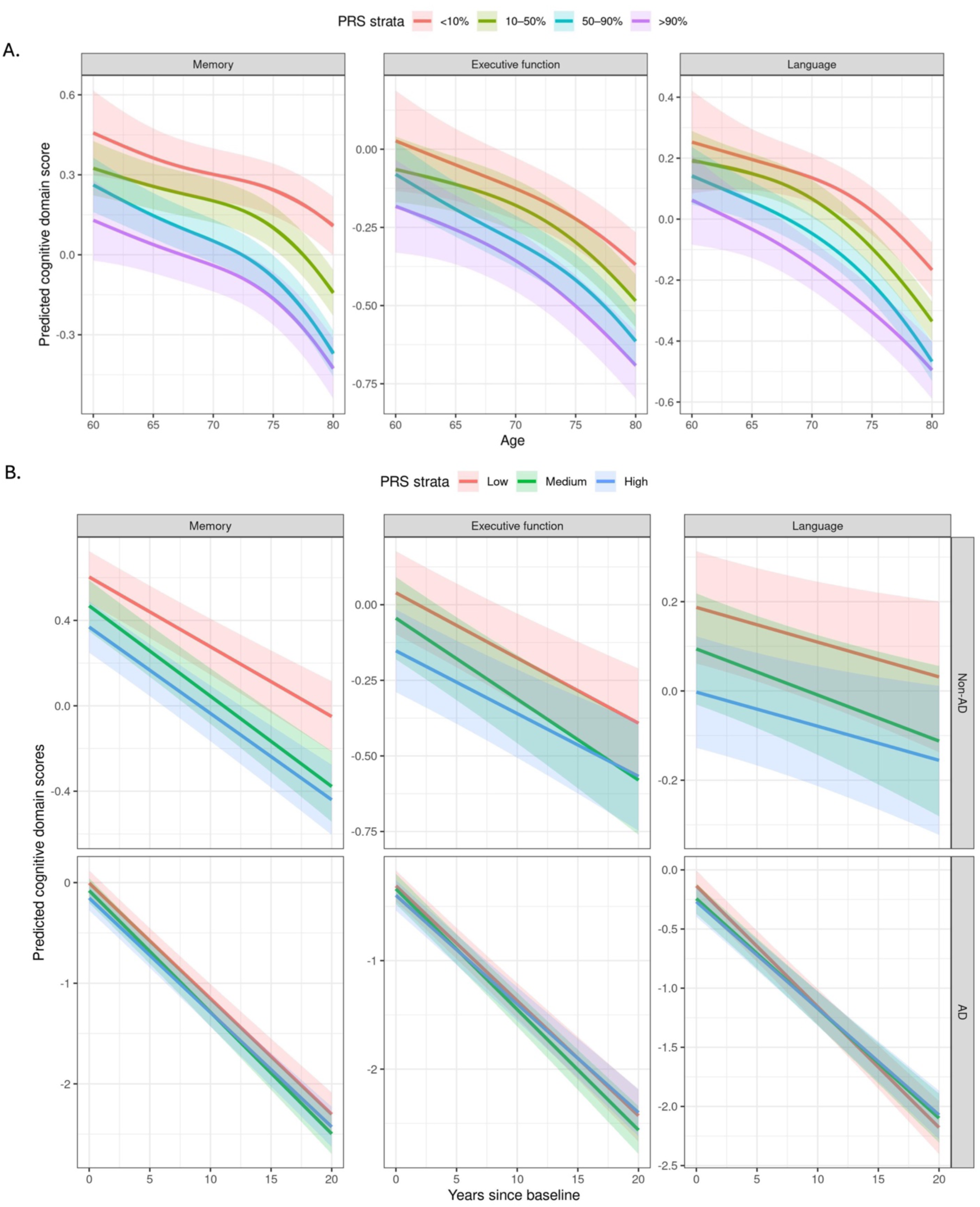
Age-related and longitudinal trajectories of cognitive performance by AD PRS in ADSP participants. Analyses included 32,167 participants in the ADSP and were adjusted for sex, education, APOE ε4 and ε2 status, ancestry group, and the first 10 principal components of ancestry. **A.** Age-related trajectories for memory, executive function, and language across PRS groups (<10%, 10–50%, 50–90%, and >90%). Solid lines depict predicted cognitive scores by age, with shaded ribbons indicating 95% confidence intervals. **B.** Longitudinal trajectories in the same domains, stratified by AD status. Solid lines depict predicted scores over time, with shaded ribbons indicating 95% confidence intervals. Models included main effects and all two-way and three-way interactions between PRS group, AD status, and time since baseline.

To test whether AD polygenic risk influences cognitive decline differentially by diagnostic status, we fit linear mixed-effects models including PRS group (low, moderate, high), AD status (AD vs. non-AD), time since first visit, and their interactions. Individuals who developed AD exhibited significantly steeper decline across memory, executive function, and language domains compared to those who remained cognitively normal (**Figure 5B).** Among AD cases, we observed subtle trends toward faster memory decline in those with a higher PRS, particularly in the years preceding clinical diagnosis. However, differences across PRS groups were modest and diminished after diagnosis. For executive function and language, PRS-related effects were minimal. Cognitive trajectories among non-AD participants declined slowly and remained largely parallel across PRS groups. Individuals in the high PRS group consistently exhibited lower cognitive scores, particularly in the memory domain. These results indicate that the AD PRS primarily captures differences in cognitive performance that emerge before clinical diagnosis, rather than predicting the rate of decline after diagnosis.

### AD PRS is associated with hippocampal atrophy

We examined the association of AD PRS with hippocampal volume using longitudinal FreeSurfer-derived structural MRI data from ADSP (n= 6,568) and FHS (n= 970) participants ages 60 and older (**Supplementary Table 12**). Hippocampal volume was adjusted for head size by normalizing to intracranial volume and subsequently log-transformed. Effect sizes reflect the change in log-transformed hippocampal volume normalized to intracranial volume, per one standard deviation increase in AD PRS. Higher AD PRS was significantly associated with reduced hippocampal volume across all time points in both cohorts (ADSP: β = –0.021, 95% CI: –0.028 to –0.013, P = 1.37 × 10⁻⁷; FHS: β = –0.017, 95% CI: –0.029 to –0.005, P = 5.23 × 10⁻³). This association was observed separately in both women and men in ADSP (women: β = –0.024, 95% CI: –0.035 to –0.014, P = 3.14 × 10⁻⁶; men: β = –0.016, 95% CI: –0.028 to –0.005, P = 6.60 × 10⁻³), whereas in FHS the association was evident only in women (β = –0.019, 95% CI: –0.035 to –0.003, P = 2.33 × 10⁻²) and not men (β = –0.017, 95% CI: –0.036 to 0.002, P = 0.073) (**Supplementary Figure 15**).

## Discussion

We developed and validated a PRS that captures the influence of common genetic variation for and is highly predictive of AD across diverse ancestries. Although *APOE* ε4 is the strongest common genetic risk factor for AD, our PRS excludes variants in the *APOE* region, which thus enables assessment and highlights the contribution of polygenic risk independent of *APOE* genotype. The score was significantly associated with clinical AD diagnosis, cognitive performance, hippocampal volume, neuropathological measures of AD severity, and CSF and plasma AD biomarkers.

This study addresses critical challenges in developing and generalizing an AD PRS across ancestrally diverse populations. Rather than relying solely on European-ancestry GWAS, we integrated summary statistics from diverse ancestries and employed ancestry-aware modeling strategies. Our multi-ancestry PRS outperformed comparative scores from the PGS Catalog and the recently published EA PRS by Nicolas et al. ^12^ in AA, CH, and EAS groups within the ADSP and KBASE datasets. The observed PRS signal in the CH group likely reflects shared African and European components represented in the training data, enabling more accurate tagging of risk variants through overlapping LD structure^23^. In contrast, the strongest AD PRS association in the NAH group was observed with the EA-derived PRS by Lourida et al ^22^, a counterintuitive result likely driven by residual European admixture and coincidental LD alignment, rather than genuine cross-population transferability, particularly given the absence of NAH GWAS data ^24–27^. Our findings underscore the potential improvement in performance of a PRS derived from multi-ancestry populations when applied to underrepresented populations, despite imbalance sample sizes.^28^. However, the diminished performance in Native American–enriched groups highlight ongoing limitations when entire ancestries are excluded from the construction of the PRS. Prior studies have shown that differences in LD structure, allele frequencies, and effect size distributions across populations limit PRS portability ^26,27^. Improving risk prediction in admixed and underrepresented populations will require expanded GWAS representation and analytical approaches that explicitly model ancestry-specific genomic architecture.

The best-performing PRS included variants in the *APOE* region but its association with AD was markedly attenuated after adjustment for *APOE* ε4 and ε2 genotype, confirming that the signal was largely *APOE* driven. To capture risk independent of *APOE*, we constructed a PRS excluding and modeled *APOE* genotype separately; it remained significantly associated with AD and showed comparable effect sizes in ε4 carriers and non-carriers, demonstrating additional polygenic risk beyond *APOE* genotype. Excluding this region may also reduce ancestry-related variability linked to local ancestry, cis- and trans-modifiers, and vascular burden^29–33^. These findings support integrating both *APOE* genotype and an *APOE*-excluded PRS for more precise risk stratification, enabling early identification even among ε4 non carriers. Translation into clinical practice, however, will require addressing variability in PRS performance across ancestries^34^, limited integration of genetic data into clinical workflows, and the absence of standardized thresholds for clinical interpretation ^35^.

Understanding the downstream effects of genetic liability measures is critical for developing biologically informed and clinically relevant tools for risk assessment. In this study, the AD PRS was associated with multiple domains of AD pathophysiology, including neuropathology, CSF and plasma biomarkers, cognition, and hippocampal volume. Significant associations were observed with CERAD plaque burden, Thal phase, Braak stage, and overall AD-related neuropathological severity, aligning with established staging frameworks and supporting the PRS as a marker of cumulative disease burden ^36^. Our AD PRS was also linked to lower CSF Aβ42 and higher t-tau, pTau181 and pTau 217 concentrations, consistent with early amyloid accumulation and downstream tau pathology characteristic of the pre-clinical stage of AD^37^. Using AT classification based on CSF amyloid and tau status, higher PRS was associated with a graded pattern across biomarker profiles, with increasing PRS corresponding to a higher predicted probability of A+T+ and a lower probability of A–T. These associations extended to plasma biomarkers, with significant effects for pTau181 in FHS^38,39^ and, importantly, in ADNI for pTau217, which is a blood-based biomarker recently cleared by the FDA for clinical use ^40^. Although modest in sample size, the convergence of genetic liability with this clinically validated biomarker underscores the translational potential of integrating PRS with scalable blood-based assays to identify high-risk individuals even before symptoms emerge, warranting extension in larger, more diverse populations.

The clinical relevance of these biological associations is supported by cognitive performance analyses. AD PRS was associated with poorer memory, executive function, and language scores across ADSP ancestry groups, although these effects were attenuated or absent in smaller cohorts including FHS, KBASE and Caribbean and Native American Hispanics. Trajectory modeling showed that individuals in the highest PRS decile had lower memory performance from age 60 onward, with the gap relative to lower-PRS groups widening with age. Executive function and language showed similar but weaker patterns. These effects were most apparent among individuals who developed AD, with group differences evident before diagnosis, consistent with prior studies linking AD PRS to cognitive performance differences in the prodromal phase ^41–43^. In cognitively normal participants declined slowly and in parallel across PRS groups, though those with high PRS consistently performed worse, particularly in memory. Among individuals with AD, PRS had only modest effects prior to diagnosis and did not predict the rate of decline once the disease was present. Together, these findings suggest that the AD PRS captures enduring differences in cognitive reserve rather than accelerating decline after onset, highlighting its potential utility in identifying subtle, subthreshold cognitive differences that could inform early intervention ^44,45^. Future studies integrating AD PRS with longitudinal cognitive performance, neuroimaging, and biomarker data will be essential to refine individualized risk prediction and guide precision prevention strategies.

Our results also suggest that the impact of aggregate genetic risk on AD biomarker levels and cognitive performance varies by sex. In ADSP, PRS associations with clinical diagnosis were similar in men and women, whereas in FHS stronger effects were seen in women, a difference likely reflecting study design, with FHS capturing incident cases prospectively and ADSP relying on cross-sectional, predominantly prevalent cases where survival or diagnostic bias may obscure sex-related effects^46^. Across both CSF and plasma biomarkers, the association between AD PRS and tau pathology was stronger in women than in men. This effect was most evident for CSF p-tau181 and extended to plasma p-tau181 and p-tau217. Predicted probability curves from AT classification demonstrated a steeper rise in A+T+ probability as PRS increased, further suggests that women may be more sensitive to developing combined amyloid and tau pathology. Although associations with hippocampal volume were modest, the direction and magnitude of effects were generally stronger among women ^47^, though further investigation in larger cohorts is warranted. In cross-sectional analyses, the association of AD PRS with memory performance was more pronounced in women, particularly within the EA group, consistent with a previous study showing significant genetic correlations with 20 memory tests in women compared to only two in men ^48^. We did not detect sex differences in longitudinal memory decline, which may reflect limited power or cognitive trajectory convergence in later disease stages. These observed sex-specific patterns likely reflect a combination of biological and environmental influences, including hormonal changes, X-linked genetic factors, and differences in vascular, metabolic, and lifestyle exposures between men and women ^49–52^.

A key strength of this study is the development and rigorous evaluation of an *APOE* genotype-independent PRS for AD. We constructed the weighted PRS using a systematically tested framework based on PRS-CS, incorporating ancestry-specific summary statistics from non-proxy GWAS spanning large EA cohorts and the largest available non-EA datasets to date. Summation weights were derived from ADGC data to enhance performance across ancestries. The resulting PRS was evaluated in clinically confirmed AD cases from the multi-ancestry ADSP dataset and further validated in the FHS, KBASE, and Rush cohorts, with consistent performance also observed in the multi-ancestry AoU dataset. Our findings from genetic risk modeling with clinical diagnosis, cognitive trajectories, a structural MRI measure of hippocampal volume, and CSF and plasma AD biomarkers demonstrate the utility of an ancestry-aware, *APOE*-independent PRS for individualized risk stratification and provide new insights into the genetic architecture and early manifestations of AD across diverse populations.

Despite these advancements, our study has several notable limitations. Non-European populations were underrepresented in both discovery and validation datasets, and the absence of ancestry-specific GWAS for key groups such as Native American and Caribbean populations may limit the generalizability of some findings. Additionally, reliance on ICD codes for case ascertainment in the MVP, FinnGen, and AoU datasets may have constrained diagnostic accuracy. Several datasets that were used to assess AD-related phenotypes, particularly among non-European ancestry groups, had modest statistical power to detect ancestry- or sex-specific effects. Nevertheless, these findings represent a critical step toward understanding AD across diverse populations and underscore the need for larger and more ancestrally representative datasets in both discovery and testing efforts. Furthermore, while we observed sex differences in the association of the AD PRS with tau pathology and memory performance, the underlying biological mechanisms remain unclear. Future studies incorporating exposures, such as hormone replacement therapy among post-menopausal women, may help elucidate the molecular pathways driving these sex-specific patterns.

In summary, this study demonstrates the utility of an *APOE* genotype-independent ancestry-aware polygenic risk score for AD, with consistent associations across cognitive, imaging, biomarker, and neuropathological features. Our findings highlight the value of integrating genetic risk into early detection and monitoring strategies and underscore the importance of expanding diverse, well-characterized cohorts to advance precision medicine approaches in Alzheimer’s disease.

## Online methods

### Study Participants

#### Alzheimer’s Disease Sequencing Project (ADSP)

We obtained whole genome sequencing (WGS) and phenotypic data included in the ADSP release 4 dataset, obtained from 36,361 individuals drawn from multiple diverse ancestry cohorts. Study protocols for all cohorts were reviewed and approved by the appropriate institutional review boards. Details of these cohorts and the generation and processing of WGS data were previously described in other studies ^53–55^.

#### The Alzheimer’s Disease Genetics Consortium (ADGC)

The Alzheimer’s Disease Genetics Consortium (ADGC) assembled genotype and phenotypic data for > 56,000 individuals from 61 cohorts ^56^. For this analysis, we identified and removed overlapping individuals between the ADGC and the ADSP datasets, as detailed in **Supplementary Note 1**. We also excluded individuals who were included in the GWAS training dataset, including those from the Japanese cohort. We then restricted the analysis to individuals who were less related than third-degree relatives (kinship coefficient < 0.04), resulting in a final dataset of 32,016 unrelated individuals from 28 cohorts and diverse populations: European ancestry (EA, *n* = 27,231), African American (AA, *n* = 4,227), Caribbean Hispanic (CH, *n* = 264), and East Asian (EAS, *n* = 294). Detailed descriptions of the ADGC cohorts, including participant recruitment, diagnostic procedures, SNP imputation and data processing are reported elsewhere ^5,6,56–58^. Further details of the genotyping and quality controls are provided in **Supplementary Note 1**. Study protocols for all cohorts were reviewed and approved by the appropriate institutional review boards

#### Framingham Heart Study (FHS)

The Framingham Heart Study (FHS) is a prospective, multigeneration cohort study that has collected comprehensive data on cardiovascular, neurological, and other disease outcomes, blood chemistry data, and lifestyle risk factors since 1948^59^. The original cohort enrolled a random sample of 5,209 participants from Framingham, Massachusetts who were monitored through biennial exams^60^. In 1971, the offspring cohort, consisting of 5,214 children of the original cohort and their spouses, was added and examined every 4 to 6 years ^61–63^. In 2002, the Third Generation Cohort, comprising 4,095 individuals who are the grandchildren of the Original Cohort, was established ^64^. Participants underwent detailed cognitive, brain MRI, and neurological evaluations, with dementia diagnoses determined through a consensus process including neurologists and neuropsychologists. Surveillance methods for dementia have been published previously ^61,62^ .The study protocol was approved by the Boston University Institutional Review Board, followed the Strengthening the Reporting of Observational Studies in Epidemiology (STROBE) reporting guideline, and was monitored by a National Heart, Lung, and Blood Institute Observational Study Monitoring Board. Additional details on genotyping, imputation, and quality control are provided in **Supplementary Note 2**. *Korean Brain Aging Study for the Early Diagnosis and Prediction of AD (KBASE)* KBASE is a prospective cohort study initiated at Seoul National University in 2014 that was designed following the protocol of the Alzheimer’s Disease Neuroimaging Initiative (ADNI). The cohort includes well-characterized individuals, comprising cognitively normal adults ages 20 to 90 years, as well as older adults diagnosed with mild cognitive impairment (MCI) or AD. Participants underwent longitudinal assessments capturing clinical, cognitive, and lifestyle information. WGS and phenotype data were obtained through the National Institute on Aging Genetics of Alzheimer’s Disease Data Storage Site NIAGADS; https://dss.niagads.org/datasets/ng00067/). Study protocols were approved by the relevant institutional review boards. Additional details on genotyping and quality control procedures are provided in **Supplementary Note 3.**

#### Rush University Medical Center (Rush) cohorts

We utilized data from four Rush cohorts, which are well-characterized, longitudinal, clinical-pathologic studies designed to investigate aging, cognitive decline, and dementia in diverse populations. The Religious Orders Study (ROS) and Rush Memory and Aging Project (MAP) ^65^ are ongoing epidemiologic studies enrolling older individuals without known dementia at baseline, with annual clinical follow-ups and high autopsy rates. The Minority Aging Research Study (MARS)^66^ and African American Core of the Rush Alzheimer’s Disease Center (AAc) ^67^ specifically focus on non-Hispanic Black participants. All four cohorts follow a harmonized study design, ensuring consistency in clinical assessments, neuropathological examinations, and cognitive evaluations across participants. This standardization allows for robust comparisons and integrated analyses of aging and neurodegenerative disease processes. For this study, we analyzed data from 897 Black participants across these cohorts, including 541 from MARS, 214 from AAc, 80 from MAP, and 62 from ROS. Through these harmonized assessments, our study provides a comprehensive and systematic evaluation of aging and neurodegenerative disease in Black individuals. The study is funded by the National Institute on Aging and approved by the Institutional Review Board of Rush University Medical Center. Further details on genotyping, imputation, and quality control are provided in **Supplementary Note 4**

#### All of Us (AoU)

The All of Us (AoU) Research Program is a large, longitudinal study aimed at enrolling over one million U.S. participants, with a focus on diverse populations for studies of the biological, clinical, social, and environmental determinants of health and disease (All of Us Research Program Investigators et al., 2019). Health data are collected from electronic medical records (EMRs) and participant surveys, which are publicly accessible via the program’s website. Eligible participants are U.S. residents aged 18 years or older who provide informed consent. Study protocols were reviewed and approved by the All of Us Institutional Review Board (IRB).

We accessed the AoU version 7 dataset (08/09/2024), which includes whole-genome sequencing (WGS) data from 245,394 individuals. Additional details on WGS and quality control procedures are provided in **Supplementary Note 5**. Alzheimer’s Disease (AD) phenotypes were defined following the Centralized Interactive Phenomics Resource (CIPHER) guidelines, specifically the classification developed by the Million Veteran Program (MVP) Cognitive Decline and Dementia During Aging Working Group^68^. AD cases were identified using ICD-9 code 333.0 and ICD-10 codes G30.1, G30.8, and G30.9. Controls were restricted to individuals aged 65 years or older without a diagnosis of dementia themselves and without a family history of AD, as determined by responses to the “Personal and Family Health History” survey question: “Including yourself, who in your family has had dementia (includes Alzheimer’s, vascular, etc.)?”. The age at diagnosis was recorded as the onset age and the current age was used for controls. After excluding related individuals and controls younger than 65 years to mitigate potential survival bias, 71,549 unrelated participants remained for analysis.

### Whole genome-sequencing (WGS) and quality control in the ADSP dataset

We obtained whole-genome sequencing (WGS) and phenotype Release 4 data from NIAGADS (https://dss.niagads.org/datasets/ng00067/) for ADSP participants. Information about sequencing protocols, variant calling, and quality control (QC) procedures for ADSP data are available elsewhere (https://www.adgenomics.org/data/). We excluded individuals with a diagnosis of MCI and variants with minor allele frequency (MAF) ≤ 0.01 or missing genotype rates > 1% for the purpose of constructing the PRS. Population ancestry was determined from clusters derived by PCA as described in **Supplementary Note 6**. Details for assessing relatedness and population substructure using the GENESIS R package are provided in **Supplementary Note 7**.

### PRS development

AD PRSs were calculated using GWAS summary statistics from datasets non-overlapping with each other or with the testing data set including data from EADB Stage I (excluding UKBB) ^6^, FinnGen^69^, MVP ^7^, and Japanese ^15^ and Korean GWAS ^16^. GWAS results from the relatively small Japanese and South Korean datasets were combined using GWAMA version 2.1 ^70^. We constructed an AD PRS using multiple methods in order to identify the PRS with the best predictive performance across multi-ancestry populations **(1) *<u>PRSice2</u>*:** We utilized the Clumping and Thresholding method in PRSice2 ^71^ following our previous approach ^72^. Ancestry-specific subsets of the ADSP dataset served as the reference panel for estimating linkage disequilibrium (LD) for each ancestry. PRSs were constructed for each GWAS dataset using clumping parameters (R²) of 0.1, 0.2, 0.3, distances of 100 Kb, 250 Kb, 500 Kb, and p-value thresholds ranging from 5×10⁻⁸ to 0.5. The best-performing PRS was identified using the coefficient of variation approach, where the ADSP dataset was split into five random, unrelated subsets, and the PRS minimizing the coefficient of variation across effect sizes was selected. **(2) *<u>LDPred2</u>*:** PRSs were developed using the auto model in the bigsnpr R package version 1.9.11 ^73^ and approximately 1.4 million SNPs from HapMap3+ ^74^. The ADSP WGS dataset served as the reference panel for LD estimation and weight calibration. After computing the weights, PRSs were calculated without clumping parameters using PRSice2. **(3) *<u>PRS-CS</u>:*** PRS-CS is a Bayesian polygenic risk-scoring method that calculates a PRS from GWAS summary statistics. Ancestry appropriate portions of the 1000 Genome projects reference panel, including HapMap3 SNPs, were used for estimating LD^75^ in each of the GWAS datasets. Default PRS-CS parameters were applied and PRSs were computed in the ADSP dataset using PRSice2 without clumping. **(4) *<u>PRS-CSx</u>:*** We employed PRS-CSx software, which integrates summary statistics from multiple ancestries using the default parameters ^76^. Each GWAS was assigned an ancestry-matched reference panel from 1KG. Since PRS-CSx requires a single dataset per ancestry, GWAS results from the EADB and FinnGen European ancestry datasets were combined by meta-analysis using GWAMA v2.1^70^. Each iteration of PRS-CSx yielded three ancestry-specific PRS weights. PRSs were then computed in the ADSP dataset using PRSice2 without applying clumping parameters. To facilitate the comparison of effect sizes across multi-ancestry analyses, each PRS was scaled and centered to have a mean of 0 and a SD of 1 in the total ADSP sample.

### Training of PRS summation weights using ADGC data

We identified and excluded from the ADGC dataset 33,402 individuals who overlapped the ADSP WGS dataset and individuals diagnosed with MCI, yielding an independent sample of 32,016 individuals. PRS weights were derived and computed for all ADGC participants using the same set of SNPs employed in the development of the PRS in the ADSP dataset. This process was conducted separately for each ADGC dataset (PRSice2, PRS-CS, PRS-CSx, and LDPred2). The PRS weights were scaled using the PRS means and SDs derived from the ADSP dataset ^77^. The association of AD with the PRS derived from each GWAS (i.e., EADB, FinnGen, MVP, and EAS) was evaluated for each method using a logistic regression model including covariates for age (either onset of AD symptoms or age at last exam), sex, ancestry, cohort or study (to account for batch effects), the first 10 PCs to adjust for population stratification, and carrier status for the *APOE* ε4 and ε2 alleles. The effect estimates from the PRSs for each GWAS (*w₁* from EADB, *w₂* from FinnGen, *w₃* from MVP, and *w₄* from EAS) were derived separately for each PRS method and used as weights for the respective PRSs. These weighted effect estimates were subsequently used to calculate the PRS summation.

### PRS evaluation in the ADSP dataset

We developed a multi-ancestry PRS by comparing its performance among three strategies used for its construction: (1) **Meta-analysis**: Combining GWAS summary statistics from all ADSP WGS datasets using GWAMA v2.1 (Mägi & Morris, 2010) and creating PRSs using PRSice2 and LDPred2; (2) **Unweighted PRS summation**: PRS₁ + PRS₂ + PRS₃ + PRS₄, applicable across all methods; (3) **Weighted PRS summation**: *w₁ × PRS₁ + w₂ × PRS₂ + w₃ × PRS₃ + w₄ × PRS₄*, applicable across all methods. To simplify the PRS summation combinations into a single PRS weight, we applied the “PRSsum Simple” approach using our previous method (https://github.com/nkurniansyah/PRSsum_Simple)^78^. The best-performing PRS was identified by comparing odds ratios (ORs), p-values and area under the curve (AUC) from tests of its association with AD. We further trained the best-performing PRS by excluding SNPs within a 1 Mb window of *APOE*. Summation weights were derived from the ADGC dataset, and the mean and SD of the final PRS calculated in the ADSP cohort are reported in **Supplementary Table 13.**

### PRS validation in independent datasets

The performance of the AD PRS, constructed excluding the *APOE* region and adjusted for *APOE* ε4 and ε2 carrier status, was validated in the AoU version 7, FHS, KBASE, and Rush cohorts using the same SNP weights from the best-performing PRS in the ADSP evaluation. For weighted PRS summation, we applied PRS combination weights derived from the ADGC dataset to calculate PRS in these cohorts. To ensure consistency across cohorts, the PRS was scaled using the mean and SD from the ADSP dataset ^77^. To evaluate the performance of our multi-ancestry PRS, we benchmarked it against previously published AD-specific variant weights curated in the PGS Catalog ^6,10,18,22,79–94^, as well as a recently published PRS ^12^. Notably, variant weights in all of these studies were derived from GWAS conducted in EA individuals only, many of which included individuals overlapping the ADGC and ADSP datasets. To avoid bias due to sample overlap, benchmarking analyses were restricted to non-European ancestry groups including AA, CH and NAH ADSP participants, and Korean KBASE participants. Published variant weights (excluding the *APOE* region) were downloaded and uniformly applied to individual-level genotype data for ADSP and KBASE participants to construct the AD PRS. Association of the PRS with AD was tested separately in each ancestry group using logistic regression models adjusted for age, sex, dosage of *APOE* ε4 and ε2, and 10 PCs. Study accession numbers and corresponding publications are provided in **Supplementary Tables 6 and 7**.

### AD-related traits

We examined the association between AD PRS and AD-related phenotypes using data from multiple cohorts listed in **Supplementary Table 14**.

#### AD Biomarkers

Cerebrospinal fluid (CSF) measures of amyloid beta (Aß), total Tau, and phosphorylated Tau 181 (pTau181) obtained from 1,389 ADSP participants were standardized and harmonized using Z-score normalization to ensure comparability ^95,96^. pTau181 was measured in plasma from 1,689 FHS participants derived from blood samples obtained at Exam 9 using the Quanterix Simoa pTau-181 Advantage V2 kit on EDTA plasma samples ^39^. Plasma p-tau217 concentrations were measured longitudinally in ADNI participants using the Fujirebio Lumipulse G1200 automated chemiluminescent enzyme immunoassay, a validated platform for high-throughput quantification of AD biomarkers^97,98^.

#### Neuropathological traits

Measures of amyloid plaque severity (CERAD score), neurofibrillary degeneration (Braak stage), and Thal phase were obtained from brain tissue donated by 2,862 ADSP participants. The CERAD score is a semi-quantitative measure of neuritic plaque density, based on established guidelines from the Consortium to Establish a Registry for Alzheimer’s Disease (CERAD) ^99,100^. Braak staging characterizes the distribution of neurofibrillary tangles (NFTs) across six stages: stages I-II indicate NFTs in the entorhinal cortex, III-IV involve limbic regions such as the hippocampus, and V-VI reflect neocortical involvement^101^. It consolidates Braak NFTs stages into four levels of severity, providing a standardized assessment of AD-related tau pathology progression ^102^. Thal phasing describes beta-amyloid deposition across five phases, progressing from neocortical to widespread deposition^103^. The Alzheimer’s Disease Neuropathologic Change (ADNC) score of AD severity determined using modified NIA-Reagan criteria was categorized as low, intermediate, or high, based on CERAD and Braak scores ^102^.

#### Cognitive domain measures

Co-calibrated factor scores for several cognitive domains (executive function, language, and memory) were generated using bifactor structural equation models described elsewhere ^104–106^. These cognitive performance measures are comparable across individuals and datasets regardless of the cognitive battery administered. Scores with a standard error (SE) > 0.6 and individuals younger than 60 years were excluded. The total sample included 59,906 ADSP, 1,762 KBASE, and 8,465 FHS participants with genetic data and harmonized longitudinal cognitive domain scores. In addition, episodic memory and global cognitive function scores were available for 895 Rush participants and were derived using established protocols ^107^.

#### Neuroimaging data

Longitudinal MRI measures of hippocampal volume were obtained for 1,945 ADSP and 970 FHS participants older than 60 years. ADSP T1-weighted scans were processed using the automated FreeSurfer v6.0 pipeline (https://surfer.nmr.mgh.harvard.edu), following standard *recon-all* procedures for cross-sectional data. Detailed processing protocols have been described previously ^108^. Visual quality control followed the ENIGMA protocol (https://enigma.ini.usc.edu/protocols/imaging-protocols/), in which ten trained raters scored cortical surface parcellations on a standardized 3-point scale; scans rated 2 (acceptable) or 3 (full pass) were retained. To harmonize imaging data across scanners, sites, and field strengths, longitudinal ComBat was applied, preserving biological variance due to age, sex, race/ethnicity, and diagnostic status ^109^.

FHS structural weighted MRI scans were processed using FreeSurfer 7.4.1. Participant confidentiality was ensured using MiDeFace, which replaces identifiable facial features with an average face template (“MIDEFACE” label) while minimally altering non-facial structures (https://surfer.nmr.mgh.harvard.edu/fswiki/MiDeFace). As part of the quality assurance (QA) pipeline, all scans were manually inspected to verify the presence of the MIDEFACE label and characteristic facial ridges, indicating that the 3D reconstruction was non-identifiable. Quality was scored on a 1–10 scale; scans rated ≥6 were retained, while those scoring ≤5 were excluded due to inadequate quality or excessive correction requirements. Total hippocampal volume was calculated as the sum of left and right hippocampal volumes. All volumes were normalized to intracranial volume to account for head size differences and subsequently log-transformed for statistical analysis^110,111^.

### Statistical analysis

We evaluated the association of AD PRS with AD using models adjusted for age, sex, ancestry, and 10 PCs, and kinship matrix to account for relatedness among subjects implemented in the GENESIS R package (version 2.16.1). Additional models included *APOE* ε4 and ε2 status. The AoU and Rush cohorts were analyzed using logistic regression models, while generalized linear mixed models with a logit link function accounted for family structure in the FHS dataset. Analyses of Rush and FHS data also adjusted for years of education. The area under the curve (AUC) for each model was calculated using the pROC R package (version 1.18.5)^112^, restricting analyses to unrelated individuals to prevent bias from familial correlations and maintain independent observation. Continuous AD related-traits (e.g. cognitive domain score, biomarker measures and hippocampal volume) were analyzed using linear regression and linear mixed-effects models (LMMs) via the lme4 package (v1.1-29), and ordinal traits (e.g., neuropathological staging) were assessed using ordinal logistic regression with the polr function from the MASS package (v7.3-60). AT classification (A−T−, A+T−, A−T+, A+T+) was modeled using multinomial logistic regression implemented in the VGAM R package (v1.1-6) ^113^, with continuous AD PRS as the predictor, adjusting for age at biomarker measurement, education*, APOE ε4 and ε2 status*, and 10 PCs. Predicted probabilities were estimated across the PRS distribution and averaged over the study population. Cognitive domain results from ADSP and FHS datasets were combined by meta-analysis using fixed-effects models in the meta R package (v8.0-1) ^114^.

To examine the influence of AD PRS on age-related changes in memory performance, we fit generalized additive mixed models (GAMMs), which extend linear mixed models by allowing nonlinear age effects while accounting for random effects, using the **gamm4** R package version 0.2-6 ^115^. Age was centered at the sample median and modeled as a penalized spline with 5 basis functions (k = 5) to allow nonlinear trajectories. Random intercepts and slopes accounted for repeated measures. Analyses included individuals ages 60-80 years with ≥2 cognitive assessments. AD PRS was modeled categorically using empirical quantiles (<10%, 10–50%, 50–90%, >90%), with an age × PRS group interaction term to assess differential trajectories. Marginal predictions and 95% confidence intervals were extracted using the ggeffects R package. To evaluate whether cognitive decline differed by PRS and AD status, we additionally fit LMMs, which allow for interpretable estimation of interaction effects and enable direct comparison of cognitive decline slopes across groups. Due to limited sample size among AD cases, AD PRS was modeled categorically using tertiles (low, moderate, high) to ensure stable group comparisons. The model included fixed effects for age at first visit, dementia diagnosis, PRS group, time since baseline, and their two- and three-way interactions:

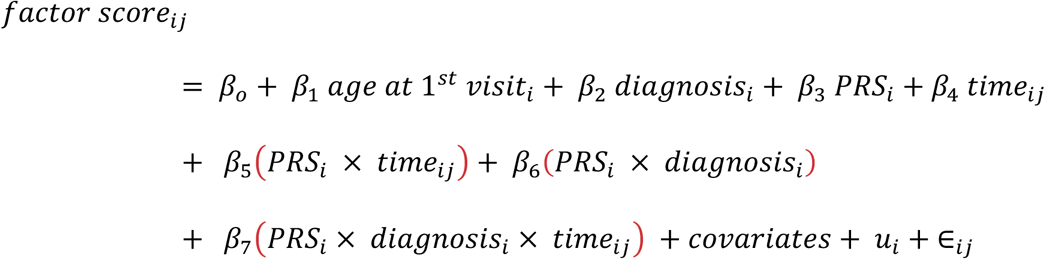

Where *u_i_* denotes participant-specific random intercepts and slopes. Both models adjusted for the same covariates: sex, education, APOE ε2/ε4 status, ancestry, and 10 PCs. Visualization of marginal predictions with 95% confidence intervals was performed using the ggeffects R package.

AD PRS variants and weights will be deposited in the PGS Catalog upon paper acceptance. These data will also be available on the GitHub repository (www.github.com/nkurniansyah/Alzheimer_PRS). Summary statistics for ADRD in the Million Veteran Program African-American dataset have been deposited in dbGaP under study accession phs001672. Summary statistics of the EADB GWAS (without UK biobank) have been deposited to the European Bioinformatics Institute GWAS Catalog (https://www.ebi.ac.uk/gwas/) under accession no. GCST90565439. Summary statistics for AD in the FinnGen European release 10 are available at the FinnGen website(www.finngen.fi/en/access_results). Summary statistics for AD in the Korean GWAS are available from the National Institute on Aging Genetics of Alzheimer’s Disease Storage Site (NIAGADS) (www.niagads.org). Summary statistics for AD in the Japanese GWAS can be accessed at the Database Center for Life Science (DBCLS)(www.humandbs.dbcls.jp/en/). Genotyping and phenotypic data for the Alzheimer’s Disease Sequencing Project (ADSP) are available through the NIAGADS under NG00067 – ADSP Umbrella. Genotyping and phenotypic data for the Alzheimer’s Disease Genetics Consortium can be requested from NIAGADS (www.dss.niagads.org/studies/sa000003). Genotyping and phenotypic data for the Rush cohorts (ROSMAP/MARS/ AAc) can be requested through the Rush Alzheimer’s Disease Center (www.radc.rush.edu). Genotyping and phenotypic data for the Framingham Heart Study (FHS) are available in dbGaP and/or through application to the FHS Research Committee (www.framinghamheartstudy.org/fhs-for-researchers). Data from the NIH All of Us study can be accessed through institutional agreements for researchers who meet eligibility requirements for confidential data access. Interested researchers must complete the registration process via the All of Us Researcher Workbench (www.researchallofus.org/register). For further inquiries, researchers can contact All of Us Researcher Workbench Support at.

## Code availability

Scripts used to perform analyses described in the paper and code to construct multi-ancestry AD PRS are available in the GitHub repository (https://github.com/nkurniansyah/Alzheimer_PRS) and the Zenodo repository (will be provided upon paper acceptance).

## Acknowledgments

This study was supported by National Institute on Aging grants R01-AG048927, U01-AG058654, U01-AG062602, P30-AG072978, U19-AG068753, U01-AG072577, R01-AG080810, and U01-AG082665; National Science Foundation grant DMS/NIGMS-2347698. NK was supported by the National Institute of General Medical Sciences grant T32-GM150533. ML was supported by MVP000 as well as MVP grant (MVP015/VA BLR&D I01BX004192) and the continuation grant (MVP040/BLR&D I01BX005749). This publication does not represent the views of the Department of Veteran Affairs or the United States Government. Acknowledgments for the Alzheimer’s Disease Sequencing Project (ADSP), Alzheimer’s Disease Genetics Consortium (ADGC) and Million Veterans Program (MVP) datasets are provided in **Supplementary Notes 8, 9 and 10.** The Korean Brain Aging Study for the Early Diagnosis and Prediction of Alzheimer’s disease (KBASE) is supported by a grant from Ministry of Science, ICT and Future Planning (Grant No: NRF-2014M3C7A1046042). This work was also supported by the Framingham Heart Study’s National Heart, Lung, and Blood Institute contract N01-HC-25195. We express our gratitude to the staff and participants of the Framingham Heart Study.

The authors thank the participants of the Minority Aging Research Study, the Rush Memory and Aging Project, the Religious Order Study, and the Rush Clinical Core for their invaluable contributions. We thank Charlene Gamboa, MPH; Tracy Colvin, MPH; Tracey Nowakowski, MS; and Theresa Jenkins for study recruitment and coordination, and John Gibbons, MS and Greg Klein for data management, and the staff of the Rush Alzheimer’s Disease Center. This research was supported by National Institute on Aging Grants P30AG10161, P30AG72975, R01AG17917. R01 AG015819, U01 AG072572, U01 AG046152, and R01AG22018 and the Illinois Department of Public Health.

The All of Us Research Program is supported by the National Institutes of Health, Office of the Director: Regional Medical Centers: 1 OT2 OD026549; 1 OT2 OD026554; 1 OT2 OD026557; 1 OT2 OD026556; 1 OT2 OD026550; 1 OT2 OD 026552; 1 OT2 OD026553; 1 OT2 OD026548; 1 OT2 OD026551; 1 OT2 OD026555; IAA #: AOD 16037; Federally Qualified Health Centers: HHSN 263201600085U; Data and Research Center: 5 U2C OD023196; Biobank: 1 U24 OD023121; The Participant Center: U24 OD023176; Participant Technology Systems Center: 1 U24 OD023163; Communications and Engagement: 3 OT2 OD023205; 3 OT2 OD023206; and Community Partners: 1 OT2 OD025277; 3 OT2 OD025315; 1 OT2 OD025337; 1 OT2 OD025276. The All of Us Research Program would not be possible without the partnership of its participants.

We thank Dr. Takeshi Ikeuchi for use of GWAS data from the Japanese Genetic Study Consortium for Alzheimer’s disease (JGSCAD) provided to the Alzheimer’s Disease Genetics Consortium and the European Alzheimer’s Disease Biobank (EADB) for providing GWAS summary statistics. Further information about the EADB is available at https://eadb.eu/. We also acknowledge the participants and investigators of the FinnGen study

## Authors’ contributions

NK, XZ, and LF conceptualized the study. NK performed analyses of ADSP, ADGC, FHS, and All of Us data. ST performed data analysis of RADC data. NK prepared the tables and figures. CZ and JF harmonized and quality controlled the ADGC and ADSP individual-level data. HR performed the neuroimaging analyses in the FHS dataset. RS, RH, VCM, MP, RZ, JMG, and ML provided summary GWAS statistics for the MVP cohort. DYL, KN, and AS provided data for the KBASE cohort. RAV, DAB and LB provided data for the ROSMAP, MARS and RADC cohorts. SR and DG supervised processing of brain imaging data. SM and EHH harmonized ADSP-PHC cognitive data. AJL, AMB, CC, PC, TH, and JM supervised harmonization of ADSP-PHC data. JG, KL, and LAF supervised the acquisition and generation of phenotype and genetic data from the GARD cohort. EM, WB, RM, JLH, MAP, L-SW, GCS and LAF supervised the acquisition and generation of ADGC and ADSP phenotype and genetic data. TFAA, RA, JM, and LAF obtained and processed FHS phenotype data. KLL provided advice and reviewed results of statistical analyses. XZ and LAF supervised the study. NK, XZ, and LAF drafted the manuscript. All authors critically reviewed the manuscript and approved its final version.

