## Supplemental Figures, Tables and Notes for "A multi-ancestry polygenic risk score for Alzheimer disease is associated with cognitive decline, hippocampal atrophy and neuropathological hallmarks in diverse populations"

#### Supplementary Information:

Supplementary Note 3: Korean Brain Aging Study for the Early Diagnosis and Prediction of Alzheimer's Disease (KBASE) genotyping methods and quality control 38

### Supplementary Figures

#### Supplementary Figure 1: Principal components of ancestry in the ADSP dataset

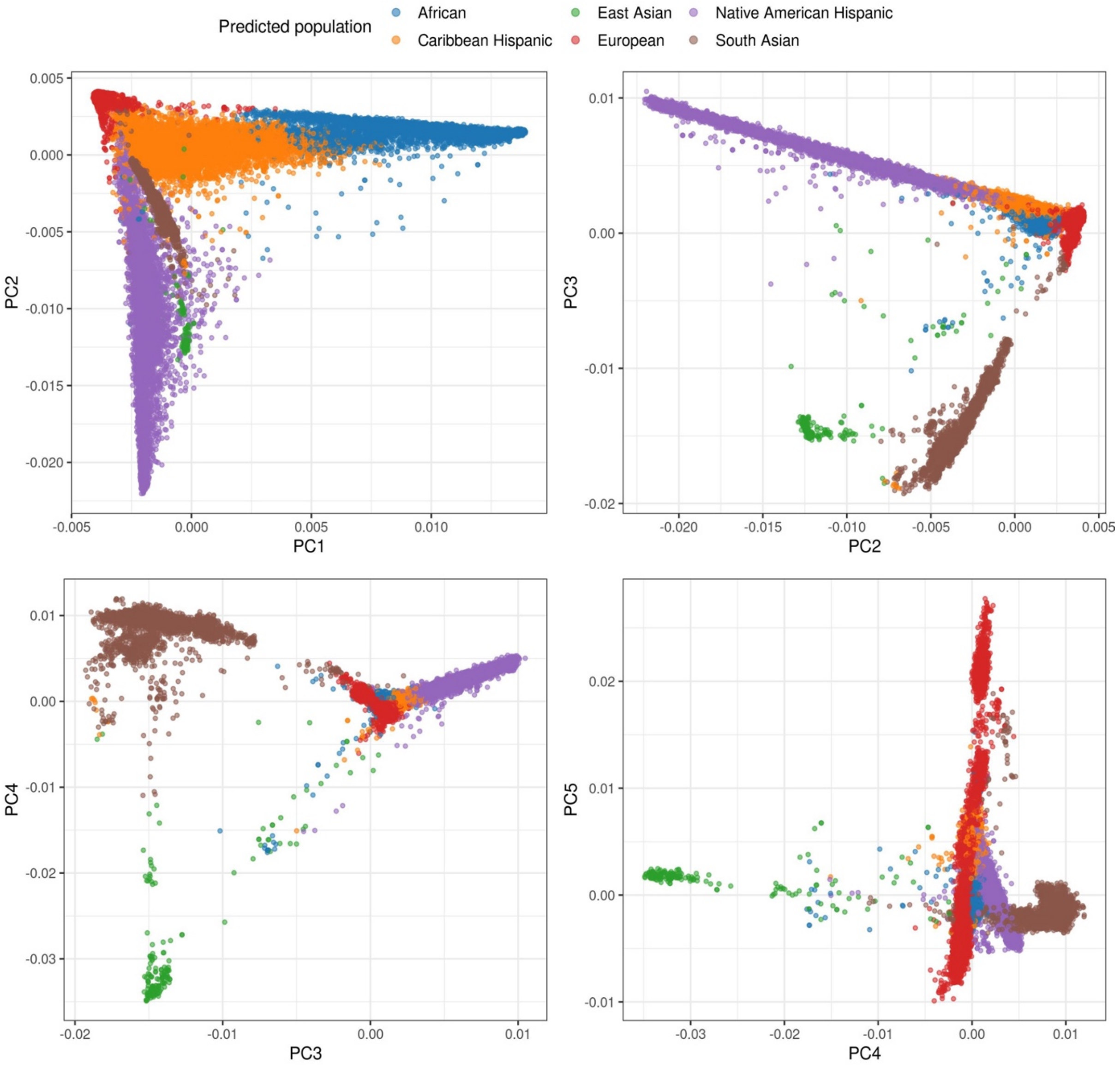

Each plot represents principal components (PC) 1 through 5, with individuals color-coded by predicted population. The x- and y-axes represent the variation captured by each PC pair.

#### Supplementary Figure 2: Ancestry inferences among individuals in the ADSP dataset stratified by population group

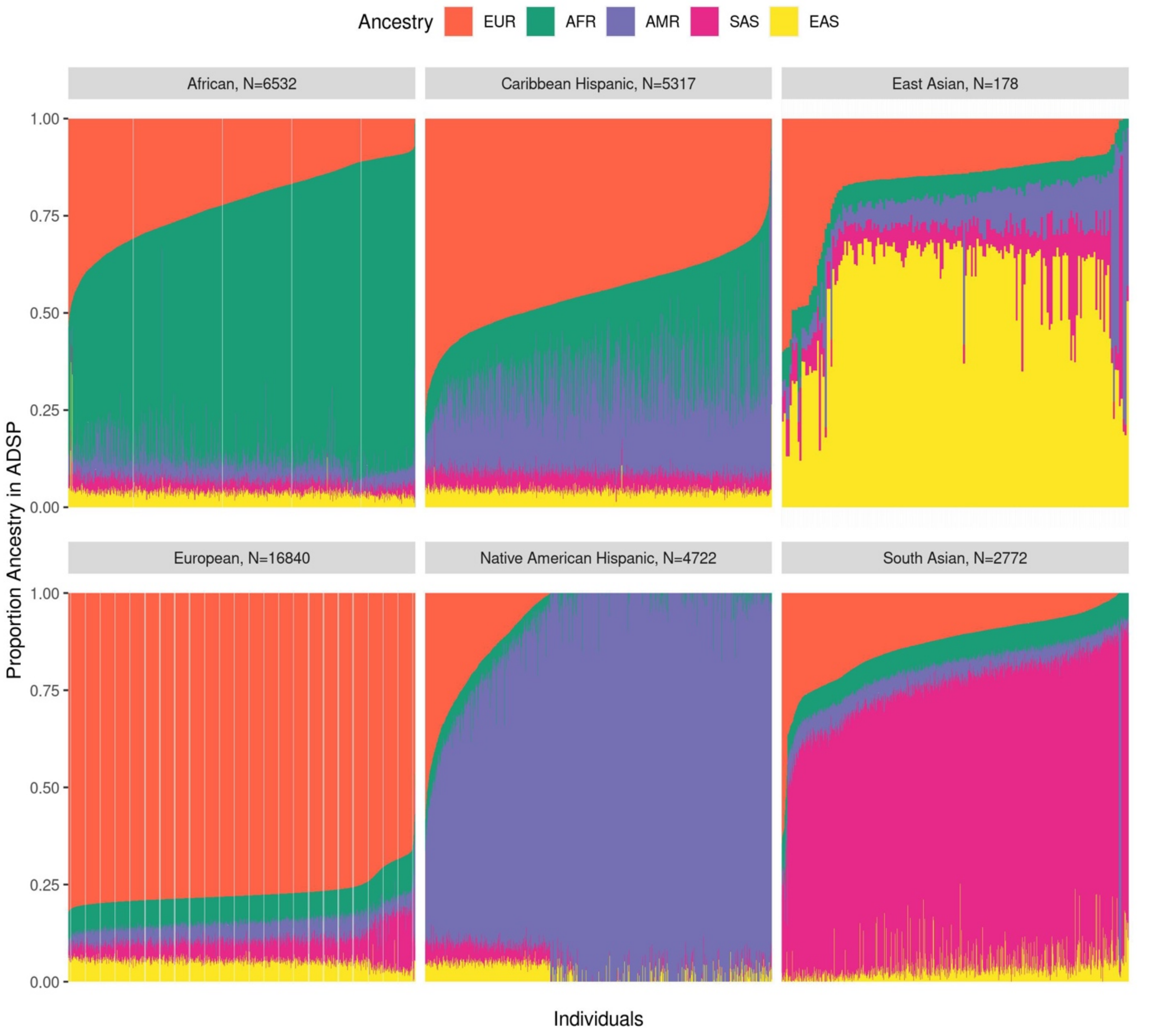

The x-axis represents individuals for each ancestry population and the color-coding highlights the proportion of ancestry inferred for each individual.

#### Supplementary Figure 3: Performance of the multi-ancestry PRS with and without adjustment for *APOE* genotype in the ADSP dataset by method

| | OR [95% CI] | P | AUC |
|---|---|---|---|
| ***Meta analysis*** | | | |
| LDPred2 | 1.46 [1.42;1.51] | $1.73 \times 10^{-107}$ | 0.77 |
| | 1.29 [1.24;1.33] | $1.26 \times 10^{-41}$ | 0.78 |
| PRSice2 | 1.36 [1.32;1.40] | $2.60 \times 10^{-93}$ | 0.77 |
| | 1.23 [1.20;1.27] | $3.47 \times 10^{-40}$ | 0.78 |
| ***Unweighted PRSsum*** | | | |
| PRS-CS | 1.43 [1.39;1.48] | $2.29 \times 10^{-112}$ | 0.78 |
| | 1.20 [1.16;1.24] | $6.45 \times 10^{-24}$ | 0.78 |
| PRS-CSx | 1.40 [1.36;1.45] | $3.10 \times 10^{-91}$ | 0.77 |
| | 1.19 [1.14;1.23] | $2.93 \times 10^{-20}$ | 0.78 |
| LDPred2 | 1.26 [1.22;1.31] | $1.75 \times 10^{-42}$ | 0.77 |
| | 1.16 [1.12;1.20] | $1.06 \times 10^{-16}$ | 0.78 |
| PRSice2 | 1.61 [1.56;1.67] | $3.17 \times 10^{-194}$ | 0.78 |
| | 1.28 [1.23;1.33] | $5.49 \times 10^{-34}$ | 0.78 |
| ***Weighted PRSsum*** | | | |
| PRS-CS | 1.66 [1.60;1.72] | $6.15 \times 10^{-181}$ | 0.78 |
| | 1.36 [1.30;1.41] | $7.16 \times 10^{-48}$ | 0.78 |
| PRS-CSx | 1.61 [1.56;1.66] | $8.51 \times 10^{-185}$ | 0.78 |
| | 1.30 [1.25;1.36] | $2.11 \times 10^{-38}$ | 0.78 |
| LDPred2 | 1.58 [1.52;1.65] | $1.77 \times 10^{-101}$ | 0.77 |
| | 1.36 [1.30;1.42] | $9.46 \times 10^{-41}$ | 0.78 |
| PRSice2 | 1.63 [1.58;1.68] | $5.37 \times 10^{-209}$ | 0.78 |
| | 1.32 [1.27;1.37] | $4.93 \times 10^{-45}$ | 0.78 |

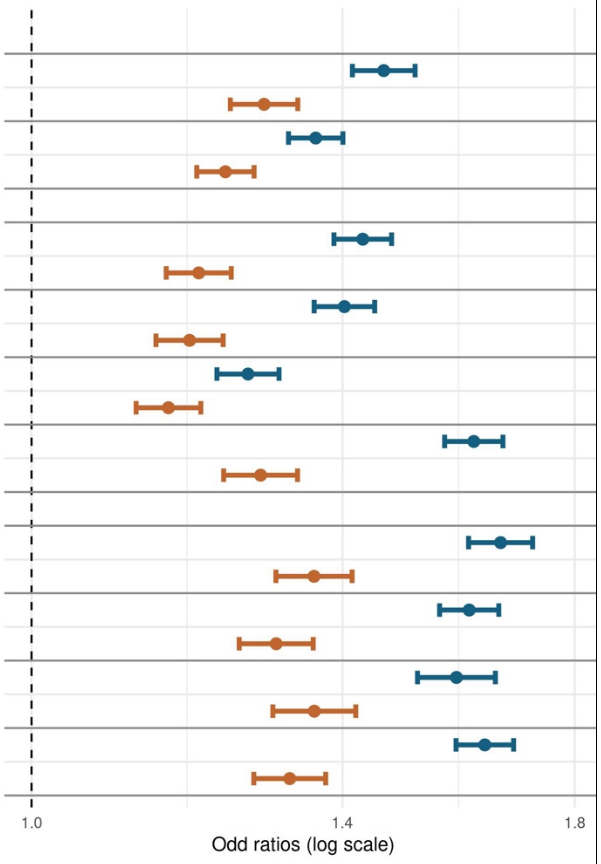

Association of AD with the AD PRS (per 1 SD increase) with AD in entire ADSP dataset (N cases = 10,612; N controls = 16,625). All PRSs include the APOE region. Blue lines represent models without *APOE* genotype adjustment; orange lines represent models adjusted for *APOE-ε4/ε2* status. Multi-ancestry PRSs were constructed using several methods (LDPred2, PRSice2, PRS-CS, and PRS-CSx) and approaches (meta-analysis, unweighted PRSsum, and weighted PRSsum). All analyses were adjusted for age, sex, ancestry group, and first 10 principal components of ancestry. AUC: Area under the curve, CI: confidence interval, OR: Odds ratio; SD: standard deviation

### Supplementary Figure 4: Association of AD with the AD PRS including the *APOE* region in the ADSP dataset by ancestry

| | case | control | OR [95% CI] | P | AUC |
|---|---|---|---|---|---|
| *All* | 10612 | 16265 | 1.66 [1.60;1.72] | $6.15 \times 10^{-181}$ | 0.78 |
| | 10610 | 16264 | 1.36 [1.30;1.41] | $7.16 \times 10^{-48}$ | 0.78 |
| *African* | 1883 | 3373 | 1.45 [1.33;1.57] | $7.37 \times 10^{-19}$ | 0.64 |
| | 1883 | 3373 | 1.14 [1.04;1.25] | $4.81 \times 10^{-03}$ | 0.70 |
| *European* | 6065 | 4763 | 1.81 [1.72;1.90] | $3.55 \times 10^{-115}$ | 0.77 |
| | 6063 | 4762 | 1.49 [1.40;1.59] | $5.52 \times 10^{-34}$ | 0.77 |
| *Caribbean Hispanic* | 1696 | 2302 | 1.53 [1.40;1.67] | $1.23 \times 10^{-20}$ | 0.66 |
| | 1696 | 2302 | 1.35 [1.22;1.49] | $5.21 \times 10^{-09}$ | 0.69 |
| *Native American Hispanic* | 705 | 3282 | 1.39 [1.25;1.56] | $3.19 \times 10^{-09}$ | 0.68 |
| | 705 | 3282 | 1.17 [1.03;1.33] | $1.37 \times 10^{-02}$ | 0.69 |
| *South Asian* | 215 | 2506 | 1.14 [0.92;1.40] | $2.28 \times 10^{-01}$ | 0.66 |
| | 215 | 2506 | 1.00 [0.80;1.26] | $9.71 \times 10^{-01}$ | 0.67 |
| *East Asian* | 48 | 39 | 2.35 [1.05;5.26] | $3.66 \times 10^{-02}$ | 0.77 |
| | 48 | 39 | 2.85 [0.97;8.37] | $5.65 \times 10^{-02}$ | 0.77 |

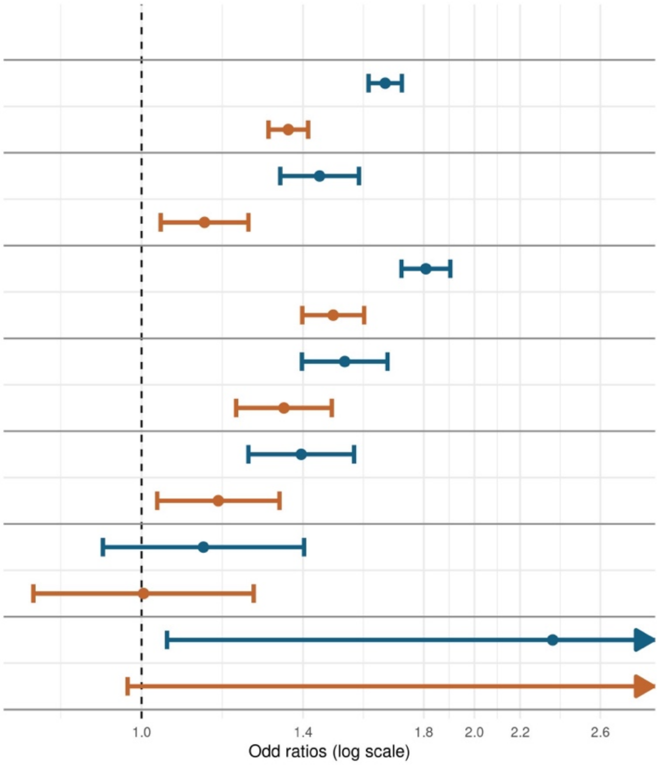

Association of AD with the AD PRS (per 1 SD increase). Models without *APOE-ε4/ε2* status are indicated by blue lines and models including *APOE-ε4/ε2* status are indicated by orange lines. Analyses were adjusted for age, sex and the first 10 principal components of ancestry. We further adjusted for ancestry group in all participants. AUC: Area under the curve, CI: confidence interval, OR: Odds ratio; SD: standard deviation

#### Supplementary Figure 5: Association of AD with the AD PRS excluding the *APOE* region in the ADSP dataset by ancestry

| | case | control | OR [95% CI] | P | AUC |
|---|---|---|---|---|---|
| *All* | 10612 | 16265 | 1.38 [1.34;1.43] | $4.30 \times 10^{-73}$ | 0.77 |
| | 10610 | 16264 | 1.39 [1.34;1.44] | $7.06 \times 10^{-71}$ | 0.78 |
| *African* | 1883 | 3373 | 1.22 [1.13;1.33] | $2.38 \times 10^{-06}$ | 0.61 |
| | 1883 | 3373 | 1.21 [1.11;1.32] | $1.04 \times 10^{-05}$ | 0.70 |
| *European* | 6065 | 4763 | 1.50 [1.43;1.58] | $1.40 \times 10^{-52}$ | 0.75 |
| | 6063 | 4762 | 1.52 [1.44;1.60] | $4.66 \times 10^{-52}$ | 0.77 |
| *Caribbean Hispanic* | 1696 | 2302 | 1.33 [1.22;1.44] | $9.92 \times 10^{-11}$ | 0.64 |
| | 1696 | 2302 | 1.34 [1.22;1.46] | $7.69 \times 10^{-11}$ | 0.69 |
| *Native American Hispanic* | 705 | 3282 | 1.14 [1.02;1.27] | $1.82 \times 10^{-02}$ | 0.67 |
| | 705 | 3282 | 1.14 [1.02;1.27] | $2.40 \times 10^{-02}$ | 0.69 |
| *South Asian* | 215 | 2506 | 0.96 [0.79;1.16] | $6.48 \times 10^{-01}$ | 0.66 |
| | 215 | 2506 | 0.96 [0.79;1.16] | $6.56 \times 10^{-01}$ | 0.67 |
| *East Asian* | 48 | 39 | 2.14 [0.81;5.64] | $1.25 \times 10^{-01}$ | 0.74 |
| | 48 | 39 | 2.05 [0.72;5.88] | $1.81 \times 10^{-01}$ | 0.74 |

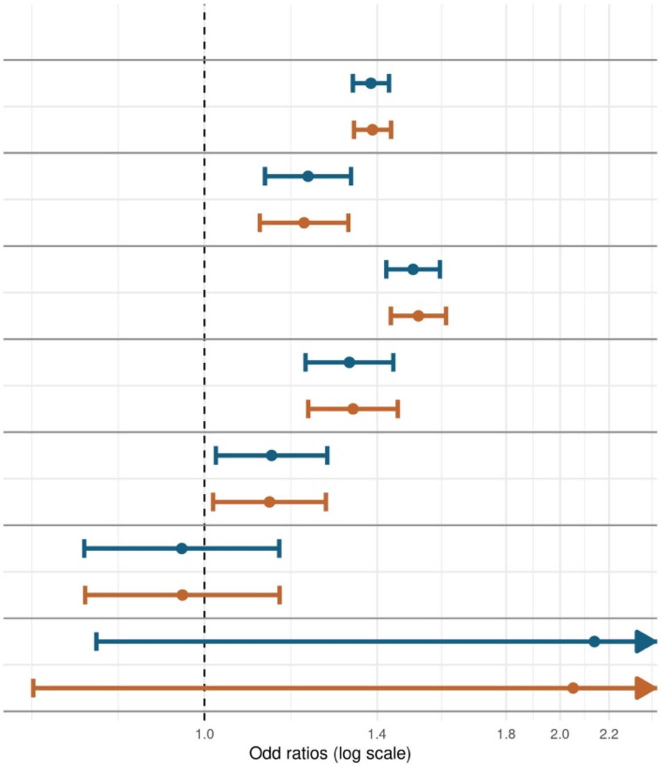

Association of AD with the AD PRS (per 1 SD increase) stratified by sex and ancestry. Models without *APOE-ε4/ε2* status are indicated by blue lines and models including *APOE-ε4/ε2* status are indicated by orange lines. Analyses were adjusted for age, sex, and the first 10 principal components of ancestry. We further adjusted for ancestry group in all participants. AUC: Area under the curve, CI: confidence interval, OR: odds ratio; SD: standard deviation

### Supplementary Figure 6: Association of AD with the AD PRS in the ADSP dataset by sex and ancestry

| Cases | Controls | OR [95% CI] | P | AUC |
|---|---|---|---|---|
| *All* | | | | |
| 10610 | 16264 | 1.39 [1.34;1.44] | $7.06 \times 10^{-71}$ | 0.785 |
| 4151 | 5927 | 1.38 [1.30;1.46] | $2.39 \times 10^{-26}$ | 0.827 |
| 6459 | 10337 | 1.39 [1.33;1.46] | $6.52 \times 10^{-48}$ | 0.759 |
| *African* | | | | |
| 1883 | 3373 | 1.21 [1.11;1.32] | $1.04 \times 10^{-05}$ | 0.704 |
| 528 | 945 | 1.27 [1.07;1.50] | $6.09 \times 10^{-03}$ | 0.717 |
| 1355 | 2428 | 1.20 [1.08;1.33] | $4.03 \times 10^{-04}$ | 0.701 |
| *European* | | | | |
| 6063 | 4762 | 1.52 [1.44;1.60] | $4.66 \times 10^{-52}$ | 0.772 |
| 2690 | 1817 | 1.52 [1.40;1.66] | $1.02 \times 10^{-22}$ | 0.821 |
| 3373 | 2945 | 1.52 [1.42;1.63] | $5.01 \times 10^{-33}$ | 0.756 |
| *Caribbean Hispanic* | | | | |
| 1696 | 2302 | 1.34 [1.22;1.46] | $7.69 \times 10^{-11}$ | 0.690 |
| 570 | 722 | 1.30 [1.12;1.52] | $6.48 \times 10^{-04}$ | 0.726 |
| 1126 | 1580 | 1.35 [1.21;1.50] | $2.30 \times 10^{-08}$ | 0.706 |
| *Native American Hispanic* | | | | |
| 705 | 3282 | 1.14 [1.02;1.27] | $2.40 \times 10^{-02}$ | 0.690 |
| 264 | 1215 | 1.06 [0.89;1.26] | $5.15 \times 10^{-01}$ | 0.652 |
| 441 | 2067 | 1.19 [1.03;1.37] | $1.73 \times 10^{-02}$ | 0.737 |
| *South Asian* | | | | |
| 215 | 2506 | 0.96 [0.50;1.16] | $6.56 \times 10^{-01}$ | 0.669 |
| 79 | 1208 | 0.95 [0.50;1.32] | $7.77 \times 10^{-01}$ | 0.672 |
| 136 | 1298 | 0.94 [0.50;1.20] | $6.19 \times 10^{-01}$ | 0.722 |

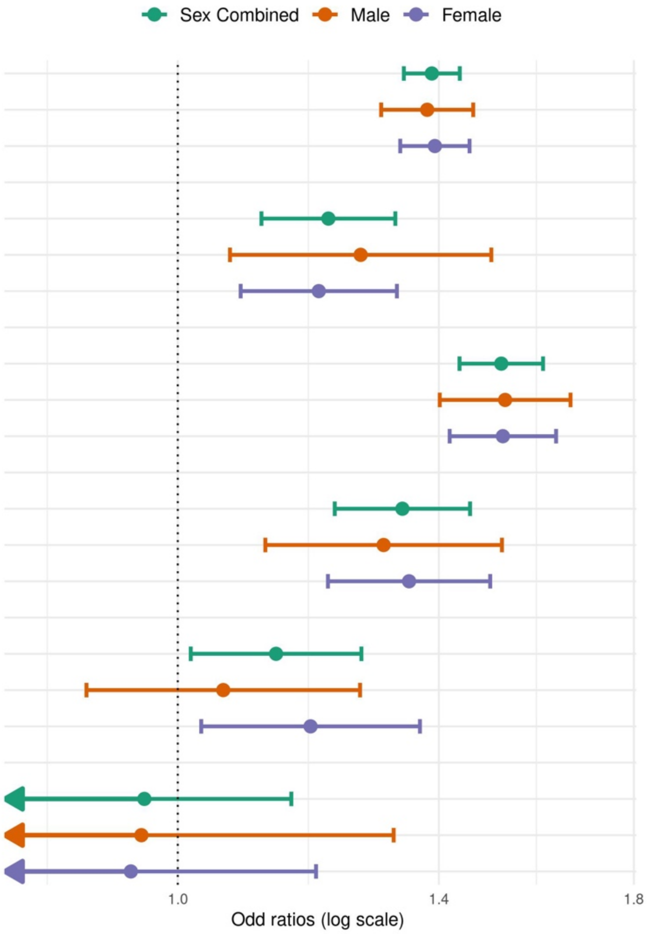

Association of AD with the AD PRS (per 1 SD increase) stratified by sex and ancestry. Analyses were adjusted for age, sex (unless stratified), *APOE-ε4/ε2* status and the first 10 principal components of ancestry. We further adjusted for ancestry group in all participants. AUC: Area under the curve, CI: confidence interval, OR: Odds ratio; SD: standard deviation

### Supplementary Figure 7: Associations of AD with the AD PRS in the ADSP dataset by *APOE* genotype and ancestry

| Ancestry | case | control | OR [95% CI] | P | AUC |
|---|---|---|---|---|---|
| All | 523 | 1609 | 1.16 [1.01;1.34] | $3.30 \times 10^{-02}$ | 0.70 |
| | 4373 | 10299 | 1.37 [1.30;1.43] | $1.63 \times 10^{-36}$ | 0.75 |
| | 4271 | 3733 | 1.45 [1.36;1.55] | $2.53 \times 10^{-31}$ | 0.73 |
| | 1188 | 325 | 1.52 [1.26;1.84] | $1.26 \times 10^{-05}$ | 0.86 |
| African | 167 | 532 | 1.07 [0.81;1.42] | $6.34 \times 10^{-01}$ | 0.62 |
| | 631 | 1632 | 1.23 [1.08;1.41] | $2.33 \times 10^{-03}$ | 0.64 |
| | 767 | 962 | 1.21 [1.05;1.39] | $9.22 \times 10^{-03}$ | 0.58 |
| | 238 | 110 | 1.52 [1.06;2.17] | $2.38 \times 10^{-02}$ | 0.46 |
| European | 200 | 496 | 1.08 [0.86;1.35] | $4.98 \times 10^{-01}$ | 0.70 |
| | 2176 | 2782 | 1.51 [1.40;1.62] | $1.49 \times 10^{-27}$ | 0.71 |
| | 2735 | 1297 | 1.58 [1.45;1.73] | $1.30 \times 10^{-24}$ | 0.71 |
| | 810 | 111 | 1.53 [1.14;2.05] | $4.66 \times 10^{-03}$ | 0.83 |
| Caribbean Hispanic | 119 | 217 | 1.39 [0.99;1.95] | $5.61 \times 10^{-02}$ | 0.69 |
| | 937 | 1549 | 1.33 [1.20;1.49] | $2.12 \times 10^{-07}$ | 0.69 |
| | 514 | 460 | 1.36 [1.14;1.62] | $5.18 \times 10^{-04}$ | 0.53 |
| | 101 | 37 | 1.70 [0.99;2.92] | $5.37 \times 10^{-02}$ | 0.57 |
| Native American Hispanic | 13 | 154 | 0.83 [0.38;1.81] | $6.38 \times 10^{-01}$ | 0.86 |
| | 474 | 2509 | 1.15 [1.01;1.31] | $3.44 \times 10^{-02}$ | 0.66 |
| | 185 | 569 | 1.10 [0.86;1.40] | $4.62 \times 10^{-01}$ | 0.64 |
| | 30 | 32 | 0.88 [0.33;2.34] | $7.95 \times 10^{-01}$ | 0.74 |

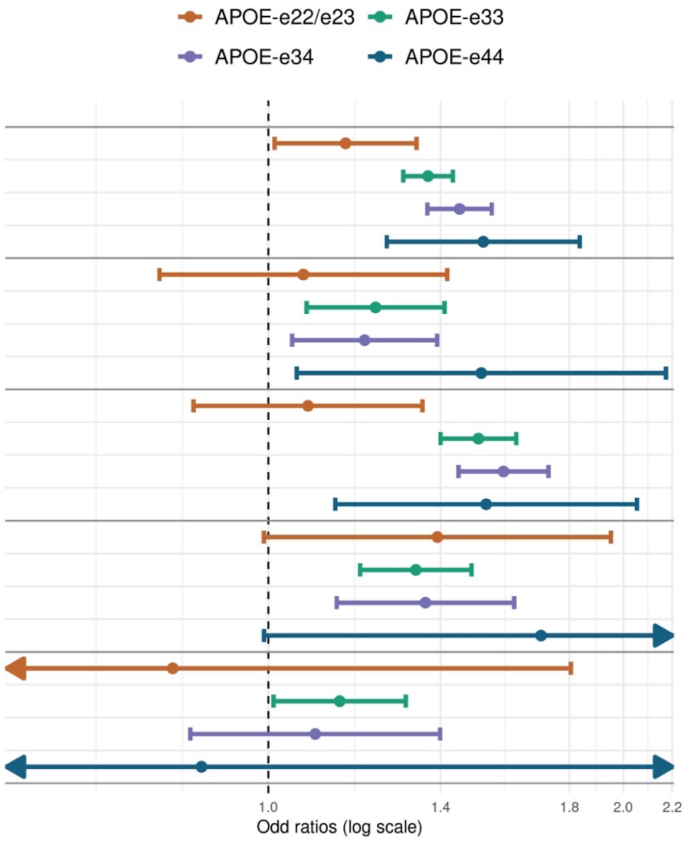

Association of AD with the AD PRS (per 1 SD increase) stratified by APOE genotype and ancestry. Analyses were adjusted for age, sex, and the first 10 principal components of ancestry. We further adjusted for ancestry group in all participants. AUC: Area under the curve, CI: confidence interval, OR: odds ratio; SD: standard deviation

#### Supplementary Figure 8. Association of AD PRS with AD in the FHS and Rush University Medical Center datasets

| Case | Control | OR [95% CI] | P | AUC |
|---|---|---|---|---|
| ***FHS*** | | | | |
| 579 | 3154 | 1.32 [1.17;1.48] | 4.57e−06 | 0.68 |
| 186 | 1478 | 1.15 [0.94;1.41] | 1.65e−01 | 0.67 |
| 393 | 1676 | 1.42 [1.23;1.65] | 2.22e−06 | 0.67 |
| ***RADC*** | | | | |
| 200 | 697 | 1.35 [1.05;1.73] | 1.92e−02 | 0.60 |
| 158 | 541 | 1.28 [0.96;1.69] | 9.34e−02 | 0.69 |
| 42 | 156 | 1.71 [0.96;3.06] | 6.90e−02 | 0.66 |

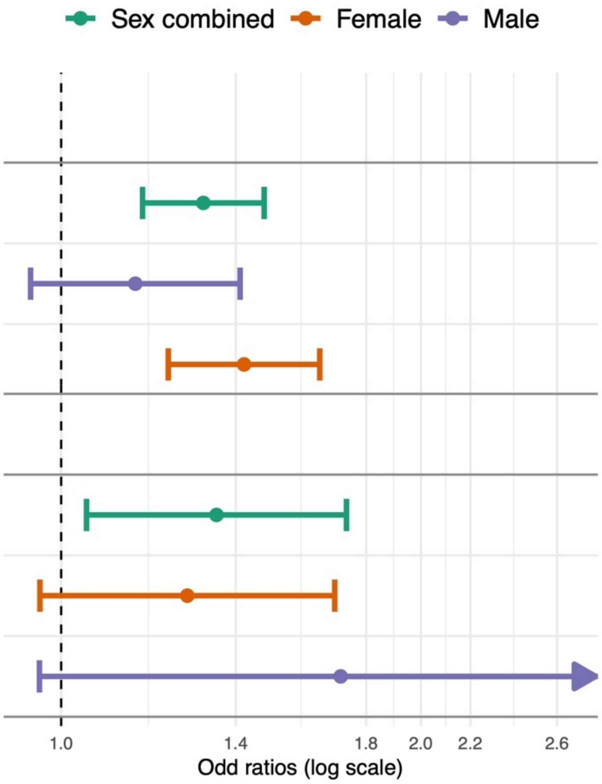

Association of AD with the AD PRS (per 1 SD increase). Analyses were adjusted for age, sex (unless stratified), *APOE-ε4/ε2* carrier status, education and the first 10 principal components of ancestry. In FHS, models were further adjusted for generation cohort and family ID as a random effect. Rush datasets include ROSMAP, MARS and the Rush Alzheimer's Disease Center. AUC: Area under the curve, CI: confidence interval, OR: odds ratio

#### Supplementary Figure 9. Association of AD PRS with plasma biomarkers in the ADNI and FHS datasets

| | N | BETA [95% CI] | P |
|---|---|---|---|
| ***ADNI*** | | | |
| pTau217 | 301 | 0.20 [ 0.05; 0.35] | $8.85 \times 10^{-03}$ |
| | 100 | 0.33 [ 0.10; 0.57] | $7.60 \times 10^{-03}$ |
| | 201 | 0.02 [-0.17; 0.22] | $8.24 \times 10^{-01}$ |
| ***FHS*** | | | |
| pTau181 | 1679 | 0.07 [ 0.01; 0.12] | $1.42 \times 10^{-02}$ |
| | 929 | 0.08 [ 0.01; 0.15] | $2.51 \times 10^{-02}$ |
| | 750 | 0.03 [-0.05; 0.12] | $4.51 \times 10^{-01}$ |

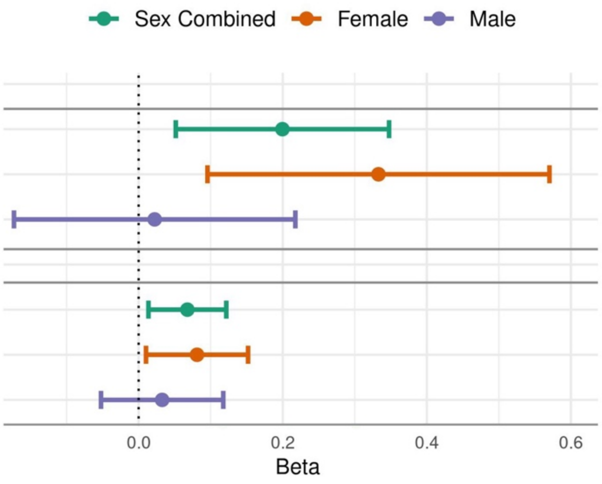

Association of the AD PRS and plasma biomarkers (pTau217 and pTau181) stratified by sex. Beta coefficients represent the change in standardized plasma biomarker level per 1 SD increase in the AD PRS. Analysis models included covariates for age at plasma biomarker measurement, sex (unless stratified), education, *APOE* ε4 status, ancestry group and the first 10 principal components of ancestry. Models in the FHS dataset were further adjusted for family ID as a random effect.

#### Supplementary Figure 10: Association of the AD PRS with neuropathological traits

| | N | OR [95% CI] | P |
|---|---|---|---|
| *CERAD Score* | 2862 | 1.37 [1.25;1.49] | $3.95 \times 10^{-12}$ |
| *Braak Stages* | 2856 | 1.41 [1.30;1.53] | $1.15 \times 10^{-15}$ |
| *Thal Phase* | 1156 | 1.38 [1.20;1.58] | $6.90 \times 10^{-06}$ |
| *ADNC* | 1152 | 1.52 [1.30;1.76] | $6.80 \times 10^{-08}$ |

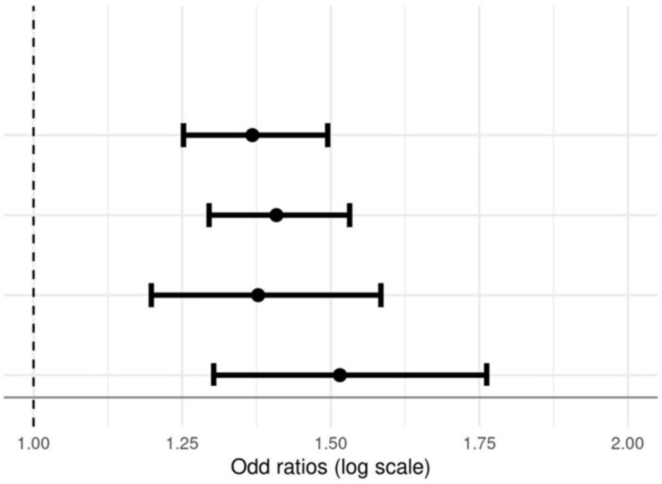

Association between AD PRS and neuropathological traits. Odds ratios represent the change in odds of greater severity per 1 SD increase in AD PRS. Models were adjusted for age at death, sex, education*, APOE ε4/ε2* status, ancestry group, and the first 10 principal components of ancestry. CI, confidence interval; OR, odds ratio.

#### Supplementary Figure 11: Association of the AD PRS with executive function by dataset and ancestry

| | N | BETA [95% CI] | P |
|---|---|---|---|
| ***ADSP*** | | | |
| *All* | 54271 | -0.11 [-0.13;-0.10] | $1.01 \times 10^{-33}$ |
| | 34318 | -0.10 [-0.12;-0.08] | $1.07 \times 10^{-17}$ |
| | 19953 | -0.14 [-0.17;-0.10] | $1.89 \times 10^{-17}$ |
| *African* | 8484 | -0.06 [-0.10;-0.02] | $3.78 \times 10^{-03}$ |
| | 6363 | -0.07 [-0.11;-0.02] | $5.50 \times 10^{-03}$ |
| | 2121 | -0.06 [-0.15; 0.03] | $1.99 \times 10^{-01}$ |
| *European* | 37554 | -0.15 [-0.17;-0.13] | $1.77 \times 10^{-32}$ |
| | 22447 | -0.14 [-0.18;-0.11] | $7.14 \times 10^{-19}$ |
| | 15107 | -0.16 [-0.20;-0.12] | $5.22 \times 10^{-15}$ |
| *Caribbean Hispanic* | 2925 | -0.04 [-0.10; 0.02] | $1.54 \times 10^{-01}$ |
| | 2060 | -0.01 [-0.07; 0.06] | $8.52 \times 10^{-01}$ |
| | 865 | -0.11 [-0.21;-0.02] | $2.34 \times 10^{-02}$ |
| *Native American Hispanic* | 4923 | -0.01 [-0.05; 0.03] | $6.97 \times 10^{-01}$ |
| | 3243 | 0.00 [-0.05; 0.05] | $9.29 \times 10^{-01}$ |
| | 1680 | -0.03 [-0.10; 0.05] | $4.92 \times 10^{-01}$ |
| ***KBASE*** | | | |
| *South Korean* | 1683 | -0.05 [-0.14; 0.03] | $2.35 \times 10^{-01}$ |
| | 931 | 0.00 [-0.12; 0.12] | $9.95 \times 10^{-01}$ |
| | 752 | -0.13 [-0.26; 0.00] | $5.52 \times 10^{-02}$ |
| ***FHS*** | | | |
| *European* | 8114 | -0.02 [-0.04; 0.00] | $3.19 \times 10^{-02}$ |
| | 4634 | -0.04 [-0.07;-0.01] | $2.77 \times 10^{-03}$ |
| | 3480 | 0.00 [-0.03; 0.03] | $9.66 \times 10^{-01}$ |
| ***ADSP+FHS*** | | | |
| *European* | 45668 | -0.07 [-0.09;-0.06] | $1.94 \times 10^{-20}$ |
| | 27081 | -0.08 [-0.10;-0.06] | $1.19 \times 10^{-15}$ |
| | 18587 | -0.06 [-0.09;-0.04] | $7.77 \times 10^{-07}$ |

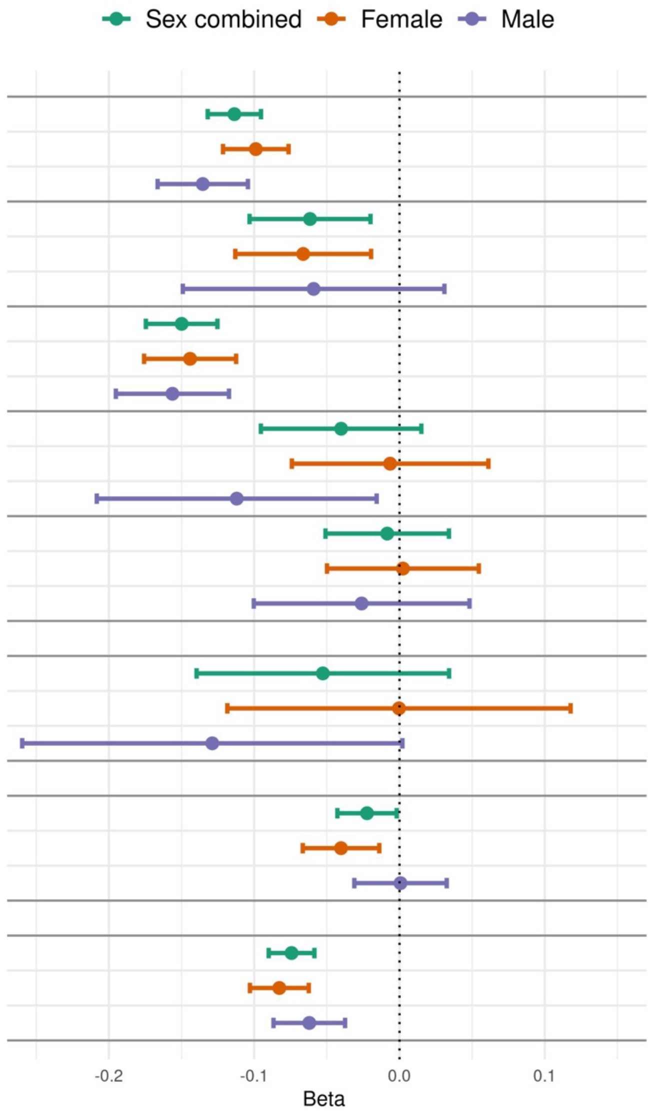

Beta coefficients represent the decrease in standardized memory scores per 1 SD increase in the AD PRS. Analyses were adjusted for age at exam, sex (unless stratified), education, ancestry group (in the combined ADSP dataset), *APOE-ε4/ε2* carrier status, and the first 10 principal components of ancestry. In the FHS dataset, models also included covariates for generation cohort and family ID as random effects. CI: confidence interval, SD: standard deviation

### Supplementary Figure 12: Association of the AD PRS with language score by dataset and ancestry

| | N | BETA [95% CI] | P |
|---|---|---|---|
| ***ADSP*** | | | |
| *All* | 55439 | -0.12 [-0.14;-0.10] | $7.21 \times 10^{-39}$ |
| | 35118 | -0.11 [-0.13;-0.08] | $4.50 \times 10^{-21}$ |
| | 20321 | -0.13 [-0.16;-0.10] | $1.81 \times 10^{-19}$ |
| *African* | 8746 | -0.06 [-0.10;-0.02] | $2.34 \times 10^{-03}$ |
| | 6527 | -0.06 [-0.10;-0.01] | $1.03 \times 10^{-02}$ |
| | 2219 | -0.07 [-0.15; 0.01] | $6.73 \times 10^{-02}$ |
| *European* | 38110 | -0.15 [-0.18;-0.13] | $5.57 \times 10^{-36}$ |
| | 22830 | -0.15 [-0.18;-0.12] | $1.70 \times 10^{-20}$ |
| | 15280 | -0.16 [-0.19;-0.12] | $4.70 \times 10^{-17}$ |
| *Caribbean Hispanic* | 3212 | -0.05 [-0.10; 0.00] | $4.56 \times 10^{-02}$ |
| | 2258 | -0.04 [-0.09; 0.02] | $2.21 \times 10^{-01}$ |
| | 954 | -0.08 [-0.16; 0.01] | $9.44 \times 10^{-02}$ |
| *Native American Hispanic* | 4984 | -0.04 [-0.08; 0.01] | $9.94 \times 10^{-02}$ |
| | 3293 | -0.03 [-0.08; 0.03] | $3.32 \times 10^{-01}$ |
| | 1691 | -0.06 [-0.14; 0.02] | $1.17 \times 10^{-01}$ |
| ***KBASE*** | | | |
| *South Korean* | 1755 | -0.04 [-0.13; 0.04] | $3.28 \times 10^{-01}$ |
| | 976 | 0.01 [-0.11; 0.13] | $8.99 \times 10^{-01}$ |
| | 779 | -0.11 [-0.23; 0.00] | $5.84 \times 10^{-02}$ |
| ***FHS*** | | | |
| *European* | 7431 | -0.02 [-0.04; 0.00] | $9.79 \times 10^{-02}$ |
| | 4289 | -0.04 [-0.07;-0.01] | $7.07 \times 10^{-03}$ |
| | 3142 | 0.02 [-0.02; 0.05] | $3.29 \times 10^{-01}$ |
| ***ADSP+FHS*** | | | |
| *European* | 45541 | -0.08 [-0.09;-0.06] | $3.81 \times 10^{-22}$ |
| | 27119 | -0.09 [-0.11;-0.07] | $3.46 \times 10^{-16}$ |
| | 18422 | -0.06 [-0.09;-0.04] | $4.49 \times 10^{-07}$ |

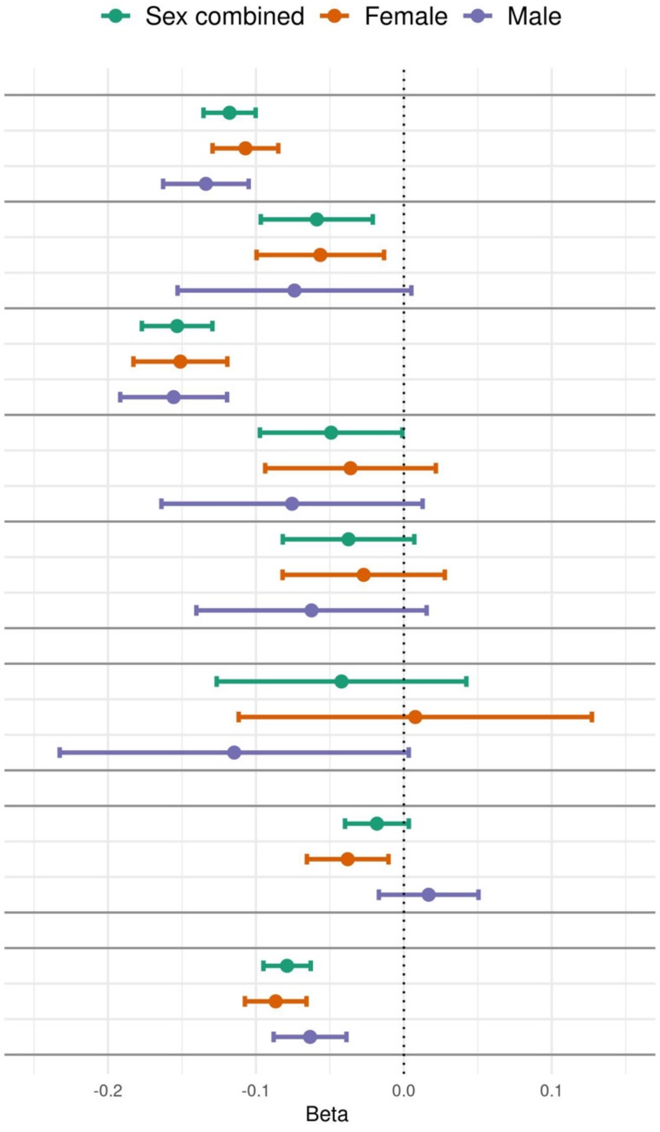

Beta coefficients represent the decrease in standardized memory scores per 1 SD increase in the AD PRS. Analyses were adjusted for age at exam, sex (unless stratified), education, ancestry group (in the combined ADSP dataset), *APOE-ε4/ε2* carrier status, and the first 10 principal components of ancestry. In the FHS dataset, models also included covariates for generation cohort and family ID as random effects. CI: confidence interval, SD: standard deviation

#### Supplementary Figure 13: Association of the AD PRS with episodic memory and global cognition by sex in the Rush University Medical Center datasets

| N | BETA [95% CI] | P |
|---|---|---|
| ***Episodic memory*** | | |
| 895 | -0.20 [-0.31;-0.10] | $1.62 \times 10^{-04}$ |
| 697 | -0.20 [-0.32;-0.08] | $9.06 \times 10^{-04}$ |
| 198 | -0.17 [-0.41; 0.06] | $1.52 \times 10^{-01}$ |
| ***Global cognition*** | | |
| 895 | -0.12 [-0.21;-0.04] | $5.99 \times 10^{-03}$ |
| 697 | -0.13 [-0.23;-0.03] | $9.94 \times 10^{-03}$ |
| 198 | -0.08 [-0.28; 0.11] | $4.06 \times 10^{-01}$ |

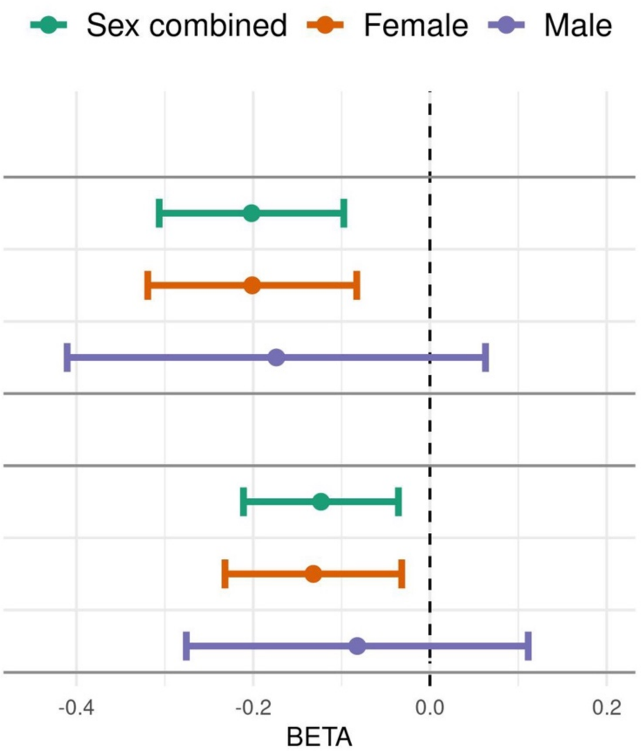

Association of the AD PRS (per 1 SD increase) with standardize global cognition and episodic memory. Analyses were adjusted for age at exam, sex (unless stratified), education, *APOE-ε4/ε2* carrier status, and the first 10 principal components of ancestry. CI: confidence interval

#### Supplementary Figure 14. Age-related trajectory of cognitive performance in the ADSP dataset by AD PRS group and sex

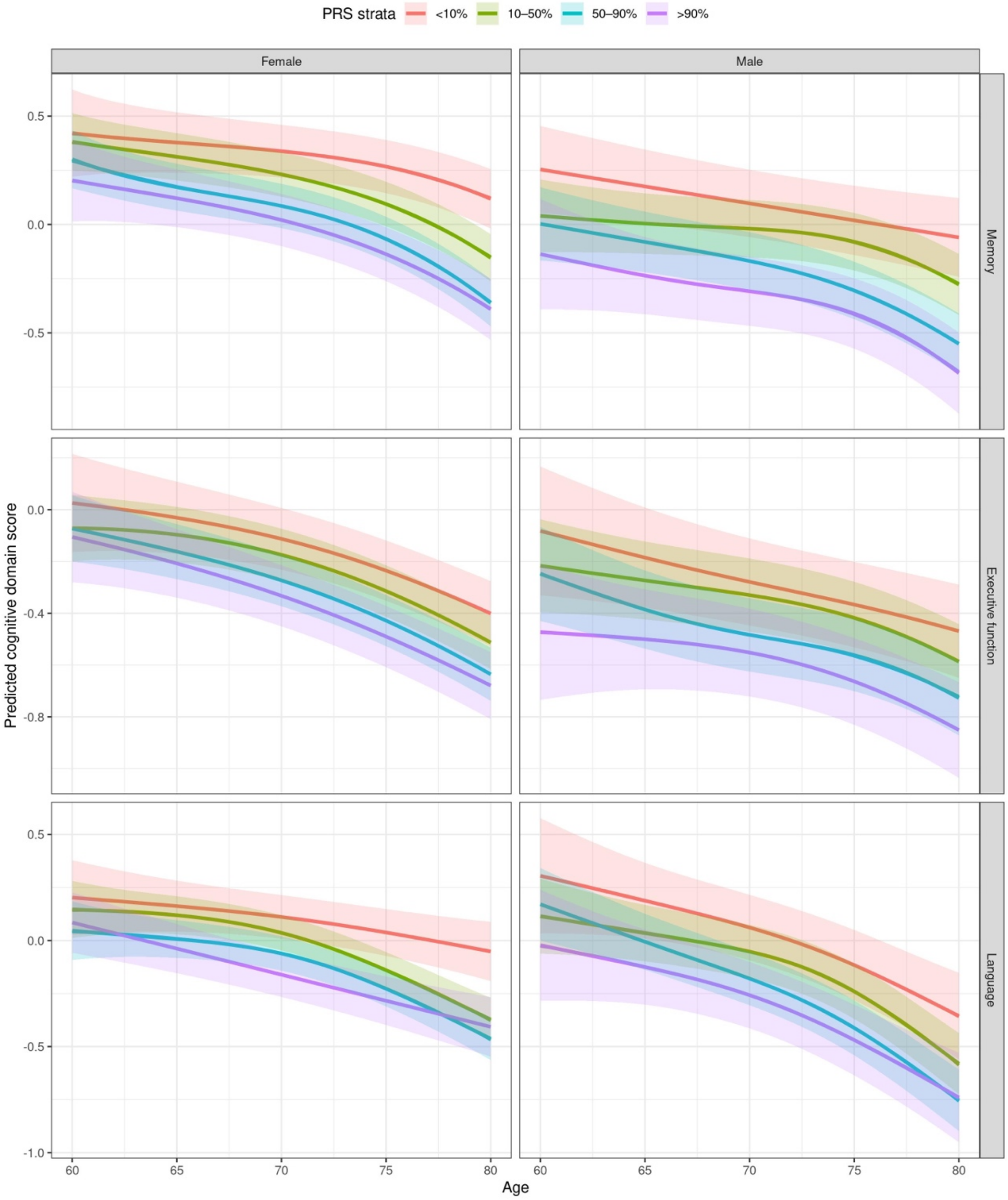

Age-related trajectories among all participants in the ADSP dataset stratified by sex (N female =20,221 and male=11,946) for three cognitive domains (memory, executive function, and language). Solid lines represent predicted cognitive scores by age for PRS groups (<10%, 10–50%, 50–90%, and >90%), with shaded ribbons indicating 95% confidence intervals. Models were adjusted for education, *APOE-ε4/ε2* carrier status, ancestry group, and 10 principal components of ancestry

#### Supplementary Figure 15. Association of the AD PRS with hippocampal volume in the ADSP and FHS datasets

| N | BETA [95% CI] | P |
|---|---|---|
| ***ADSP*** | | |
| 6568 | -0.021 [-0.028;-0.013] | $1.37 \times 10^{-07}$ |
| 3330 | -0.024 [-0.035;-0.014] | $3.14 \times 10^{-06}$ |
| 3238 | -0.016 [-0.028;-0.005] | $6.60 \times 10^{-03}$ |
| ***FHS*** | | |
| 970 | -0.017 [-0.029;-0.005] | $5.23 \times 10^{-03}$ |
| 560 | -0.019 [-0.035;-0.003] | $2.33 \times 10^{-02}$ |
| 410 | -0.017 [-0.036; 0.002] | $7.32 \times 10^{-02}$ |

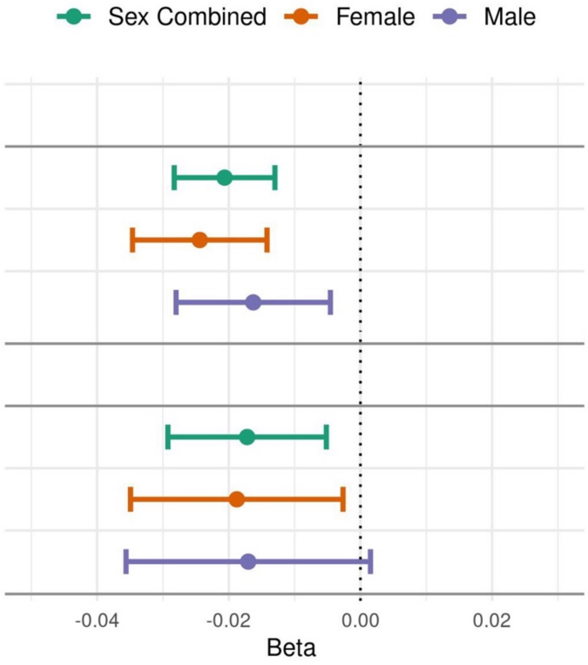

Association of the AD PRS with log-transformed hippocampal volume. Effect sizes represent the change in hippocampal volume (normalized to intracranial volume and log-transformed) per 1 SD higher PRS, adjusted for age at MRI, sex (unless stratified), education, APOE ε4/ε2 status, ancestry group, and the first 10 principal components. FHS models additionally adjusted for generation cohort and included family ID as a random effect. CI, confidence interval.

### Supplementary Tables

#### Supplementary Table 1: Characteristics of ADSP and KBASE participants

| | ADSP | | | | | | KBASE |
|---|---|---|---|---|---|---|---|
| **Ancestry** | African | Caribbean Hispanic | European | Native American Hispanic | South Asian | East Asian | Korean |
| # AD cases/controls | 1883/3373 | 1696/2302 | 6063/4762 | 705/3282 | 215/2506 | 48/39 | 141/284 |
| Sex Female (%) | 3783 (72.0) | 2706 (67.7) | 6318 (58.4) | 2508 (62.9) | 1434 (52.7) | 47 (54.0) | 236 (55.5) |
| Age (mean (SD)) | 74.37 (8.36) | 74.79 (8.03) | 74.81 (10.78) | 68.04 (10.05) | 69.52 (7.28) | 68.56 (9.62) | 70.51 (11.46) |
| *APOE* genotype (%) | | | | | | | |
| ε2/ε2 | 50 (1.0) | 9 (0.2) | 32 (0.3) | 4 (0.1) | 5 (0.2) | 0 (0.0) | 1 (0.2) |
| ε2/ε3 | 649 (12.3) | 327 (8.2) | 664 (6.1) | 163 (4.1) | 218 (8.0) | 0 (0.0) | 39 (9.2) |
| ε2/ε4 | 216 (4.1) | 64 (1.6) | 218 (2.0) | 21 (0.5) | 31 (1.1) | 11 (12.6) | 5 (1.2) |
| ε3/ε3 | 2263 (43.1) | 2486 (62.2) | 4958 (45.8) | 2983 (74.8) | 1940 (71.3) | 2 ( 2.3) | 249 (58.6) |
| ε3/ε4 | 1729 (32.9) | 974 (24.4) | 4032 (37.2) | 754 (18.9) | 491 (18.0) | 42 (48.3) | 112 (26.4) |
| ε4/ε4 | 348 (6.6) | 138 (3.5) | 921 (8.5) | 62 (1.6) | 36 (1.3) | 24 (27.6) | 19 (4.5) |

#### Supplementary Table 2: Characteristics of ADGC participants by ancestry

| | African | East Asian | European | Hispanic |
|---|---|---|---|---|
| # AD cases/controls | 1296 /2391 | 98/ 196 | 12466/14765 | 101/163 |
| # Females (%) | 1299 (30.7) | 103 (35.0) | 11215 (41.2) | 86 (32.6) |
| Mean age (SD) | 77.56 (8.55) | 72.90 (8.98) | 75.00 (7.86) | 70.28 (9.77) |
| APOE genotype (%) | | | | |
| ε2/ε2 | 33 (0.8) | 0 (0.0) | 104 (0.4) | 0 (0.0) |
| ε2/ε3 | 554 (14.0) | 26 (10.8) | 2324 (9.2) | 15 (5.9) |
| ε2/ε4 | 181 (4.6) | 3 (1.2) | 671 (2.7) | 6 (2.3) |
| ε3/ε3 | 1700 (42.9) | 146(60.6) | 12533 (49.8) | 160 (62.5) |
| ε3/ε4 | 1270 (32.0) | 59 (24.5) | 7780 (30.9) | 60 (23.4) |
| ε4/ε4 | 228 (5.7) | 7 (2.9) | 1766 (7.0) | 15 (5.9) |

#### Supplementary Table 3: Characteristics of All of Us participants

| | Overall |
|---|---|
| # AD cases/controls | 501/71056 |
| Mean age (SD) | 74.21 (6.89) |
| # females (%) | 38753 (54.2) |
| Ancestry (%) | |
| African | 12749 (17.8) |
| Amerindian | 7359 (10.3) |
| European | 49649 (69.4) |
| Other | 1800 ( 2.5) |

#### Supplementary Table 4: Characteristics of Rush University Medical Center (ROSMAP, MARS, and RADC) participants

| | Female | Male |
|---|---|---|
| #AD cases/controls | 158/541 | 42/156 |
| Age on set (mean (SD)) | 82.81 (7.12) | 82.45 (6.78) |
| APOE genotype (%) | | |
| ε2/ε2 | 7 (1.0) | 2 (1.0) |
| ε2/ε3 | 87 (12.4) | 40 ( 20.2) |
| ε2/ε4 | 35 (5.0) | 16 (8.1) |
| ε3/ε3 | 351 (50.2) | 87 (43.9) |
| ε3/ε4 | 189 (27.0) | 47 ( 23.7) |
| ε4/ε4 | 30 (4.3) | 6 (3.0) |
| Mean years of education (SD) | 14.85 (3.43) | 14.65 (3.95) |
| Mean global cognition score (SD) | -0.53 (0.96) | -0.69 (0.94) |
| Mean episodic memory score (SD) | -0.32 (1.12) | -0.65 (1.16) |

#### Supplementary Table 5: Characteristics of FHS participants

| | Female | Male |
|---|---|---|
| #AD cases/controls | 403/1839 | 193/1645 |
| Mean age (SD) | 80.53 | 78.04 |
| Mean age at baseline (SD) | 35.32 (9.18) | 35.41 (9.58) |
| Education (%) | | |
| < High school | 225 (10.2) | 181 (10.1) |
| High school | 745 (33.7) | 521 (29.0) |
| Some college | 674 (30.5) | 400 (22.2) |
| College graduate | 566 (25.6) | 697 (38.7) |
| APOE genotype (%) | | |
| ε2/ε2 | 7 (0.3) | 8 (0.5) |
| ε2/ε3 | 276 (13.3) | 178 (10.7) |
| ε2/ε4 | 36 (1.7) | 27 (1.6) |
| ε3/ε3 | 1322 (63.9) | 1124 (67.5) |
| ε3/ε4 | 401 (19.4) | 299 (18.0) |
| ε4/ε4 | 27 (1.3) | 28 (1.7) |

#### Supplementary Table 6: Performance of the AD PRS in ADSP participants by ancestry, constructed using variant weights from other AD PRS studies

| PGS Catalog Identifier | Study | OR [95 % CI] | P-value | Populations |
|---|---|---|---|---|
| | Current Study | 1.21[1,11;1.32] | 1.04E-05 | African |
| | Nicolas A et al. [1] | 1.12 [1.05;1.18] | 2.88E-04 | |
| PGS004863 | Sleiman PM et al. [2] | 1.11 [1.04;1.19] | 2.48E-03 | |
| PGS004898 | Vasiljevic E et al. [3] | 1.10 [1.02;1.17] | 8.00E-03 | |
| PGS004229 | Green RE et al. [4] | 1.01 [0.84;1.21] | 9.30E-01 | |
| PGS003440 | Petrican R et al. [5] | 1.03 [0.97;1.10] | 3.13E-01 | |
| PGS002280 | Bellenguez C et al. [6] | 1.11 [1.05;1.18] | 5.82E-04 | |
| PGS002289 | Zimmerman SC et al. [7] | 1.03 [0.98;1.08] | 2.52E-01 | |
| PGS002731 | Xicota L et al. [8] | 1.06 [0.98;1.15] | 1.41E-01 | |
| PGS003574 | Mukadam N et al. [9] | 1.07 [1.01;1.15] | 3.21E-02 | |
| PGS004600 | Tomassen J et al. [10] | 1.11 [1.05;1.18] | 2.60E-04 | |
| PGS004589 | Jung SH et al. [11] | 1.10 [1.03;1.18] | 6.93E-03 | |
| PGS001775 | Ebenau JL et al. [12] | 1.11 [1.03;1.19] | 4.24E-03 | |
| PGS000811 | Najar J et al. [13] | 1.04 [0.96;1.11] | 3.25E-01 | |
| PGS000898 | de Rojas I et al. [14] | 1.10 [1.03;1.18] | 4.76E-03 | |
| PGS000334 | Zhang Q et al. [15] | 1.07 [1.01;1.14] | 2.75E-02 | |
| PGS000779 | Zhou X et al. [16] | 1.03 [0.97;1.09] | 3.25E-01 | |
| PGS004918 | Lawingco T et al. [17] | 0.99 [0.91;1.07] | 7.70E-01 | |
| PGS002249 | Lourida I et al. [18] | 0.99 [0.76;1.30] | 9.70E-01 | |
| PGS000876 | Leonenko G et al. [19] | 1.10 [1.02;1.19] | 1.21E-02 | |
| PGS000823 | van der Lee SJ et al. [20] | 1.06 [0.99;1.13] | 7.75E-02 | |
| PGS000026 | Desikan RS et al.[21] | 1.07 [0.99;1.16] | 7.27E-02 | |
| PGS000054 | Tosto G et al. [22] | 0.96 [0.91;1.01] | 1.53E-01 | |
| PGS000025 | Chouraki V et al. [23] | 1.07 [1.01;1.15] | 3.37E-02 | |
| | Current Study | 1.34 [1.22;1.46] | 7.69E-11 | Caribbean Hispanic |
| | Nicolas A et al. [1] | 1.23 [1.14;1.32] | 5.69E-08 | |
| PGS004863 | Sleiman PM et al. [2] | 1.16 [1.08;1.25] | 3.17E-05 | |
| PGS004898 | Vasiljevic E et al. [3] | 1.16 [1.08;1.25] | 3.14E-05 | |
| PGS004229 | Green RE et al. [4] | 1.25 [1.03;1.52] | 2.43E-02 | |
| PGS003958 | Sofer T et al. [24] | 1.29 [1.15;1.45] | 1.18E-05 | |
| PGS003440 | Petrican R et al. [5] | 1.11 [1.04;1.20] | 2.88E-03 | |
| PGS002280 | Bellenguez C et al. [6] | 1.23 [1.14;1.32] | 5.73E-08 | |
| PGS002289 | Zimmerman SC et al. [7] | 1.06 [0.99;1.15] | 1.08E-01 | |

| | | | | |
|---|---|---|---|---|
| PGS002731 | Xicota L et al. [8] | 1.16 [1.07;1.25] | 2.70E-04 | |
| PGS003574 | Mukadam N et al. [9] | 1.08 [1.01;1.16] | 3.57E-02 | |
| PGS004600 | Tomassen J et al. [10] | 1.24 [1.15;1.33] | 1.04E-08 | |
| PGS004589 | Jung SH et al. [11] | 1.16 [1.08;1.24] | 6.14E-05 | |
| PGS001775 | Ebenau JL et al. [12] | 1.20 [1.11;1.29] | 1.44E-06 | |
| PGS000811 | Najar J et al. [13] | 1.13 [1.06;1.22] | 4.47E-04 | |
| PGS000898 | de Rojas I et al. [14] | 1.20 [1.11;1.29] | 1.76E-06 | |
| PGS000334 | Zhang Q et al. [15] | 1.12 [1.04;1.21] | 4.05E-03 | |
| PGS000779 | Zhou X et al. [16] | 0.96 [0.90;1.03] | 2.60E-01 | |
| PGS004918 | Lawingco T et al. [17] | 1.07 [1.00;1.15] | 4.61E-02 | |
| PGS002249 | Lourida I et al. [18] | 1.25 [0.96;1.61] | 9.49E-02 | |
| PGS000876 | Leonenko G et al. [19] | 1.18 [1.10;1.27] | 4.37E-06 | |
| PGS000823 | van der Lee SJ et al. [20] | 1.12 [1.04;1.20] | 2.50E-03 | |
| PGS000026 | Desikan RS et al.[21] | 1.14 [1.06;1.23] | 5.59E-04 | |
| PGS000054 | Tosto G et al. [22] | 0.94 [0.86;1.01] | 9.81E-02 | |
| PGS000025 | Chouraki V et al. [23] | 1.13 [1.05;1.21] | 7.61E-04 | |
| | Current Study | 1.14 [1.02;1.27] | 1.40E-02 | Native American Hispanic |
| | Nicolas A et al. [1] | 1.23 [1.12;1.35] | 1.46E-05 | |
| PGS004863 | Sleiman PM et al. [2] | 1.10 [1.01;1.20] | 2.31E-02 | |
| PGS004898 | Vasiljevic E et al. [3] | 1.13 [1.03;1.24] | 6.82E-03 | |
| PGS004229 | Green RE et al. [4] | 1.14 [0.88;1.47] | 3.19E-01 | |
| PGS003958 | Sofer T et al.[24] | 1.11 [0.96;1.28] | 1.75E-01 | |
| PGS003440 | Petrican R et al. [5] | 1.10 [1.00;1.20] | 5.37E-02 | |
| PGS002280 | Bellenguez C et al. [6] | 1.25 [1.13;1.37] | 5.64E-06 | |
| PGS002289 | Zimmerman SC et al. [7] | 1.01 [0.90;1.13] | 9.12E-01 | |
| PGS002731 | Xicota L et al. [8] | 1.11 [1.00;1.22] | 4.11E-02 | |
| PGS003574 | Mukadam N et al. [9] | 1.06 [0.97;1.16] | 1.91E-01 | |
| PGS004600 | Tomassen J et al. [10] | 1.24[1.13;1.37] | 5.11E-06 | |
| PGS004589 | Jung SH et al. [11] | 1.05[0.96;1.15] | 3.15E-01 | |
| PGS001775 | Ebenau JL et al. [12] | 1.15[1.05;1.26] | 2.20E-03 | |
| PGS000811 | Najar J et al. [13] | 1.12[1.03;1.22] | 1.01E-02 | |
| PGS000898 | de Rojas I et al. [14] | 1.15[1.05;1.27] | 2.12E-03 | |
| PGS000334 | Zhang Q et al. [15] | 1.05[0.95;1.17] | 3.57E-01 | |
| PGS000779 | Zhou X et al. [16] | 1.06[0.99;1.14] | 1.13E-01 | |
| PGS004918 | Lawingco T et al. [17] | 1.04[0.94;1.15] | 4.46E-01 | |
| PGS002249 | Lourida I et al. [18] | 1.45[1.02;2.06] | 3.79E-02 | |
| PGS000876 | Leonenko G et al. [19] | 1.10[1.00;1.21] | 4.70E-02 | |
| PGS002289 | van der Lee SJ et al. [20] | 1.01[0.90;1.13] | 9.12E-01 | |
| PGS000026 | Desikan RS et al.[21] | 1.11[1.01;1.21] | 2.87E-02 | |

| | | | |
|---|---|---|---|
| PGS000054 | Tosto G et al. [22] | 0.96[0.84;1.09] | 4.96E-01 |
| PGS000025 | Chouraki V et al. [23] | 1.09[1.00;1.19] | 5.95E-02 |

Associations between PRSs, constructed using variant weights for AD curated from the polygenic score (PGS) catalog and publicly available data, and AD risk were evaluated in the ADSP cohort. Because most of the variant weights used in PRS construction were derived from GWAS that included European ancestry individuals from ADGC and ADSP, analyses were restricted to non-European participants to avoid sample overlap. Analyses were conducted separately in African (1,883 cases, 3,373 controls), Caribbean Hispanic (1,696 cases, 2,302 controls), and Native American Hispanic (705 cases, 3,282 controls) ancestry groups. ORs and 95% CIs represent the association of each PRS (per 1 SD increase) with AD risk, estimated using logistic regression models adjusted for age, sex, *APOE ε4 and ε2* carrier status, and the first ten principal components of ancestry. CI: confidence interval; OR: odds ratio; SD: standard deviation.

### Supplementary Table 7: Performance of the AD PRS in KBASE participants constructed using variants weights from other AD PRS studies

| PGS Catalog Identifier | Study | OR [95 % CI] | P-value |
|---|---|---|---|
| | Current Study | 1.65[1.17;2.34] | 4.72E-03 |
| | Nicolas A et al. [1] | 1.17[0.89;1.52] | 2.55E-01 |
| **PGS004863** | Sleiman PM et al. [2] | 1.32[1.04;1.68] | 2.38E-02 |
| **PGS004898** | Vasiljevic E et al. [3] | 1.12[0.88;1.41] | 3.55E-01 |
| **PGS004229** | Green RE et al. [4] | 1.19[0.63;2.24] | 5.90E-01 |
| **PGS003958** | Sofer T et al. [24] | 1.30[0.88;1.92] | 1.93E-01 |
| **PGS004590** | Lake J et al. [25] | 1.33[0.98;1.80] | 7.01E-02 |
| **PGS003440** | Petrican R et al. [5] | 1.11[0.87;1.41] | 4.18E-01 |
| **PGS002280** | Bellenguez C et al. [6] | 1.17[0.90;1.52] | 2.42E-01 |
| **PGS002289** | Zimmerman SC et al. [7] | 1.05[0.83;1.32] | 6.98E-01 |
| **PGS002731** | Xicota L et al. [8] | 1.15[0.91;1.46] | 2.40E-01 |
| **PGS003574** | Mukadam N et al. [9] | 1.06[0.86;1.30] | 6.14E-01 |
| **PGS004600** | Tomassen J et al. [10] | 1.21[0.93;1.58] | 1.54E-01 |
| **PGS004589** | Jung SH et al. [11] | 1.20[0.91;1.58] | 2.03E-01 |
| **PGS001775** | Ebenau JL et al. [12] | 1.15[0.91;1.46] | 2.54E-01 |
| **PGS000811** | Najar J et al. [13] | 1.25[0.96;1.63] | 9.56E-02 |
| PGS000898 | de Rojas I et al. [14] | 1.15[0.91;1.47] | 2.45E-01 |
| **PGS000334** | Zhang Q et al. [15] | 1.00[0.77;1.30] | 9.96E-01 |
| **PGS000779** | Zhou X et al. [16] | 0.94[0.80;1.11] | 4.52E-01 |
| **PGS004918** | Lawingco T et al. [17] | 0.87[0.57;1.31] | 4.95E-01 |
| **PGS002249** | Lourida I et al. [18] | 0.54[0.20;1.45] | 2.20E-01 |
| **PGS000876** | Leonenko G et al. [19] | 1.14[0.90;1.45] | 2.87E-01 |
| **PGS000823** | van der Lee SJ et al. [20] | 1.09[0.87;1.35] | 4.56E-01 |
| **PGS000026** | Desikan RS et al.[21] | 1.17[0.92;1.50] | 2.09E-01 |
| **PGS000054** | Tosto G et al. [22] | 1.16[0.84;1.61] | 3.67E-01 |
| **PGS000025** | Chouraki V et al. [23] | 1.09[0.88;1.36] | 4.44E-01 |

Associations between PRSs and AD risk were evaluated in the KBASE cohort (141 cases, 284 controls) using scores constructed from previously published variant weights curated from the Polygenic Score (PGS) Catalog, alongside our AD PRS. Odds ratios (ORs) and 95% confidence intervals (CIs) represent the association of each PRS (per 1 SD increase) with AD risk, estimated using logistic regression models adjusted for age, sex, *APOE ε4 and ε2* status, and the first ten principal components of ancestry. CI: confidence interval; OR: odds ratio; SD: standard deviation.

#### Supplementary Table 8: Characteristics of participants with CSF biomarker (ADSP) and plasma biomarker (FHS) data

| | ADSP | | FHS | |
|---|---|---|---|---|
| | Female | Male | Female | Male |
| Number of participants | 701 | 692 | 1097 | 897 |
| Mean age at exam (SD) | 72.92 (7.20) | 74.64 (7.50) | 72.10 (7.28) | 72.05 (6.95) |
| APOE genotype (%) | | | | |
| ε2/ε2 | 0 (0.0) | 2 (0.3) | 2 (0.2) | 3 (0.3) |
| ε2/ε3 | 60 ( 8.6) | 41 ( 5.9) | 158 (14.8) | 108 (12.6) |
| ε2/ε4 | 15 (2.1) | 7 (1.0) | 19 (1.8) | 16 (1.9) |
| ε3/ε3 | 302 (43.1) | 313 (45.2) | 673 (63.2) | 557 (64.9) |
| ε3/ε4 | 265 (37.8) | 242 (35.0) | 201 (18.9) | 157 (18.3) |
| ε4/ε4 | 59 ( 8.4) | 87 (12.6) | 12 (1.1) | 17 (2.0) |
| Ancestry (%) | | | | |
| African | 55 ( 7.8) | 38 ( 5.5) | - | - |
| Caribbean Hispanic | 12 (1.7) | 11 ( 1.6) | - | - |
| East Asian | 5 (0.7) | 7 (1.0) | - | - |
| European | 618 ( 88.2) | 627 ( 90.6) | 1097(55.02) | 897(44.98) |
| Native American Hispanic | 8 (1.1) | 5 (0.7) | - | - |
| South Asian | 3 (0.4) | 4 (0.6) | - | - |
| Education (%) | | | | |
| College | 222 ( 31.9) | 311 ( 45.0) | 360(32.8) | 456(51.1) |
| > High School | 453 ( 65.0) | 357 ( 51.7) | 365(33.3) | 206(23.1) |
| < No High School | 22 ( 3.2) | 23 (3.3) | 371(33.9) | 230(25.8) |
| Mean pTau181 level (SD) | 0.17 (1.03) | 0.15 (1.01) | 2.66 (1.33) | 2.96 (1.52) |
| Mean Tau level (SD) | 0.08 (1.07) | 0.08 (1.00) | - | - |
| Mean Aβ42 level (SD) | -0.01 (0.99) | -0.15 (1.03) | - | - |
| #AD cases/controls | 216/451 | 273/388 | 65/839 | 56/675 |

#### Supplementary Table 9: Characteristics of ADNI participants with longitudinal plasma pTau217 data

| | Women | Men |
|---|---|---|
| **Number** | 100 | 201 |
| **Mean age across all exams (SD)** | 76.94 (6.95) | 79.04 (7.35) |
| **Number of *APOE* ε4 alleles (%)** | | |
| **0** | 41 (41.0) | 81 (40.3) |
| **1** | 49 (49.0) | 95 (47.3) |
| **2** | 10 (10.0) | 25 (12.4) |
| **Education (%)** | | |
| **No High School or Diploma** | 5 ( 5.0) | 0 ( 0.0) |
| **Higher than High School** | 70 (70.0) | 97 (48.3) |
| **Higher Education** | 25 (25.0) | 104 (51.7) |
| **Mean pT217 level across all exams (SD)** | 0.69 (0.72) | 0.46 (0.33) |
| **Number of AD cases (%)** | 51 (51.0) | 105 (52.2) |

#### Supplementary Table 10: Characteristics of ADSP participants with the neuropathological data

| | |
|---|---|
| Number of participants | 3548 |
| Meanage at death (SD) | 82.01 (9.04) |
| Sex Female (%) | 1982 (55.9) |
| Education (%) | |
| < High School | 142 ( 4.9) |
| > High School | 1727 (60.2) |
| College | 1001 (34.9) |
| APOE genotype(%) | |
| ε2/ε2 | 13 (0.4) |
| ε2/ε3 | 190 ( 5.4) |
| ε2/ε4 | 94 ( 2.7) |
| ε3/ε3 | 1515 (42.7) |
| ε3/ε4 | 1406 (39.7) |
| ε4/ε4 | 327 ( 9.2) |
| Ancestry (%) | |
| African | 176 ( 5.0) |
| Caribbean Hispanic | 33 ( 0.9) |
| East Asian | 5 (0.1) |
| European | 3253 (91.8) |
| Native American Hispanic | 71 ( 2.0) |
| South Asian | 4 (0.1) |
| CERAD score (%) | |
| 0 | 564 (15.9) |
| 1 | 349 ( 9.9) |
| 2 | 673 (19.0) |
| 3 | 1955 (55.2) |
| BRAAK stage (%) | |
| 0 | 53 ( 1.5) |
| 1 | 167 ( 4.7) |
| 2 | 303 ( 8.6) |
| 3 | 392 (11.1) |
| 4 | 592 (16.8) |
| 5 | 847 (24.1) |
| 6 | 1163 (33.1) |

| | |
|---|---|
| Thal phase (%) | |
| 0 | 84 ( 7.2) |
| 1 | 67 ( 5.7) |
| 2 | 60 ( 5.1) |
| 3 | 129 (11.0) |
| 4 | 219 (18.8) |
| 5 | 609 (52.1) |
| ADNC (%) | |
| 0 | 84 ( 7.2) |
| 1 | 159 (13.7) |
| 2 | 238 (20.4) |
| 3 | 683 (58.7) |

#### Supplementary Table 11: Characteristics of ADSP, KBASE, and FHS participants with cognitive domain score data

| | ADSP | | | | | | FHS | KBASE |
|---|---|---|---|---|---|---|---|---|
| | African | Caribbean Hispanic | East Asian | European | Native American Hispanic | South Asian | European | Korean |
| **Number of participants** | 9000 | 3426 | 251 | 39454 | 5084 | 51 | 9336 | 1838 |
| **Unique N** | 2108 | 1117 | 55 | 7344 | 1288 | 46 | 4158 | 485 |
| **Sex= Female(%)** | 1540 (73.1) | 771(69.0) | 31 (56.4) | 4280 (58.3) | 853 (66.2) | 27 (58.7) | 2389(54.2) | 282(58.1) |
| **Mean age across all cognitive exams (SD)** | 76.38 (7.55) | 76.22 (7.64) | 76.29 (8.59) | 77.94 (7.81) | 73.58 (8.01) | 74.40 (8.79) | 74.95(9.71) | 71.75(7.17) |
| **Mean age at baseline cognitive exam (SD)** | 73.00 (7.38) | 72.67 (7.07) | 71.36 (8.14) | 73.90 (8.21) | 70.99 (7.86) | 71.71 (7.96) | 69.32(8.17) | 73.62(7.19) |
| **Mean age at last cognitive exam (SD)** | 78.80 (7.60) | 78.73 (7.71) | 76.68 (9.19) | 80.03 (7.85) | 75.18 (8.22) | 76.38 (7.71) | 78.87(8.68) | 74.88(7.43) |
| **Education (%)** | | | | | | | | |
| **< High School** | 502 (24.1) | 607(57.3) | 1 (1.9) | 233( 3.2) | 495 (39.6) | 10 (22.2) | 1475(35.5) | 185(39.8) |
| **> High School** | 1129 (54.3) | 338(31.9) | 29 (53.7) | 4234 (57.8) | 654 (52.3) | 20 (44.4) | 1110(26.7) | 237(51.0) |
| **College** | 450 (21.6) | 114(10.8) | 24 (44.4) | 2852 (39.0) | 102 ( 8.2) | 15 (33.3) | 1569(37.8) | 43(92) |
| **APOE genotype (%)** | | | | | | | | |
| **ε2/ε2** | 13 (0.6) | 2 (0.2) | 0 (0.0) | 21( 0.3) | 1 (0.1) | 0 (0.0) | 22(0.5) | 2 (0.4) |
| **ε2/ε3** | 247 (11.7) | 109( 9.8) | 4 (7.3) | 432( 5.9) | 70 ( 5.4) | 5 (10.9) | 486(12.0) | 39(8.0) |
| **ε2/ε4** | 94( 4.5) | 20 (1.8) | 0 (0.0) | 126( 1.7) | 9 (0.7) | 0 (0.0) | 84( 2.1) | 6 (1.2) |
| **ε3/ε3** | 939 (44.5) | 673(60.3) | 29 (52.7) | 3585 (48.8) | 905 (70.3) | 28 (60.9) | 2599(64.4) | 294(60.0) |
| **ε3/ε4** | 679 (32.2) | 276(24.7) | 26 (29.1) | 2567 (35.0) | 269 (20.9) | 13 (28.3) | 768(19.0) | 124(25.6) |
| **ε4/ε4** | 136( 6.5) | 37( 3.3) | 6 (10.9) | 613( 8.3) | 34( 2.6) | 0 (0.0) | 75(1.9) | 20 (4.1) |
| **Man memory score across all exams (SD)** | 0.22 (0.84) | -0.03 (0.86) | -0.30 (0.94) | 0.28 (1.02) | 0.23 (0.87) | -0.14 (0.94) | 0.29(0.64) | -0.16(0.89) |
| **Mean memory score at baseline (SD)** | 0.19 (0.72) | 0.07 (0.73) | -0.08 (0.81) | 0.22 (0.84) | 0.14 0.76) | 0.07 (0.78) | 0.42(0.53) | -0.18 (0.82) |
| **Mean memory score at last exam (SD)** | -0.01 (1.00) | -0.20 (0.96) | -0.70 (1.16) | -0.15 (1.19) | 0.12 (1.00) | -0.01 (1.00) | 0.16(0.72) | -0.30 (0.98) |

| | | | | | | | | |
|---|---|---|---|---|---|---|---|---|
| **Mean language score across all exams (SD)** | 0.06 (0.78) | -0.37 (0.77) | -0.17 (0.82) | 0.36 (0.90) | -0.15 (0.74) | -0.21 (0.75) | 0.67(0.17) | 0.14 (0.81) |
| **Mean language score at baseline (SD)** | 0.06 (0.72) | -0.33 (0.70) | 0.00 (0.79) | 0.38 (0.77) | -0.19 (0.68) | -0.11 (0.59) | 0.27(0.61) | 0.18 (0.80) |
| **Mean language score at last exam (SD)** | -0.15 (0.87) | -0.51 (0.84) | -0.51 (1.08) | -0.02 (1.05) | -0.27 (0.83) | -0.31 (0.92) | 0.06(0.74) | 0.001 (0.91) |
| **Mean executive function score across all exams (SD)** | -0.09 (0.86) | -0.53 (0.87) | -0.12 (0.77) | 0.35 (0.88) | -0.35 (0.79) | -0.10 (0.81) | -0.02(0.70) | 0.33 (0.87) |
| **Mean executive function score at baseline (SD)** | -0.10 (0.83) | -0.52 (0.84) | 0.19 (0.71) | 0.38 (0.82) | -0.38 (0.75) | -0.17 (0.90) | 0.16(0.62) | 0.16 (0.80) |
| **Mean executive function at last exam (SD)** | -0.32 (0.91) | -0.63 (0.91) | -0.57 (1.04) | -0.05 (1.02) | -0.47 (0.85) | -0.38 (0.83) | -018(0.73) | 0.23 (0.93) |
| **# AD (%)** | 462 (22.5) | 194(17.8) | 20 (37) | 2141 (30.2) | 259 (20.7) | 15 (33.3) | 489(19.0) | 88 (18.2) |

#### Supplementary Table 12: Characteristics of ADSP and FHS participants with longitudinal cognitive and hippocampal volume data

| | ADSP | | FHS | |
|---|---|---|---|---|
| ` | Female | Male | Female | Male |
| **Number of participants** | 3,334 | 3,241 | 570 | 419 |
| **Mean age across all cognitive exams (SD)** | 74.75 (7.22) | 75.93 (6.94) | 72.68 (8.27) | 71.20 (7.70) |
| **Ancestry (%)** | | | | |
| **African** | 312 ( 9.4) | 92 ( 2.8) | | |
| **Caribbean Hispanics** | 70 ( 2.1) | 52 ( 1.6) | | |
| **East Asian** | 30 ( 0.9) | 41 ( 1.3) | | |
| **European** | 2,794 (83.8) | 2,992 (92.3) | 570 (100) | 419(100) |
| **Native American Hispanics** | 121 ( 3.6) | 44 ( 1.4) | | |
| **South Asian** | 7 ( 0.2) | 20 ( 0.6) | | |
| **Education (%)** | | | | |
| **No High School or Diploma** | 156 ( 4.7) | 92 ( 2.8) | 215 (37.7) | 127 (30.3) |
| **Higher than High School** | 1,991 (59.8) | 1,565 (48.3) | 193 (33.9) | 102 (24.3) |
| **Higher Education** | 1,183 (35.5) | 1,581 (48.8) | 162 (28.4) | 190 (45.3) |
| **APOE genotype (%)** | | | | |
| **ε2/ε2** | 4 ( 0.1) | 0 ( 0.0) | 2 (0.2) | 1 (0.1) |
| **ε2/ε3** | 306 ( 9.2) | 224 ( 6.9) | 70 (12.5) | 47 (11.5) |
| **ε2/ε4** | 88 ( 2.6) | 62 ( 1.9) | 8 (1.4) | 8 (2.0_ |
| **ε3/ε3** | 1,578 (47.3) | 1,546 (47.7) | 392 (70.0) | 293 (71.5) |
| **ε3/ε4** | 1,121 (33.6) | 1,109 (34.2) | 84 (15.0) | 51 (12.4) |
| **ε4/ε4** | 237 ( 7.1) | 300 ( 9.3) | 4 (0.7) | 10 (2.4) |
| **Mean hippocampal volume across all MRI exams (SD) *** | -5.37 (0.17) | -5.45 (0.18) | -5.33 (0.15) | -5.41 (0.14) |

* values were log-transformed

#### Supplementary Table 13: PRS-CS weighting summary used in AD PRS construction (excluding the APOE region) in the ADSP cohort.

| Study | Mean | SD | PRS summation weight |
|---|---|---|---|
| FinnGen | 4.35E-08 | 4.30E-08 | 0.12 |
| EADB | 2.64E-07 | 1.20E-07 | 0.29 |
| Asian | 1.50E-07 | 2.53E-07 | 0.04 |
| MVP | -1.20E-06 | 6.47E-07 | 0.01 |
| Final PRS summation | 3.02E-17 | 3.50E-01 | |

#### Supplementary Table 14: Cohorts contributing AD endophenotype data to AD PRS association analyses

| AD related traits | Cohorts |
|---|---|
| **Cognitive domain scores** | ACT, ADNI, EFIGA, Knight ADRC, ROSMAP, NACC, NIA-LOAD, WHICAP, WRAP, KBASE, FHS Gen 1-3 |
| **Episodic memory and global cognition scores** | Rush (ROSMAP, MARS, RADC) |
| **CSF biomarkers** | ADNI, NACC, Knight ADRC, NIA-AD FBS |
| **Plasma p-tau181** | FHS Gen 2 |
| **Plasma p-tau217** | ADNI |
| **Neuropathological data** | ROSMAP, NACC, NIA-LOAD |
| **Hippocampal volume** | FHS Gen1-3, Knight ADRC, ROSMAP, NACC, WHICAP, WRAP |

### Supplementary Notes

#### Supplementary Note 1: Alzheimer's Disease Genetics Consortium (ADGC) quality control of genotype data

To ensure sample independence between ADGC and ADSP datasets, we first curated a set of informative variants for relatedness inference using publicly available markers from https://github.com/brentp/somalier. Genotype data from ADSP whole-genome sequencing (WGS) and ADGC arrays were merged using this variant set. Variants with >10% missingness were excluded. We then used the using KING version 2.3.1 [26] (--duplicate option) to identify genetically identical pairs. Duplicate samples were removed within each dataset (ADSP WGS and ADGC), and additional duplicated individuals were excluded between ADSP and ADGC to ensure non-overlapping samples in downstream analyses.

Following sample-level quality control, genotype data from the ADGC were processed separately by ancestry group. We first ensured sample independence (as described above), then filtered imputed genotype data in Variant Call Format (VCF) using **BCFtools v1.21** [27]retaining only single nucleotide polymorphisms (SNPs) with high imputation quality (INFO score > 0.8) and those passing initial variant-level quality filters. The filtered VCFs were converted to PLINK binary format (BED) using **PLINK v1.9** [28]. and individual BED files were subsequently merged by ancestry group. Variants deviating from Hardy–Weinberg equilibrium ($P < 1\times10^{-6}$) were excluded within each ancestry group. After merging across ancestries, we removed variants with a minor allele frequency (MAF) < 0.01, SNPs with > 2% missing genotypes, and individuals with > 2% missingness. Population structure was controlled by computing principal components and relatedness matrices using the same approach applied in the ADSP dataset (see **Supplementary Note 6**).

#### Supplementary Note 2: Framingham Heart Study genotyping methods and quality control

In the 1990s and early 2000s, DNA samples were collected from participants across the three generations of the Framingham Heart Study (FHS) for genetic research, with all individuals providing consent for genotyping. In 2007, the FHS initiated genotyping for the NHLBI-funded Single Nucleotide Polymorphism (SNP)-Health Association Resource (SHARe) project. This project utilized approximately 550,000 SNPs, employing Affymetrix 250K Nsp and 250K Sty mapping arrays, along with an Affymetrix 50K gene-centered supplemental array, in 9,274 participants from the three generations, including over 1,500 families. Individuals who did not meet the QC criteria—such as a call rate below 97%, extreme heterozygosity, or a high Mendelian error rate—were excluded from the analysis[29]. Imputation was conducted on the Michigan Imputation Server using the TOPMed reference panel. SNPs included in the imputation passed stringent criteria: a call rate of at least 98%, Hardy-Weinberg equilibrium P-value of at least $10x10^{-06}$, fewer than 1,000 Mendelian errors, and a minor allele frequency (MAF) of at least 1%. For this analysis, we further excluded SNPs with low imputation quality ($R^2 \leq 0.8$) and those with a genotype missing rate greater than 2%. To adjust for population stratification in the FHS dataset, we applied the same approach used for defining relatedness and principal components in the ADSP dataset. Details of this methodology are provided in **Supplementary Note 7.**

#### Supplementary Note 3: Korean Brain Aging Study for the Early Diagnosis and Prediction of Alzheimer's Disease (KBASE) genotyping methods and quality control

WGS data for 603 KBASE participants were obtained from the GCAD release of the Alzheimer's Disease Sequencing Project (ADSP) release 5, available through NIAGADS (accession NG00067). VCF files were imported into a MatrixTable using HAIL version 0.2.133 and filtered to retain only PASS variants with call rate > 0.99, minor allele frequency (MAF) ≥ 0.01, genotype missingness ≤ 0.01, and Hardy–Weinberg equilibrium $P \geq 1\times10^{-6}$. The resulting filtered data was exported to both VCF and PLINK (BED) formats for downstream analysis. To adjust for population stratification in the KBASE dataset, we applied the same approach used for defining relatedness and principal components in the ADSP dataset. Details of this methodology are provided in **Supplementary Note 7.**

#### Supplementary Note 4: Rush University Medical Center cohorts genotyping and quality control

The genotyping, quality control, imputation procedures, and relatedness checks for the Rush University Medical Center cohorts (Minority Aging Research Study, ROSMAP, Rush Alzheimer's Disease Center) have been previously described. Briefly, genotyping was performed using three arrays (Global Screening Array-24 v3.0, Affymetrix GeneChip 6.0, and Illumina HumanOmniExpress)[30,31]. Imputation was conducted using the TOPMed reference panel (hg38)[32]. For PRS construction, we excluded variants with low genotype rates (≤99%), low minor allele frequency (MAF <1%), and from Hardy-Weinberg equilibrium ($P < 1\times10^{-6}$).

#### Supplementary Note 5: All of Us ancestry analysis methods and genotype data quality control

We utilized WGS data from the All of Us (AoU) research program, version 7, which includes 245,394 samples from diverse U.S. populations. Detailed information on sequencing and quality control can be found (https://support.researchallofus.org/hc/en-us/categories/4537007565204-Genomics The WGS data were provided in Variant Dataset (VDS) format. Ancestry was predicted using a principal component analysis (PCA)-based approach, with categorical ancestry definitions aligned with those used in gnomAD and the 1000 Genomes Project, including African/African American (afr), American Admixed/Latino (amr), East Asian (eas), European (eur), Middle Eastern (mid), South Asian (sas), and Other (oth; not belonging to one of the other categories or representing a balanced admixture). This information was provided by the AoU team, and Further details on ancestry prediction, PCA, and relatedness are available at (https://support.researchallofus.org/hc/en-us/categories/4537007565204-Genomics). We filtered the variant datasets (VDS), focusing only on HapMap3 SNPs, using the Spark environment and Hail version 0.2.130 on the AoU Workbench. We subset the data to include only individuals of non-European ancestry, following the instructions in the AoU Workbench documentation under "How to Work with All of Us Genomic Data (Hail - Plink) (v7)." Briefly, we subset the SNPs and non-European samples, converted them into a Hail matrix table, filtered for passing variants, and applied a missing call rate threshold of 2%. The resulting data were then converted to BED file format. For European ancestry, we used available WGS data provided in Community Workspaces under the Polygenic Risk Scores and Physical Activity CW, as previously described [33]. This dataset contains 133,581 European samples and HapMap3 SNPs in Plink format with variant positions in Hg37. Variants from non-European ancestry samples were re-coded to Hg37 using the UCSC hg19 to hg38 chain file via the rtracklayer R package version 1.46.0 [34]. The non-European and European BED files were then merged using PLINK v1.9 [28], and we applied filters for minor allele frequency (MAF) > 0.01 and a missing genotype threshold of 2%.

#### Supplementary Note 6: Ancestry analysis of ADSP participants

We defined ancestry based on genetic similarity using principal component analysis (PCA). Individuals were grouped into super populations as defined by the 1000 Genomes Project (1KG), which includes European, African, American Indian, South Asian, and East Asian populations [35]. To refine the ancestry classification, we incorporated self-reported race/ethnicity information. A substantial number of individuals identified as Hispanic or Latino, reflecting the known admixture within these populations. Accordingly, we further classified American Indian individuals into Caribbean Hispanic/Latino and Native American Hispanic/Latino groups, following the classifications in 1KG. Each ADSP sample was projected onto the principal components (PCs) derived from the 1KG reference panel. To prepare the reference dataset, we filtered 1KG samples to include only variants with a minor allele frequency (MAF) > 0.01. We removed long-range linkage disequilibrium (LD) regions, including chr6: 25–35 Mb and chr8: 7–13 Mb, and excluded related individuals, retaining only one individual from each pair with a second-degree or closer relationship[36].

The ADSP genotype data were filtered to include variants with MAF > 1%. We merged the filtered ADSP data with the 1KG reference dataset, retaining only single nucleotide polymorphisms (SNPs) common to both datasets. Individuals with genotype missing rates > 0.02 were excluded. To further improve data quality, LD pruning was performed using PLINK v1.9 with the parameters --indep-pairwise 500 50 0.1. PCA was then conducted on the merged dataset using the GENESIS R package [37], and ADSP samples were projected onto the top principal components calculated from the 1KG reference panel.

To assign ancestry, we trained a random forest model using the top 6 PCs from the 1KG dataset. ADSP samples were classified into six 1KG population groups, with ancestry assigned when the predicted probability for a given population exceeded 0.8. Additionally, we performed ancestry inference using the five super populations from the 1KG, applying the same data to compute the PCs. We followed the guidelines from SCOPE software to conduct supervised population structure inference for each individual in the ADSP [38].

#### Supplementary Note 7: Kinship and Population Structure analyses of ADSP participants

To adjust for population stratification in our analysis, we followed the established TOPMed Pipeline (https://github.com/UW-GAC/analysis_pipeline). Briefly, we performed principal component analysis (PCA) using high-quality SNPs, applying stringent filters such as minor allele frequency (MAF) ≥ 0.01, genotype missing rate ≤ 0.01, and limiting the analysis to autosomal chromosomes. SNPs were pruned using PLINK v1.9 [28] with the parameters --indep-pairwise 500 50 0.1 to minimize linkage disequilibrium. The estimate of unrelated individuals was identified using KING version 2.3.1 [26] which estimates kinship for close relatives using the "IBD" method and provides a full matrix of population divergence estimates for all sample pairs. We used the pcairPartition function from the GENESIS R package version 2.16.1 [37] to select a set of unrelated samples, applying a third-degree kinship threshold of 0.044. Subsequently, we computed PCA for all samples using the GENESIS R package version 2.16.1, which accounts for relatedness in the analysis. Finally, we computed the kinship matrix while adjusting for population structure, ensuring proper correction for population stratification in our downstream analyses.

#### Supplementary Note 8: Acknowledgment for ADSP

The Alzheimer's Disease Sequencing Project (ADSP) is comprised of two Alzheimer's Disease (AD) genetics consortia and three National Human Genome Research Institute (NHGRI) funded Large Scale Sequencing and Analysis Centers (LSAC). The two AD genetics consortia are the Alzheimer's Disease Genetics Consortium (ADGC) funded by NIA (U01 AG032984), and the Cohorts for Heart and Aging Research in Genomic Epidemiology (CHARGE) funded by NIA (R01 AG033193), the National Heart, Lung, and Blood Institute (NHLBI), other National Institute of Health (NIH) institutes and other foreign governmental and non-governmental organizations. The Discovery Phase analysis of sequence data is supported through UF1AG047133 (to Drs. Schellenberg, Farrer, Pericak-Vance, Mayeux, and Haines); U01AG049505 to Dr. Seshadri; U01AG049506 to Dr. Boerwinkle; U01AG049507 to Dr. Wijsman; and U01AG049508 to Dr. Goate and the Discovery Extension Phase analysis is supported through U01AG052411 to Dr. Goate, U01AG052410 to Dr. Pericak-Vance and U01 AG052409 to Drs. Seshadri and Fornage.

Sequencing for the Follow Up Study (FUS) is supported through U01AG057659 (to Drs. PericakVance, Mayeux, and Vardarajan) and U01AG062943 (to Drs. Pericak-Vance and Mayeux). Data generation and harmonization in the Follow-up Phase is supported by U54AG052427 (to Drs. Schellenberg and Wang). The FUS Phase analysis of sequence data is supported through U01AG058589 (to Drs. Destefano, Boerwinkle, De Jager, Fornage, Seshadri, and Wijsman), U01AG058654 (to Drs. Haines, Bush, Farrer, Martin, and Pericak-Vance), U01AG058635 (to Dr. Goate), RF1AG058066 (to Drs. Haines, Pericak-Vance, and Scott), RF1AG057519 (to Drs. Farrer and Jun), R01AG048927 (to Dr. Farrer), and RF1AG054074 (to Drs. Pericak-Vance and Beecham).

The ADGC cohorts include: Adult Changes in Thought (ACT) (U01 AG006781, U19 AG066567), the Alzheimer's Disease Research Centers (ADRC) (P30 AG062429, P30 AG066468, P30 AG062421, P30 AG066509, P30 AG066514, P30 AG066530, P30 AG066507, P30 AG066444, P30 AG066518, P30 AG066512, P30 AG066462, P30 AG072979, P30 AG072972, P30 AG072976, P30 AG072975, P30 AG072978, P30 AG072977, P30 AG066519, P30 AG062677, P30 AG079280, P30 AG062422, P30

AG066511, P30 AG072946, P30 AG062715, P30 AG072973, P30 AG066506, P30 AG066508, P30 AG066515, P30 AG072947, P30 AG072931, P30 AG066546, P20 AG068024, P20 AG068053, P20 AG068077, P20 AG068082, P30 AG072958, P30 AG072959), the Chicago Health and Aging Project (CHAP) (R01 AG11101, RC4 AG039085, K23 AG030944), Indiana Memory and Aging Study (IMAS) (R01 AG019771), Indianapolis Ibadan (R01 AG009956, P30 AG010133), the Memory and Aging Project (MAP) ( R01 AG17917), Mayo Clinic (MAYO) (R01 AG032990, U01 AG046139, R01 NS080820, RF1 AG051504, P50 AG016574), Mayo Parkinson’s Disease controls (NS039764, NS071674, 5RC2HG005605), University of Miami (R01 AG027944, R01 AG028786, R01 AG019085, IIRG09133827, A2011048), the Multi-Institutional Research in Alzheimer’s Genetic Epidemiology Study (MIRAGE) (R01 AG09029, R01 AG025259), the National Centralized Repository for Alzheimer’s Disease and Related Dementias (NCRAD) (U24 AG021886), the National Institute on Aging Late Onset Alzheimer’s Disease Family Study (NIA- LOAD) (U24 AG056270), the Religious Orders Study (ROS) (P30 AG10161, R01 AG15819), the Texas Alzheimer’s Research and Care Consortium (TARCC) (funded by the Darrell K Royal Texas Alzheimer’s Initiative), Vanderbilt University/Case Western Reserve University (VAN/CWRU) (R01 AG019757, R01 AG021547, R01 AG027944, R01 AG028786, P01 NS026630, and Alzheimer’s Association), the Washington Heights-Inwood Columbia Aging Project (WHICAP) (RF1 AG054023), the University of Washington Families (VA Research Merit Grant, NIA: P50AG005136, R01AG041797, NINDS: R01NS069719), the Columbia University Hispanic Estudio Familiar de Influencia Genetica de Alzheimer (EFIGA) (RF1 AG015473), the University of Toronto (UT) (funded by Wellcome Trust, Medical Research Council, Canadian Institutes of Health Research), and Genetic Differences (GD) (R01 AG007584).

The CHARGE cohorts are supported in part by National Heart, Lung, and Blood Institute (NHLBI) infrastructure grant HL105756 (Psaty), RC2HL102419 (Boerwinkle) and the neurology working group is supported by the National Institute on Aging (NIA) R01 grant AG033193. The CHARGE cohorts participating in the ADSP include the following: Austrian Stroke Prevention Study (ASPS), ASPS-Family study, and the Prospective Dementia Registry-Austria (ASPS/PRODEM-Aus), the Atherosclerosis Risk in Communities (ARIC) Study, the Cardiovascular Health Study (CHS), the Erasmus Rucphen Family Study (ERF), the Framingham Heart Study (FHS), and the Rotterdam Study (RS). ASPS is funded by the Austrian Science Fond (FWF) grant number P20545-P05 and P13180 and the Medical University of Graz. The ASPS-Fam is funded by the Austrian Science Fund (FWF) project I904), the EU Joint Programme – Neurodegenerative Disease Research (JPND) in frame of the BRIDGET project (Austria, Ministry of Science) and the Medical University of Graz and the Steiermärkische Krankenanstalten Gesellschaft. PRODEM-Austria is supported by the Austrian Research Promotion agency (FFG) (Project No. 827462) and by the Austrian National Bank (Anniversary Fund, project 15435. ARIC research is carried out as a collaborative study supported by NHLBI contracts (HHSN268201100005C, HHSN268201100006C, HHSN268201100007C, HHSN268201100008C, HHSN268201100009C, HHSN268201100010C, HHSN268201100011C, and HHSN268201100012C). Neurocognitive data in ARIC is collected by U01 2U01HL096812, 2U01HL096814, 2U01HL096899, 2U01HL096902, 2U01HL096917 from the NIH (NHLBI, NINDS, NIA and NIDCD), and with previous brain MRI examinations funded by R01-HL70825 from the NHLBI. CHS research was supported by contracts HHSN268201200036C, HHSN268200800007C, N01HC55222, N01HC85079, N01HC85080, N01HC85081, N01HC85082, N01HC85083, N01HC85086, and grants U01HL080295 and U01HL130114 from the NHLBI with additional contribution from the National Institute of Neurological Disorders and Stroke (NINDS). Additional support was provided by R01AG023629, R01AG15928, and R01AG20098 from the NIA. FHS research is supported by NHLBI contracts N01-HC-25195 and HHSN268201500001I. This study was also supported by additional grants from the NIA (R01s AG054076, AG049607 and AG033040 and NINDS (R01 NS017950). The ERF study as a part of EUROSPAN (European Special Populations Research Network) was supported by European Commission FP6 STRP grant number 018947 (LSHG-CT-2006-01947) and also received funding from the European Community's Seventh Framework Programme (FP7/2007-2013)/grant agreement HEALTH-F4- 2007-201413 by the European Commission under the programme "Quality of Life and Management of the Living Resources" of 5th Framework Programme (no.

QLG2-CT-2002- 01254). High-throughput analysis of the ERF data was supported by a joint grant from the Netherlands Organization for Scientific Research and the Russian Foundation for Basic Research (NWO-RFBR 047.017.043). The Rotterdam Study is funded by Erasmus Medical Center and Erasmus University, Rotterdam, the Netherlands Organization for Health Research and Development (ZonMw), the Research Institute for Diseases in the Elderly (RIDE), the Ministry of Education, Culture and Science, the Ministry for Health, Welfare and Sports, the European Commission (DG XII), and the municipality of Rotterdam. Genetic data sets are also supported by the Netherlands Organization of Scientific Research NWO Investments (175.010.2005.011, 911-03-012), the Genetic Laboratory of the Department of Internal Medicine, Erasmus MC, the Research Institute for Diseases in the Elderly (014-93-015; RIDE2), and the Netherlands Genomics Initiative (NGI)/Netherlands Organization for Scientific Research (NWO) Netherlands Consortium for Healthy Aging (NCHA), project 050-060-810. All studies are grateful to their participants, faculty and staff. The content of these manuscripts is solely the responsibility of the authors and does not necessarily represent the official views of the National Institutes of Health or the U.S. Department of Health and Human Services.

The FUS cohorts include: the Alzheimer's Disease Research Centers (ADRC) (P30 AG062429, P30 AG066468, P30 AG062421, P30 AG066509, P30 AG066514, P30 AG066530, P30 AG066507, P30 AG066444, P30 AG066518, P30 AG066512, P30 AG066462, P30 AG072979, P30 AG072972, P30 AG072976, P30 AG072975, P30 AG072978, P30 AG072977, P30 AG066519, P30 AG062677, P30 AG079280, P30 AG062422, P30 AG066511, P30 AG072946, P30 AG062715, P30 AG072973, P30 AG066506, P30 AG066508, P30 AG066515, P30 AG072947, P30 AG072931, P30 AG066546, P20 AG068024, P20 AG068053, P20 AG068077, P20 AG068082, P30 AG072958, P30 AG072959), Alzheimer's Disease Neuroimaging Initiative (ADNI) (U19AG024904), Amish Protective Variant Study (RF1AG058066), Cache County Study (R01AG11380, R01AG031272, R01AG21136, RF1AG054052), Case Western Reserve University Brain Bank (CWRUBB) (P50AG008012), Case Western Reserve University Rapid Decline (CWRURD) (RF1AG058267, NU38CK000480), CubanAmerican Alzheimer's Disease Initiative (CuAADI) (3U01AG052410), Estudio Familiar de

Influencia Genetica en Alzheimer (EFIGA) (5R37AG015473, RF1AG015473, R56AG051876), Genetic and Environmental Risk Factors for Alzheimer Disease Among African Americans Study (GenerAAtions) (2R01AG09029, R01AG025259, 2R01AG048927), Gwangju Alzheimer and Related Dementias Study (GARD) (U01AG062602), Hillblom Aging Network (2014-A-004-NET, R01AG032289, R01AG048234), Hussman Institute for Human Genomics Brain Bank (HIHGBB) (R01AG027944, Alzheimer's Association "Identification of Rare Variants in Alzheimer Disease"), Ibadan Study of Aging (IBADAN) (5R01AG009956), Longevity Genes Project (LGP) and LonGenity (R01AG042188, R01AG044829, R01AG046949, R01AG057909, R01AG061155, P30AG038072), Mexican Health and Aging Study (MHAS) (R01AG018016), Multi-Institutional Research in Alzheimer's Genetic Epidemiology (MIRAGE) (2R01AG09029, R01AG025259, 2R01AG048927), Northern Manhattan Study (NOMAS) (R01NS29993), Peru Alzheimer's Disease Initiative (PeADI) (RF1AG054074), Puerto Rican 1066 (PR1066) (Wellcome Trust (GR066133/GR080002), European Research Council (340755)), Puerto Rican Alzheimer Disease Initiative (PRADI) (RF1AG054074), Reasons for Geographic and Racial Differences in Stroke (REGARDS) (U01NS041588), Research in African American Alzheimer Disease Initiative (REAAADI) (U01AG052410), the Religious Orders Study (ROS) (P30 AG10161, P30 AG72975, R01 AG15819, R01 AG42210), the RUSH Memory and Aging Project (MAP) (R01 AG017917, R01 AG42210Stanford Extreme Phenotypes in AD (R01AG060747), University of Miami Brain Endowment Bank (MBB), University of Miami/Case Western/North Carolina A&T African American (UM/CASE/NCAT) (U01AG052410, R01AG028786), Wisconsin Registry for Alzheimer's Prevention (WRAP) (R01AG027161 and R01AG054047), Mexico-Southern California Autosomal Dominant Alzheimer's Disease Consortium (R01AG069013), Center for Cognitive Neuroscience and Aging (R01AG047649), and the A4 Study (R01AG063689, U19AG010483 and U24AG057437).

The four LSACs are: the Human Genome Sequencing Center at the Baylor College of Medicine (U54 HG003273), the Broad Institute Genome Center (U54HG003067), The American Genome Center at the Uniformed Services University of the Health Sciences (U01AG057659), and the Washington University Genome

Institute (U54HG003079). Genotyping and sequencing for the ADSP FUS is also conducted at John P. Hussman Institute for Human Genomics (HIHG) Center for Genome Technology (CGT).

Biological samples and associated phenotypic data used in primary data analyses were stored at Study Investigators institutions, and at the National Centralized Repository for Alzheimer's Disease and Related Dementias (NCRAD, U24AG021886) at Indiana University funded by NIA. Associated Phenotypic Data used in primary and secondary data analyses were provided by Study Investigators, the NIA funded Alzheimer's Disease Centers (ADCs), and the National Alzheimer's Coordinating Center (NACC, U24AG072122) and the National Institute on Aging Genetics of Alzheimer's Disease Data Storage Site (NIAGADS, U24AG041689) at the University of Pennsylvania, funded by NIA. Harmonized phenotypes were provided by the ADSP Phenotype Harmonization Consortium (ADSP-PHC), funded by NIA (U24 AG074855, U01 AG068057 and R01 AG059716) and Ultrascale Machine Learning to Empower Discovery in Alzheimer's Disease Biobanks (AI4AD, U01 AG068057). This research was supported in part by the Intramural Research Program of the National Institutes of health, National Library of Medicine. Contributors to the Genetic Analysis Data included Study Investigators on projects that were individually funded by NIA, and other NIH institutes, and by private U.S. organizations, or foreign governmental or nongovernmental organizations.

The ADSP Phenotype Harmonization Consortium (ADSP-PHC) is funded by NIA (U24 AG074855, U01 AG068057 and R01 AG059716). The harmonized cohorts within the ADSP-PHC include: the Anti-Amyloid Treatment in Asymptomatic Alzheimer's study (A4 Study), a secondary prevention trial in preclinical Alzheimer's disease, aiming to slow cognitive decline associated with brain amyloid accumulation in clinically normal older individuals. The A4 Study is funded by a public-private-philanthropic partnership, including funding from the National Institutes of Health-National Institute on Aging, Eli Lilly and Company, Alzheimer's Association, Accelerating Medicines Partnership, GHR Foundation, an anonymous foundation and additional private donors, with in-kind support from Avid and Cogstate. The companion observational Longitudinal Evaluation of Amyloid Risk and Neurodegeneration (LEARN) Study is funded by the Alzheimer's

Association and GHR Foundation. The A4 and LEARN Studies are led by Dr. Reisa Sperling at Brigham and Women's Hospital, Harvard Medical School and Dr. Paul Aisen at the Alzheimer's Therapeutic Research Institute (ATRI), University of Southern California. The A4 and LEARN Studies are coordinated by ATRI at the University of Southern California, and the data are made available through the Laboratory for Neuro Imaging at the University of Southern California. The participants screening for the A4 Study provided permission to share their de-identified data in order to advance the quest to find a successful treatment for Alzheimer's disease. We would like to acknowledge the dedication of all the participants, the site personnel, and all of the partnership team members who continue to make the A4 and LEARN Studies possible. The complete A4 Study Team list is available on: a4study.org/a4-study-team.; the Adult Changes in Thought study (ACT), U01 AG006781, U19 AG066567; Alzheimer's Disease Neuroimaging Initiative (ADNI): Data collection and sharing for this project was funded by the Alzheimer's Disease Neuroimaging Initiative (ADNI) (National Institutes of Health Grant U01 AG024904) and DOD ADNI (Department of Defense award number W81XWH-12-2-0012). ADNI is funded by the National Institute on Aging, the National Institute of Biomedical Imaging and Bioengineering, and through generous contributions from the following: AbbVie, Alzheimer's Association; Alzheimer's Drug Discovery Foundation; Araclon Biotech; BioClinica, Inc.; Biogen; Bristol-Myers Squibb Company; CereSpir, Inc.; Cogstate; Eisai Inc.; Elan Pharmaceuticals, Inc.; Eli Lilly and Company; EuroImmun; F. Hoffmann-La Roche Ltd and its affiliated company Genentech, Inc.; Fujirebio; GE Healthcare; IXICO Ltd.;Janssen Alzheimer Immunotherapy Research & Development, LLC.; Johnson & Johnson Pharmaceutical Research & Development LLC.; Lumosity; Lundbeck; Merck & Co., Inc.;Meso Scale Diagnostics, LLC.; NeuroRx Research; Neurotrack Technologies; Novartis Pharmaceuticals Corporation; Pfizer Inc.; Piramal Imaging; Servier; Takeda Pharmaceutical Company; and Transition Therapeutics. The Canadian Institutes of Health Research is providing funds to support ADNI clinical sites in Canada. Private sector contributions are facilitated by the Foundation for the National Institutes of Health (www.fnih.org). The grantee organization is the Northern California Institute for Research and Education, and the study is coordinated by the Alzheimer's Therapeutic Research Institute at the University of Southern California. ADNI data are disseminated by the Laboratory for Neuro Imaging at the University of Southern California; Estudio Familiar de Influencia Genetica en Alzheimer (EFIGA): 5R37AG015473, RF1AG015473, R56AG051876; the Health & Aging Brain Study – Health Disparities (HABS-HD), supported by the National Institute on Aging of the National Institutes of Health under Award Numbers R01AG054073, R01AG058533, R01AG070862, P41EB015922, and U19AG078109; the Korean Brain Aging Study for the Early Diagnosis and Prediction of Alzheimer's disease (KBASE), which was supported by a grant from Ministry of Science, ICT and Future Planning (Grant No: NRF-2014M3C7A1046042); Memory & Aging Project at Knight Alzheimer's Disease Research Center (MAP at Knight ADRC): The Memory and Aging Project at the Knight-ADRC (Knight-ADRC). This work was supported by the National Institutes of Health (NIH) grants R01AG064614, R01AG044546, RF1AG053303, RF1AG058501, U01AG058922 and R01AG064877 to Carlos Cruchaga. The recruitment and clinical characterization of research participants at Washington University was supported by NIH grants P30AG066444, P01AG03991, and P01AG026276. Data collection and sharing for this project was supported by NIH grants RF1AG054080, P30AG066462, R01AG064614 and U01AG052410. We thank the contributors who collected samples used in this study, as well as patients and their families, whose help and participation made this work possible. This work was supported by access to equipment made possible by the Hope Center for Neurological Disorders, the Neurogenomics and Informatics Center (NGI: https://neurogenomics.wustl.edu/) and the Departments of Neurology and Psychiatry at Washington University School of Medicine; National Alzheimer's Coordinating Center (NACC): The NACC database is funded by NIA/NIH Grant U24 AG072122. SCAN is a multi-institutional project that was funded as a U24 grant (AG067418) by the National Institute on Aging in May 2020. Data collected by SCAN and shared by NACC are contributed by the NIA-funded ADRCs as follows: P30 AG062429 (PI James Brewer, MD, PhD), P30 AG066468 (PI Oscar Lopez, MD), P30 AG062421 (PI Bradley Hyman, MD, PhD), P30 AG066509 (PI Thomas Grabowski, MD), P30 AG066514 (PI Mary Sano, PhD), P30 AG066530 (PI Helena Chui, MD), P30 AG066507 (PI Marilyn Albert, PhD), P30 AG066444 (PI John Morris, MD), P30 AG066518 (PI Jeffrey Kaye, MD), P30 AG066512 (PI Thomas Wisniewski, MD), P30

AG066462 (PI Scott Small, MD), P30 AG072979 (PI David Wolk, MD), P30 AG072972 (PI Charles DeCarli, MD), P30 AG072976 (PI Andrew Saykin, PsyD), P30 AG072975 (PI David Bennett, MD), P30 AG072978 (PI Neil Kowall, MD), P30 AG072977 (PI Robert Vassar, PhD), P30 AG066519 (PI Frank LaFerla, PhD), P30 AG062677 (PI Ronald Petersen, MD, PhD), P30 AG079280 (PI Eric Reiman, MD), P30 AG062422 (PI Gil Rabinovici, MD), P30 AG066511 (PI Allan Levey, MD, PhD), P30 AG072946 (PI Linda Van Eldik, PhD), P30 AG062715 (PI Sanjay Asthana, MD, FRCP), P30 AG072973 (PI Russell Swerdlow, MD), P30 AG066506 (PI Todd Golde, MD, PhD), P30 AG066508 (PI Stephen Strittmatter, MD, PhD), P30 AG066515 (PI Victor Henderson, MD, MS), P30 AG072947 (PI Suzanne Craft, PhD), P30 AG072931 (PI Henry Paulson, MD, PhD), P30 AG066546 (PI Sudha Seshadri, MD), P20 AG068024 (PI Erik Roberson, MD, PhD), P20 AG068053 (PI Justin Miller, PhD), P20 AG068077 (PI Gary Rosenberg, MD), P20 AG068082 (PI Angela Jefferson, PhD), P30 AG072958 (PI Heather Whitson, MD), P30 AG072959 (PI James Leverenz, MD); National Institute on Aging Alzheimer's Disease Family Based Study (NIA-AD FBS): U24 AG056270; Religious Orders Study (ROS): P30AG10161,R01AG15819, R01AG42210; Memory and Aging Project (MAP - Rush): R01AG017917, R01AG42210; Minority Aging Research Study (MARS): R01AG22018, R01AG42210; the Texas Alzheimer's Research and Care Consortium (TARCC), funded by the Darrell K Royal Texas Alzheimer's Initiative, directed by the Texas Council on Alzheimer's Disease and Related Disorders; Washington Heights/Inwood Columbia Aging Project (WHICAP): RF1 AG054023;and Wisconsin Registry for Alzheimer's Prevention (WRAP): R01AG027161 and R01AG054047. Additional acknowledgments include the National Institute on Aging Genetics of Alzheimer's Disease Data Storage Site (NIAGADS, U24AG041689) at the University of Pennsylvania, funded by NIA.

#### Supplementary Note 9: Acknowledgment for ADGC

The National Institutes of Health, National Institute on Aging (NIH-NIA) supported this work through the following grants: ADGC, U01 AG032984, RC2 AG036528; Samples from the National Cell Repository for Alzheimer's Disease (NCRAD), which receives government support under a cooperative agreement grant (U24 AG21886) awarded by the National Institute on Aging (NIA), were used in this study.

We thank contributors who collected samples used in this study, as well as patients and their families, whose help and participation made this work possible; Data for this study were prepared, archived, and distributed by the National Institute on Aging Alzheimer's Disease Data Storage Site (NIAGADS) at the University of Pennsylvania (U24-AG041689-01); NACC, U01 AG016976; NIA LOAD, U24 AG026395, R01AG041797; Banner Sun Health Research Institute P30 AG019610; Boston University, P30 AG013846, U01 AG10483, R01 CA129769, R01 MH080295, R01 AG017173, R01 AG025259, R01AG33193; Columbia University, P50 AG008702, R37 AG015473; Duke University, P30 AG028377, AG05128; Emory University, AG025688; Group Health Research Institute, UO1 AG006781, UO1 HG004610, UO1 HG006375; Indiana University, P30 AG10133; Johns Hopkins University, P50 AG005146, R01 AG020688; Massachusetts General Hospital, P50 AG005134; Mayo Clinic, P50 AG016574; Mount Sinai School of Medicine, P50 AG005138, P01 AG002219; New York University, P30 AG08051, UL1 RR029893, 5R01AG012101, 5R01AG022374, 5R01AG013616, 1RC2AG036502, 1R01AG035137; Northwestern University, P30 AG013854; Oregon Health & Science University, P30 AG008017, R01 AG026916; Rush University, P30 AG010161, R01 AG019085, R01 AG15819, R01 AG17917, R01 AG30146; TGen, R01 NS059873; University of Alabama at Birmingham, P50 AG016582; University of Arizona, R01 AG031581; University of California, Davis, P30 AG010129; University of California, Irvine, P50 AG016573; University of California, Los Angeles, P50 AG016570; University of California, San Diego, P50 AG005131; University of California, San Francisco, P50 AG023501, P01 AG019724; University of Kentucky, P30 AG028383, AG05144; University of Michigan, P50 AG008671; University of Pennsylvania, P30 AG010124; University of Pittsburgh, P50 AG005133, AG030653, AG041718, AG07562, AG02365; University of Southern California, P50 AG005142;

University of Texas Southwestern, P30 AG012300; University of Miami, R01 AG027944, AG010491, AG027944, AG021547, AG019757; University of Washington, P50 AG005136; University of Wisconsin, P50 AG033514; Vanderbilt University, R01 AG019085; and Washington University, P50 AG005681, P01 AG03991.

The Kathleen Price Bryan Brain Bank at Duke University Medical Center is funded by NINDS grant # NS39764, NIMH MH60451 and by Glaxo Smith Kline. Genotyping of the TGEN2 cohort was supported by Kronos Science. The TGen series was also funded by NIA grant AG041232 to AJM and MJH, The Banner Alzheimer's Foundation, The Johnnie B. Byrd Sr. Alzheimer's Institute, the Medical Research Council, and the state of Arizona and also includes samples from the following sites: Newcastle Brain Tissue Resource (funding via the Medical Research Council, local NHS trusts and Newcastle University), MRC London Brain Bank for Neurodegenerative Diseases (funding via the Medical Research Council),South West Dementia Brain Bank (funding via numerous sources including the Higher Education Funding Council for England (HEFCE), Alzheimer's Research Trust (ART), BRACE as well as North Bristol NHS Trust Research and Innovation Department and DeNDRoN), The Netherlands Brain Bank (funding via numerous sources including Stichting MS Research, Brain Net Europe, Hersenstichting Nederland Breinbrekend Werk, International Parkinson Fonds, Internationale Stiching Alzheimer Onderzoek), Institut de Neuropatologia, Servei Anatomia Patologica, Universitat de Barcelona.

ADNI data collection and sharing was funded by the National Institutes of Health Grant U01 AG024904 and Department of Defense award number W81XWH-12-2-0012. ADNI is funded by the National Institute on Aging, the National Institute of Biomedical Imaging and Bioengineering, and through generous contributions from the following: AbbVie, Alzheimer's Association; Alzheimer's Drug Discovery Foundation; Araclon Biotech; BioClinica, Inc.; Biogen; Bristol-Myers Squibb Company; CereSpir, Inc.; Eisai Inc.; Elan Pharmaceuticals, Inc.; Eli Lilly and Company; EuroImmun; F. Hoffmann-La Roche Ltd and its affiliated company Genentech, Inc.; Fujirebio; GE Healthcare; IXICO Ltd.; Janssen Alzheimer Immunotherapy Research & Development, LLC.; Johnson & Johnson Pharmaceutical Research & Development LLC.; Lumosity; Lundbeck; Merck & Co., Inc.; Meso Scale Diagnostics, LLC.; NeuroRx Research; Neurotrack Technologies;

Novartis Pharmaceuticals Corporation; Pfizer Inc.; Piramal Imaging; Servier; Takeda Pharmaceutical Company; and Transition Therapeutics.

The Canadian Institutes of Health Research is providing funds to support ADNI clinical sites in Canada. Private sector contributions are facilitated by the Foundation for the National Institutes of Health (www.fnih.org). The grantee organization is the Northern California Institute for Research and Education, and the study is coordinated by the Alzheimer's Disease Cooperative Study at the University of California, San Diego. ADNI data are disseminated by the Laboratory for Neuro Imaging at the University of Southern California. We thank Drs. D. Stephen Snyder and Marilyn Miller from NIA who are *ex-officio* ADGC members.

Support was also from the Alzheimer’s Association (LAF, IIRG-08-89720; MP-V, IIRG-05-14147) and the US Department of Veterans Affairs Administration, Office of Research and Development, Biomedical Laboratory Research Program. P.S.G.-H. is supported by Wellcome Trust, Howard Hughes Medical Institute, and the Canadian Institute of Health Research.

#### Supplementary Note 10: Acknowledgement for VA Million Veteran Program Core

**MVP Program Office**

- Sumitra Muralidhar, PhD, Program Director, US Department of Veterans Affairs, 810 Vermont Avenue NW, Washington, DC 20420
- Jennifer Moser, PhD, Associate Director, Scientific Programs, US Department of Veterans Affairs, 810 Vermont Avenue NW, Washington, DC 20420
- Jennifer E. Deen, BS, Associate Director, Cohort & Public Relations, US Department of Veterans Affairs, 810 Vermont Avenue NW, Washington, DC 20420

**MVP Executive Committee**

- Co-Chair: Philip S. Tsao, PhD, VA Palo Alto Health Care System, 3801 Miranda Avenue, Palo Alto, CA 94304
- Co-Chair: Sumitra Muralidhar, PhD, US Department of Veterans Affairs, 810 Vermont Avenue NW, Washington, DC 20420
- J. Michael Gaziano, MD, MPH, VA Boston Healthcare System, 150 S. Huntington Avenue, Boston, MA 02130
- Elizabeth Hauser, PhD, Durham VA Medical Center, 508 Fulton Street, Durham, NC 27705
- Amy Kilbourne, PhD, MPH, VA HSR&D, 2215 Fuller Road, Ann Arbor, MI 48105
- Michael Matheny, MD, MS, MPH, VA Tennessee Valley Healthcare System, 1310 24th Ave. South, Nashville, TN 37212
- Dave Oslin, MD, Philadelphia VA Medical Center, 3900 Woodland Avenue, Philadelphia, PA 19104
- Deepak Voora, MD, Durham VA Medical Center, 508 Fulton Street, Durham, NC 27705

**MVP Co-Principal Investigators**

- J. Michael Gaziano, MD, MPH, VA Boston Healthcare System, 150 S. Huntington Avenue, Boston, MA 02130
- Philip S. Tsao, PhD, VA Palo Alto Health Care System, 3801 Miranda Avenue, Palo

Alto, CA 94304

**MVP Core Operations**

- Jessica V. Brewer, MPH, Director, MVP Cohort Operations, VA Boston Healthcare System, 150 S. Huntington Avenue, Boston, MA 02130
- Mary T. Brophy MD, MPH, Director, VA Central Biorepository, VA Boston Healthcare System, 150 S. Huntington Avenue, Boston, MA 02130
- Kelly Cho, MPH, PhD, Director, MVP Phenomics, VA Boston Healthcare System, 150 S. Huntington Avenue, Boston, MA 02130
- Lori Churby, BS, Director, MVP Regulatory Affairs, VA Palo Alto Health Care System, 3801 Miranda Avenue, Palo Alto, CA 94304
- Scott L. DuVall, PhD, Director, VA Informatics and Computing Infrastructure, (VINCI) VA Salt Lake City Health Care System, 500 Foothill Drive, Salt Lake City, UT 84148
- Saiju Pyarajan PhD, Director, Data and Computational Sciences, VA Boston Healthcare System, 150 S. Huntington Avenue, Boston, MA 02130
- Robert Ringer, PharmD, Director, VA Albuquerque Central Biorepository, New Mexico VA Health Care System, 1501 San Pedro Drive SE, Albuquerque, NM 87108
- Luis E. Selva, PhD, Director, MVP Biorepository Coordination, VA Boston Healthcare System, 150 S. Huntington Avenue, Boston, MA 02130
- Shahpoor (Alex) Shayan, MS, Director, MVP PRE Informatics, VA Boston Healthcare System, 150 S. Huntington Avenue, Boston, MA 02130
- Brady Stephens, MS, Principal Investigator, MVP Information Center, Canandaigua VA Medical Center, 400 Fort Hill Avenue, Canandaigua, NY 14424
- Stacey B. Whitbourne, PhD, Director, MVP Cohort Development and Management VA Boston Healthcare System, 150 S. Huntington Avenue, Boston, MA 02130
